# Clinical Outcomes, Costs, and Cost-effectiveness of Strategies for People Experiencing Sheltered Homelessness During the COVID-19 Pandemic

**DOI:** 10.1101/2020.08.07.20170498

**Authors:** Travis P. Baggett, Justine A. Scott, Mylinh H. Le, Fatma M. Shebl, Christopher Panella, Elena Losina, Clare Flanagan, Jessie M. Gaeta, Anne Neilan, Emily P. Hyle, Amir Mohareb, Krishna P. Reddy, Mark J. Siedner, Guy Harling, Milton C. Weinstein, Andrea Ciaranello, Pooyan Kazemian, Kenneth A. Freedberg

## Abstract

**Importance:** Approximately 356,000 people stay in homeless shelters nightly in the US. They are at high risk for COVID-19.

**Objective:** To assess clinical outcomes, costs, and cost-effectiveness of strategies for COVID-19 management among sheltered homeless adults.

**Design:** We developed a dynamic microsimulation model of COVID-19 in sheltered homeless adults in Boston, Massachusetts. We used cohort characteristics and costs from Boston Health Care for the Homeless Program. Disease progression, transmission, and outcomes data were from published literature and national databases. We examined surging, growing, and slowing epidemics (effective reproduction numbers [R_e_] 2.6, 1.3, and 0.9). Costs were from a health care sector perspective; time horizon was 4 months, from April to August 2020.

**Setting & Participants:** Simulated cohort of 2,258 adults residing in homeless shelters in Boston.

**Interventions:** We assessed daily symptom screening with polymerase chain reaction (PCR) testing of screen-positives, universal PCR testing every 2 weeks, hospital-based COVID-19 care, alternate care sites [ACSs] for mild/moderate COVID-19, and temporary housing, each compared to no intervention.

**Main Outcomes and Measures:** Cumulative infections and hospital-days, costs to the health care sector (US dollars), and cost-effectiveness, as incremental cost per case prevented of COVID-19.

**Results:** We simulated a population of 2,258 sheltered homeless adults with mean age of 42.6 years. Compared to no intervention, daily symptom screening with ACSs for pending tests or confirmed COVID-19 and mild/moderate disease led to 37% fewer infections and 46% lower costs (R_e_=2.6), 75% fewer infections and 72% lower costs (R_e_=1.3), and 51% fewer infections and 51% lower costs (R_e_=0.9). Adding PCR testing every 2 weeks further decreased infections; incremental cost per case prevented was $1,000 (R_e_=2.6), $27,000 (R_e_=1.3), and $71,000 (R_e_=0.9). Temporary housing with PCR every 2 weeks was most effective but substantially more costly than other options. Results were sensitive to cost and sensitivity of PCR and ACS efficacy in preventing transmission.

**Conclusions & Relevance:** In this modeling study of simulated adults living in homeless shelters, daily symptom screening and ACSs were associated with fewer COVID-19 infections and decreased costs compared with no intervention. In a modeled surging epidemic, adding universal PCR testing every 2 weeks was associated with further decrease in COVID-19 infections at modest incremental cost and should be considered during future surges.

**Key Points:** *Question:* What are the projected clinical outcomes and costs of strategies for reducing COVID-19 infections among people experiencing sheltered homelessness?

*Findings:* In this microsimulation modeling study, daily symptom screening with polymerase chain reaction (PCR) testing of screen-positive individuals, paired with non-hospital care site management of people with mild to moderate COVID-19, substantially reduced infections and lowered costs over 4 months compared to no intervention, across a wide range of epidemic scenarios. In a surging epidemic, adding periodic universal PCR testing to symptom screening and non-hospital care site management improved clinical outcomes at modestly increased costs. Periodic universal PCR testing paired with temporary housing further reduced infections but at much higher cost.

*Meaning:* Daily symptom screening with PCR testing of screen-positive individuals and use of alternate care sites for COVID-19 management among sheltered homeless people was associated with substantially reduced new cases and costs compared to other strategies.

## INTRODUCTION

Over 1.4 million people experience sheltered homelessness annually in the US, including approximately 356,000 each night.^1,2^ The crowded circumstances of homeless shelters place this population at increased risk for coronavirus disease 2019 (COVID-19). The United States (US) Centers for Disease Control and Prevention (CDC) issued comprehensive guidance for preventing and mitigating COVID-19 among people experiencing sheltered homelessness, including recommendations for infection control practices in shelters, symptom screening of shelter guests, and dedicated settings for isolation and management of individuals with symptoms or confirmed illness.^3^ The high burden of COVID-19 among sheltered homeless populations^4–7^ highlights an urgent need to understand the clinical outcomes and costs of CDC-recommended and other prevention and treatment strategies. After a cluster of COVID-19 cases at a single large shelter in Boston, universal polymerase chain reaction (PCR) testing of 408 shelter residents found that 36% had SARS-CoV-2 infection.^4^ Eighty-eight percent of these individuals reported no symptoms at the time of testing, raising questions about how to identify COVID-19 disease in this population and the role of non-hospital alternate care sites (ACSs) to isolate those who do not require hospitalization. Our objective was to project the clinical and economic impact of COVID-19 management approaches for adults experiencing sheltered homelessness.

## METHODS

### Analytic Overview

We developed the Clinical and Economic Analysis of COVID-19 interventions (CEACOV) model, a dynamic microsimulation of the natural history of COVID-19 disease and the impact of prevention, testing, and treatment interventions. We used CEACOV to project the clinical impact, costs, and cost-effectiveness of various COVID-19 management strategies for people experiencing sheltered homelessness, including different combinations of symptom screening, PCR testing, ACSs, and relocating all shelter residents to temporary housing. Using data from the early stage of an outbreak among homeless adults in Boston, Massachusetts, we modeled a cohort of sheltered homeless adults and examined management strategies under various epidemic scenarios, given evolving and heterogenous epidemic dynamics across the US.^4,8^ We evaluated 3 scenarios over a 4-month time horizon, from April to August 2020, with different effective reproduction numbers (R_e_) representing surging (R_e_=2.6), growing (R_e_=1.3), and slowing (R_e_=0.9) epidemics. Outcomes included number of infections, utilization of hospital and intensive care unit (ICU) beds, costs, and cost per COVID-19 case was conducted from a health care sector perspective. This study was approved by the Partners prevented. The analysis Human Research Committee.

### Model Structure

#### Disease states and progression

CEACOV is a dynamic microsimulation model of COVID-19 based on an SEIR framework, including susceptible, exposed, infectious, recovered, and death states.^9^ Infected individuals face daily probabilities of disease progression through 6 COVID-19 states: pre-infectious latency, asymptomatic, mild/moderate, severe, critical, and recuperation. With mild/moderate disease, individuals have mild symptoms, such as cough or fever, that generally do not require inpatient management in a stably housed population. With severe disease, symptoms warrant inpatient management. With critical disease, patients require ICU care. Recovered individuals cannot transmit and are assumed immune from repeat infection.^10^ eFigure1 displays how patients move through the model. We describe model validation in the Supplemental Methods.

#### Transmission

Individuals with COVID-19 transmit to susceptible individuals at health state-stratified rates. We model a closed cohort, with transmissions occurring between people experiencing sheltered homelessness. All susceptible people face equal probabilities of contacting infected individuals and becoming infected (homogenous mixing). The number of projected infections depends on COVID-19 prevalence, proportion of the population susceptible, transmission rates, and interventions that change contact rates or infectivity per contact. Transmission rates are calibrated to achieve the desired R_e_, which captures the average number of transmissions per case. More details can be found in the Supplemental Methods.

#### Testing and care interventions

Symptom screens or PCR tests are offered at intervals defined in each strategy; test sensitivities and specificities depend on COVID-19 health state. Care interventions include hospital care, ACSs, and temporary housing. Since adequate isolation for COVID-19 is not possible within congregate homeless shelters, care of homeless individuals with mild/moderate COVID-19 occurs either in hospitals or ACSs, such as large tents or non-hospital facilities with on-site medical staff.^11,12^ ACSs reduce transmission and hospital use for people with mild/moderate illness. Temporary housing reduces transmission by preemptively moving everyone from shelters to individual living units (e.g., hotel or dormitory rooms) for the entire simulation period. Anyone who develops mild/moderate COVID-19 remains in temporary housing, which offers health monitoring and space for isolation but less intensive staffing and infection control than ACSs.

#### Resource use, costs, cost-effectiveness, and budget impact

The model tallies resource utilization, including tests and days in hospital, ICU, ACS, or temporary housing, and daily costs, including medical supplies and personnel. We included a budget impact analysis to determine total costs over the 4-month simulation. To understand the tradeoffs between cost and infections prevented and highlight the relative “return on investment” for each strategy, we present efficiency frontiers, plotting number of infections prevented against total cost for each strategy.^13^ Since we focus on a cohort relevant to an individual city, and since overall COVID-19 mortality is low, we report incremental cost per COVID-19 case prevented as an outcome; $1,000/case prevented is approximately equivalent to $61,000/quality-adjusted life year (QALY) gained at current case fatality levels (Table 2, notes).

**Table 1.**
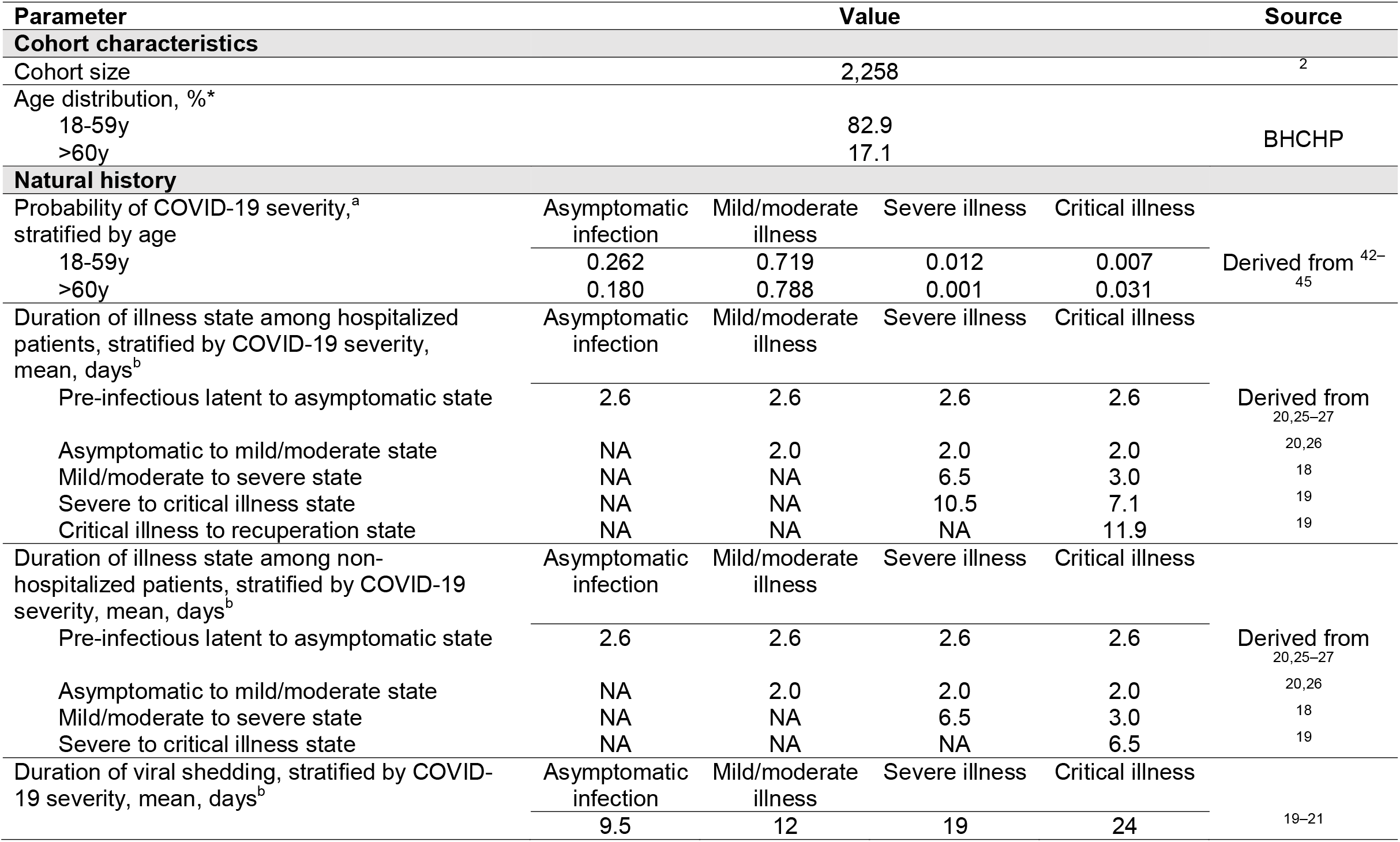
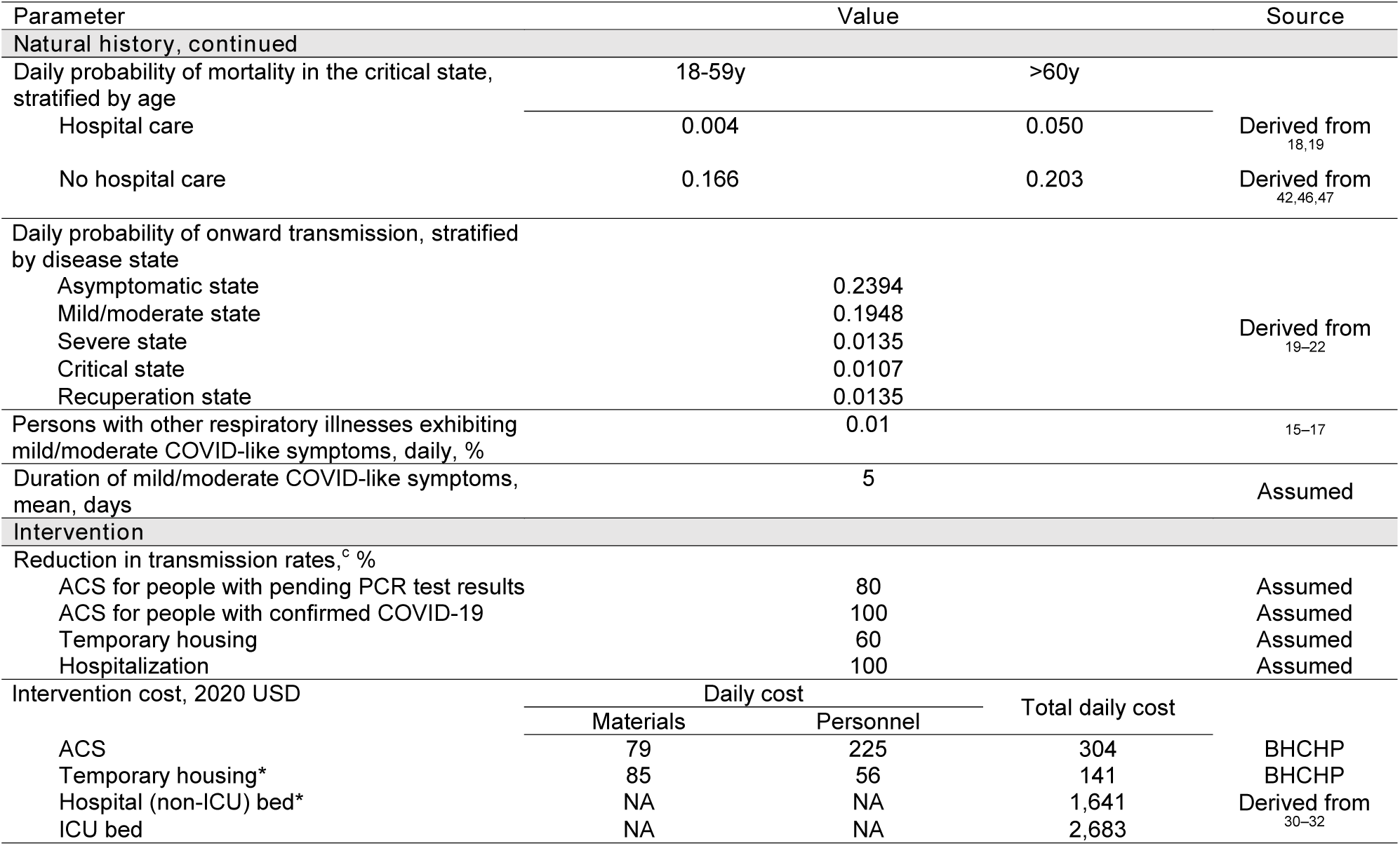
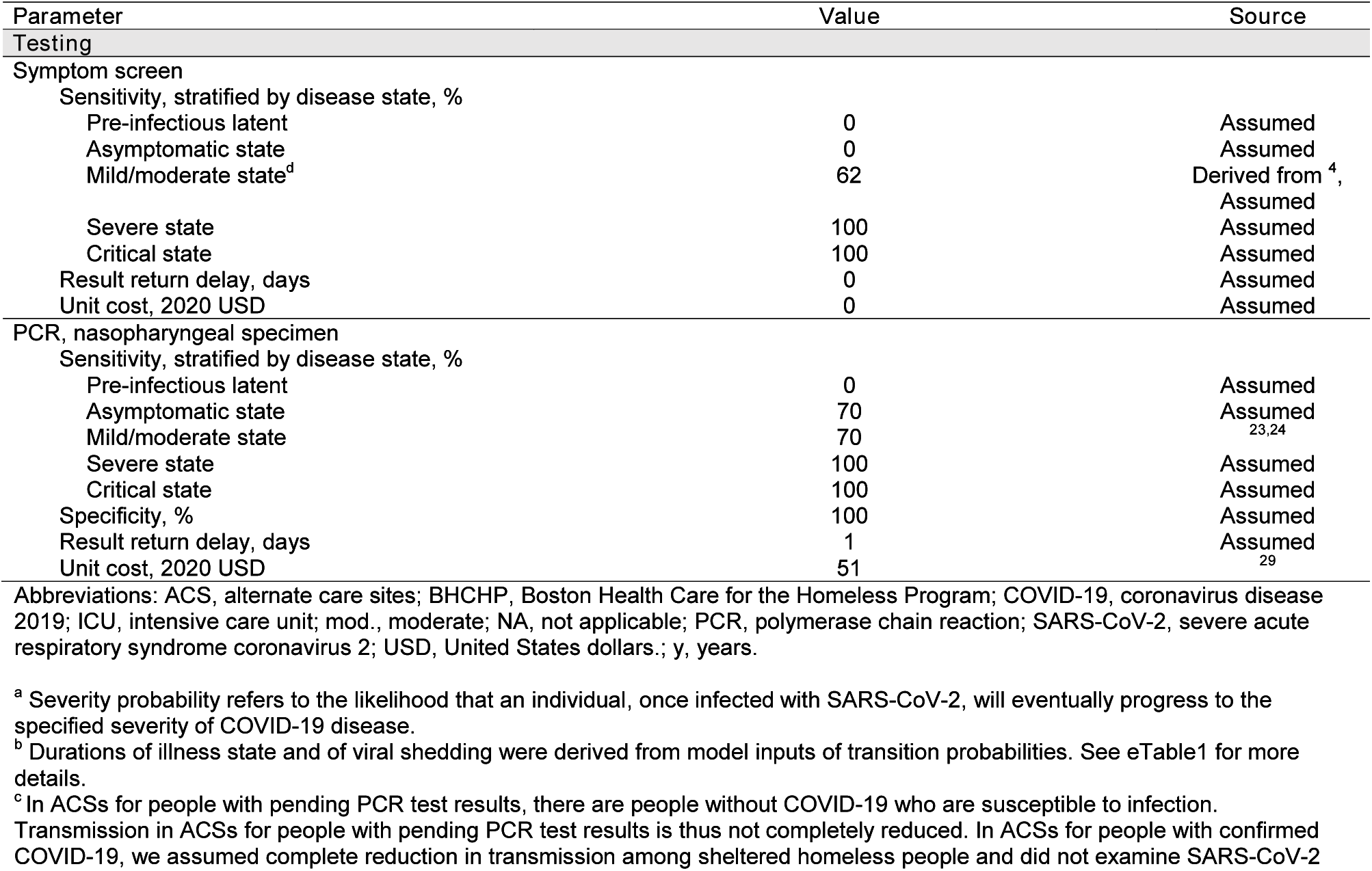
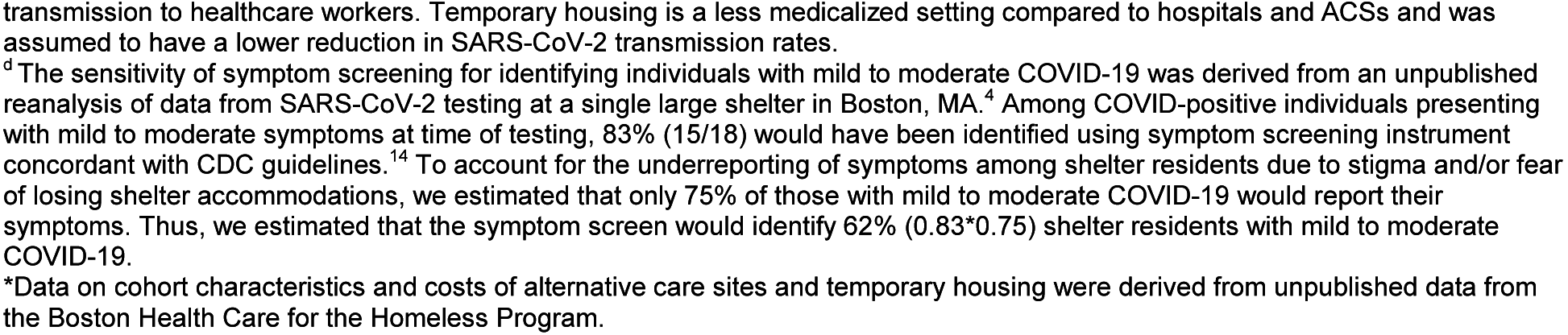
Input parameters for an analysis of management strategies for people experiencing sheltered homelessness during the COVID-19 pandemic.

**Table 2.** Results of an analysis of management strategies for people experiencing sheltered homelessness during the COVID-19 pandemic at 4 months (n=2,258).

| Strategy | Cumulative infections, n | Reduction in cases,<sup>a</sup> % | Peak daily hospital bed use, n | Total hospital days, n | Total cost,<sup>b</sup> 2020 USD | Cost compared with *NoIntervention*,<sup>b</sup> 2020 USD | Incr. cost per case prevented,<sup>b, c</sup> 2020 USD |
| --- | --- | --- | --- | --- | --- | --- | --- |
| **Effective reproduction number ($R_e$) = 2.6** |  |  |  |  |  |  |  |
| *SxScreen/PCR/ACS* | 1,239 | 36.6 | 5 | 394 | 3,267,000 | - 2,831,000 | NA |
| *Hybrid/ACS* | 985 | 49.6 | 4 | 305 | 3,628,000 | - 2,470,000 | 1,000 |
| *UniversalPCR/ACS* | 1,681 | 14.0 | 9 | 569 | 4,143,000 | - 1,955,000 | Dominated |
| *NoIntervention* | 1,954 | NA | 64 | 3,567 | 6,098,000 | NA | Dominated |
| *Hybrid/Hospital* | 967 | 50.5 | 80 | 6,796 | 12,202,000 | + 6,104,000 | Dominated |
| *SxScreen/PCR/Hospital* | 1,133 | 42.0 | 93 | 7,656 | 12,620,000 | + 6,522,000 | Dominated |
| *UniversalPCR/Hospital* | 1,679 | 14.1 | 112 | 7,165 | 12,914,000 | + 6,816,000 | Dominated |
| *UniversalPCR/TempHousing* | 376 | 80.8 | 1 | 121 | 39,119,000 | + 33,021,000 | 58,000 |
| **Effective reproduction number ($R_e$) = 1.3** |  |  |  |  |  |  |  |
| *SxScreen/PCR/ACS* | 137 | 74.5 | 1 | 48 | 409,000 | - 1,052,000 | NA |
| *Hybrid/ACS* | 103 | 80.8 | 1 | 69 | 1,325,000 | - 136,000 | 27,000 |
| *UniversalPCR/ACS* | 207 | 61.5 | 1 | 34 | 1,426,000 | - 35,000 | Dominated |
| *NoIntervention* | 538 | NA | 9 | 867 | 1,461,000 | NA | Dominated |
| *SxScreen/PCR/Hospital* | 125 | 76.7 | 22 | 966 | 1,604,000 | + 143,000 | Dominated |
| *Hybrid/Hospital* | 100 | 81.4 | 23 | 815 | 2,368,000 | + 907,000 | 382,000 |
| *UniversalPCR/Hospital* | 207 | 61.4 | 19 | 977 | 2,631,000 | + 1,170,000 | Dominated |
| *UniversalPCR/TempHousing* | 95 | 82.3 | 1 | 39 | 38,974,000 | + 37,513,000 | 6,854,000 |

**Table 2 continued.** Results of an analysis of management strategies for people experiencing sheltered homelessness during the COVID-19 pandemic at 4 months (n=2,258).
| Strategy | Cumulative infections, n | Reduction in cases,<sup>a</sup> % | Peak daily hospital bed use, n | Total hospital days, n | Total cost,<sup>b</sup> 2020 USD | Cost compared with *NoIntervention*,<sup>b</sup> 2020 USD | Incr. cost per case prevented,<sup>b, c</sup> 2020 USD |
| --- | --- | --- | --- | --- | --- | --- | --- |
| **Effective reproduction number ($R_e$) = 0.9** |  |  |  |  |  |  |  |
| *SxScreen/PCR/ACS* | 85 | 51.2 | 1 | 30 | 264,000 | - 276,000 | NA |
| *NoIntervention* | 174 | 0.0 | 5 | 318 | 540,000 | NA | Dominated |
| *SxScreen/PCR/Hospital* | 82 | 53.2 | 20 | 669 | 1,113,000 | + 573,000 | Dominated |
| *UniversalPCR/ACS* | 94 | 45.7 | 1 | 31 | 1,226,000 | + 686,000 | Dominated |
| *Hybrid/ACS* | 71 | 59.1 | 1 | 25 | 1,240,000 | + 700,000 | 71,000 |
| *UniversalPCR/Hospital* | 95 | 45.5 | 19 | 534 | 1,901,000 | + 1,361,000 | Dominated |
| *Hybrid/Hospital* | 71 | 59.4 | 22 | 595 | 2,004,000 | + 1,464,000 | Dominated |
| *UniversalPCR/TempHousing* | 71 | 59.2 | 1 | 29 | 38,954,000 | + 38,414,000 | Dominated |

Abbreviations: ACS, alternate care site; COVID-19, coronavirus disease 2019; Dominated, less clinically effective and more costly than an alternative strategy, or a combination of two alternative strategies;<sup>48</sup> Incr., incremental; NA, not applicable; *PCR*, polymerase chain reaction; *UniversalPCR*, universal polymerase chain reaction test for everyone; USD, United States dollars; *SxScreen*, symptom screen; *TempHousing*, temporary housing.
<sup>a</sup> Reduction in cases are calculated by dividing the number of cases prevented with the use of an alternative strategy by the number of cumulative cases for *NoIntervention*.
<sup>b</sup> All costs are rounded to the nearest thousands.
<sup>c</sup> Incremental costs per case prevented are calculated by dividing the difference in total costs by the difference in cumulative infections compared to the next most expensive strategy. All strategies are listed in order of ascending total costs, per convention of cost-effectiveness analysis.
Using 9.50 years of life lost per COVID-19 death from the model, and a mean age-stratified utility of 0.85 for the modeled population,<sup>42,49–51</sup> a cost per case prevented of \$1,000 is equivalent to an incremental cost-effectiveness ratio (ICER) of \$61,000/quality-adjusted life year (QALY) gained. A ratio of \$27,000/case prevented is equivalent to \$1,728,000/QALY gained. Any higher cost per case prevented has an even higher ICER.

### Strategies

We assessed 8 strategies:

1. *NoIntervention*: Only basic infection control practices are implemented in shelters.
2. *SxScreen/PCR/Hospital*: CDC-recommended symptom screening daily in shelters.^14^ Screen-negative individuals remain in shelters. Screen-positive individuals are sent to the hospital for PCR testing. PCR-positive individuals remain in hospital; PCR-negative individuals return to shelter.
3. *SxScreen/PCR/ACS*: CDC-recommended symptom screening daily in shelters. Screen-negative individuals remain in shelters. Screen-positive individuals are sent to an ACS for people under investigation, where they undergo PCR testing and await results. PCR-positive individuals with mild/moderate illness are transferred to ACSs for confirmed COVID-19 cases. PCR-negative individuals return to shelter.
4. *UniversalPCR/Hospital*: Universal PCR testing every 2 weeks in shelters. Those with symptoms at the time of testing await results at the hospital; individuals without symptoms await results in shelters. PCR-negative individuals return to or stay in shelters. PCR-positive individuals, regardless of illness severity, remain in or are sent to the hospital.
5. *UniversalPCR/ACS*: Universal PCR testing every 2 weeks in shelters. Those with symptoms at the time of testing are sent to an ACS for people under investigation while awaiting results; individuals without symptoms await results in shelters. PCR-negative individuals return to or stay in shelters. PCR-positive individuals with mild/moderate illness are transferred to ACSs for confirmed COVID-19 cases.
6. *UniversalPCR*/*TempHousing*: All shelter residents are pre-emptively moved to temporary housing for the duration of the 4-month period. Universal PCR testing occurs every 2 weeks. PCR-positive individuals with mild/moderate illness remain in temporary housing and are transferred to the hospital if they progress to severe or critical disease.
7. *Hybrid/Hospital*: This includes the *SxScreen/PCR/Hospital* strategy and adds shelter-based universal PCR testing every 2 weeks for those without symptoms.
8. *Hybrid/ACS*: This includes the *SxScreen/PCR/ACS* strategy and adds shelter-based universal PCR testing every 2 weeks for those without symptoms.

In all 8 strategies, people with severe or critical illness are sent to the hospital. Individuals are eligible for repeat PCR testing after 5 days since their most recent negative test. See eFigure2 for details.

### Input Parameters

#### Cohort characteristics

The simulated cohort represents 2,258 adults living in Boston homeless shelters.^2^ 83% are aged 18-59 years, and 17% are ≥60 years (Table 1). Initial prevalence of active or past COVID-19 is assumed to be 2.2%. To reflect symptoms similar to but not due to COVID-19 (e.g., from other respiratory viruses or seasonal rhinitis), susceptible and recovered individuals have a 0.01% daily probability of exhibiting mild/moderate COVID-like symptoms.^15–17^

#### Progression of COVID-19 and transmission

Average duration of each COVID-19 state varies by severity (eTable1). The probabilities of developing severe or critical disease or dying increase with age.^18,19^ Transmission rates are highest for individuals in asymptomatic and mild/moderate states; individuals in severe and critical states have fewer infectious contacts due to hospitalization.^19–22^

#### Testing

We assumed symptom screen sensitivity of 0% for asymptomatic infection, 62% for mild/moderate COVID-19, and 100% for severe or critical COVID-19.^4^ The PCR test is a nasopharyngeal sample with one-day result delay, 70% sensitivity for people with no symptoms or mild/moderate symptoms,^23,24^ 100% sensitivity for severe or critical illness, and 100% specificity.

#### Hospitalization, alternate care sites, and temporary housing

Mortality was decreased with hospitalization among those with critical illness.^18,19^ We assumed hospitalization reduces transmission by 100%, while ACSs reduce transmission by 80% and temporary housing by 60%. Temporary housing was assumed less effective at reducing transmission compared to ACSs due to less stringent infection control measures in temporary housing and potential mixing of uninfected and infected individuals. Length-of-stay at hospitals and ACSs depends on severity and duration of illness.^18–21,25–28^

#### Resource use and costs

The nasopharyngeal PCR test costs $51.^29^ Hospitalization costs $1,641/day; ICU costs $2,683/day (Table 1; Supplement).^30–32^ ACSs cost $304/day; temporary housing costs $141/day (data from BHCHP).

### Sensitivity Analyses

In one-way sensitivity analyses, we examined: 1) PCR sensitivity, PCR frequency, and symptom screen sensitivity (eTables2-4); 2) efficacy of ACS and temporary housing in reducing transmission (eTables5-6); and 3) costs of PCR test, symptom screen, hospital care, ACS, and temporary housing (eTables7-11). In two-way sensitivity analyses, we varied influential parameters simultaneously (eTables12-13). To relate these findings to other settings, eTable14 displays outcomes per 1,000 homeless adults and the number of sheltered homeless adults in select US cities.

## RESULTS

### Base Case

#### Surging epidemic (R_e_=2.6)

With R_e_=2.6, the number of projected COVID-19 cases was highest with *NoIntervention* (1,954) and lowest with *UniversalPCR/TempHousing* (376) (Table 2; Figure 1). Other than the temporary housing strategy, strategies that rely on daily symptom screening were more effective in preventing infections (1,133 to 1,239 cumulative infections) than those with universal PCR testing every two weeks alone (1,679 to 1,681 cumulative infections). Hybrid strategies involving daily symptom screening plus universal PCR testing every two weeks performed better than either alone (967 to 985 cumulative infections).

**Figure 1.**
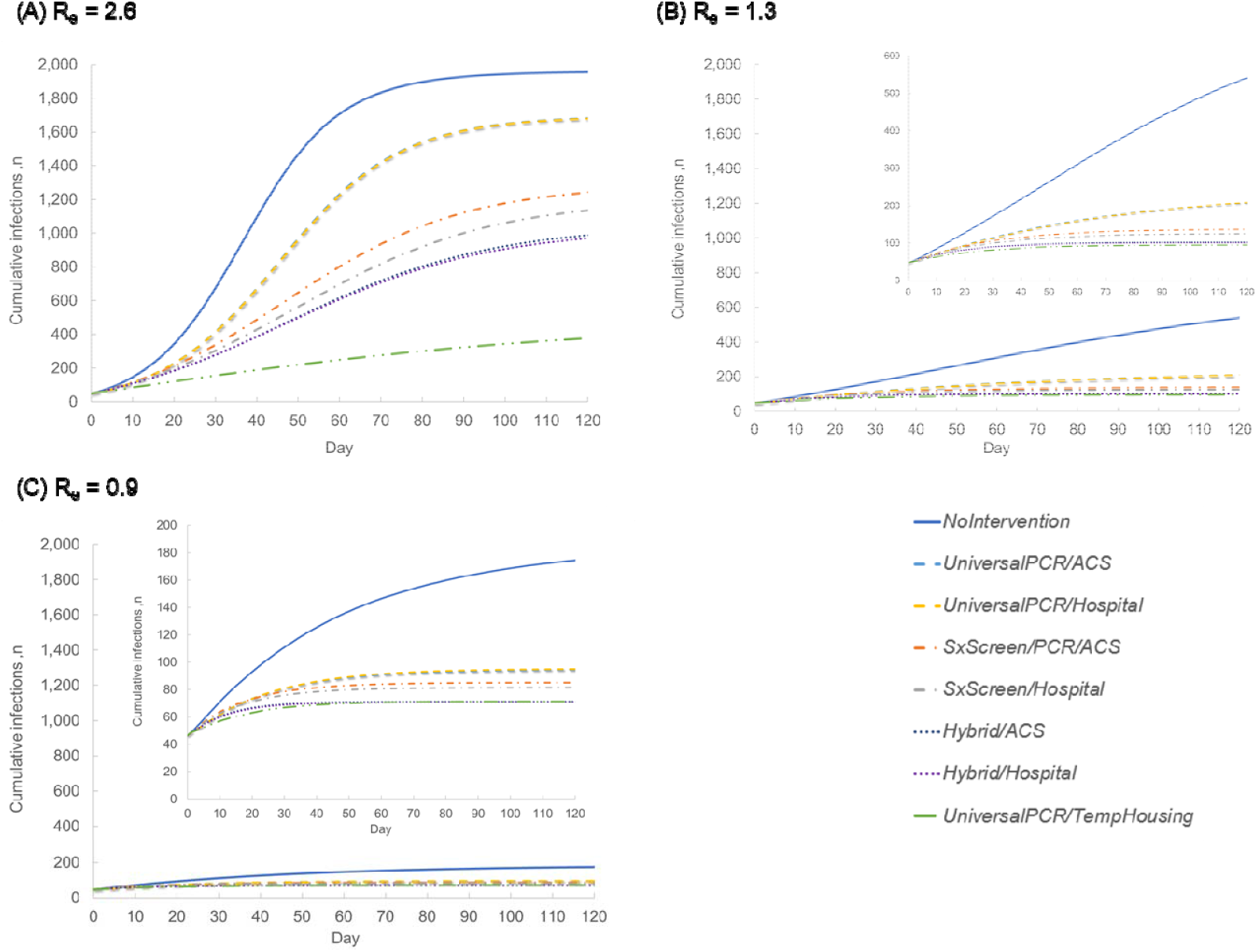
Cumulative infections by management strategy for people experiencing sheltered homelessness in Boston during the COVID-19 pandemic over a 4-month period. These panels depict the projected number of cumulative infections over time by management strategy. Panels A, B, and C show model results for R_e_ of 2.6, 1.3, and 0.9, respectively. In each panel, time 0 on the horizontal axis represents the start of model simulation, with SARS-CoV-2 infection prevalence of 2.2%. *UniversalPCR/Hospital* and *UniversalPCR/ACS* are overlapping lines since they differ only in costs; they are shown separately for clarity. The same is true for *Hybrid/Hospital* and *Hybrid/ACS*. The insets in Panels B and C magnify the vertical axis for clarity. See Methods for strategy definitions. Abbreviations: ACS, alternate care site; COVID-19, coronavirus disease 2019; *PCR*, polymerase chain reaction; *UniversalPCR*, universal polymerase chain reaction test for everyone; SARS-CoV-2, severe acute respiratory syndrome coronavirus 2; *SxScreen*, symptom screen; *TempHousing*, temporary housing.

With R_e_=2.6, all ACS-based strategies had lower total costs ($3.27 to $4.14 million) than hospital-based strategies ($12.20 to $12.91 million) and *NoIntervention* ($6.10 million; Table 2; Figure 2, eTable15). *UniversalPCR/TempHousing* was most costly ($39.12 million), and *SxScreen/PCR/ACS* was least costly ($3.27 million).

**Figure 2.**
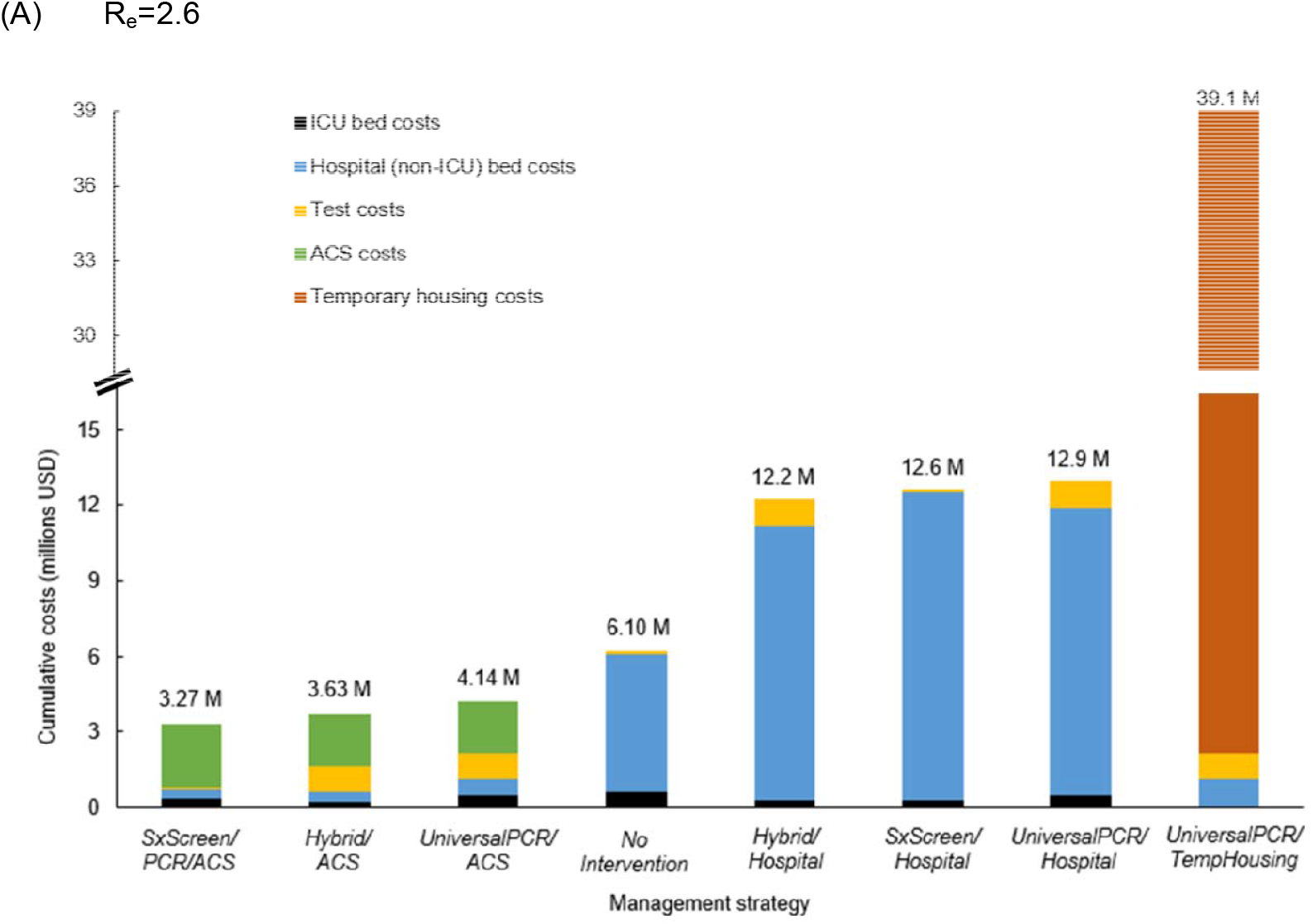
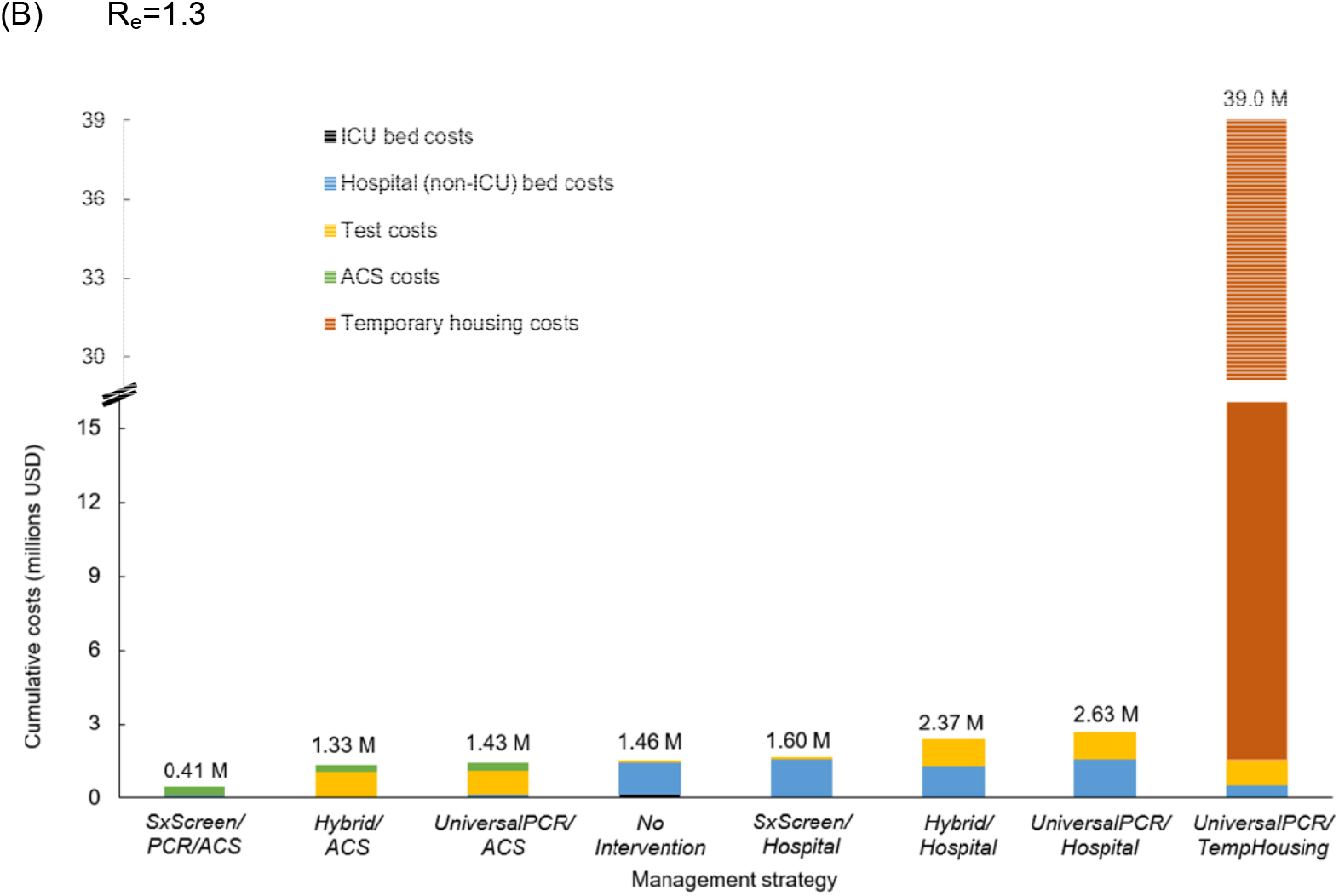
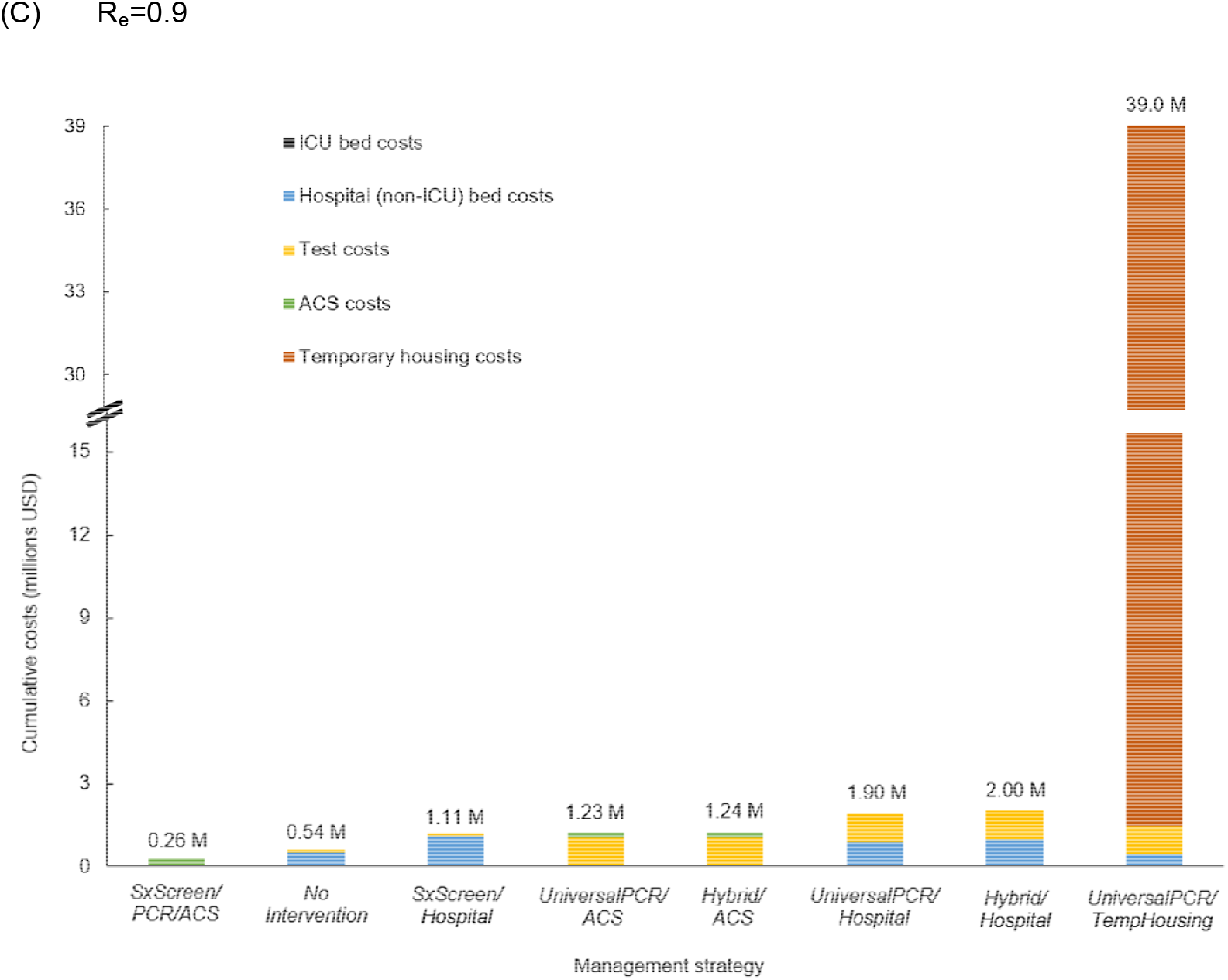
Health care sector costs of implementing different management strategies for people experiencing sheltered homelessness in Boston during the COVID-19 pandemic over a 4-month period. These panels show the total and component COVID-19-related health care costs, from a health care sector perspective, associated with different intervention strategies when applied to the adult sheltered homeless population in Boston. Panels A, B, and C show model results for R_e_ of 2.6, 1.3, and 0.9, respectively. Costs are derived from model-generated results and are undiscounted. See Methods for strategy definitions. Abbreviations: ACS, alternate care site; COVID-19, coronavirus disease 2019; ICU, intensive care unit; M, millions; *PCR*, polymerase chain reaction; *UniversalPCR*, universal polymerase chain reaction test for everyone; USD, United States dollars; *SxScreen*, symptom screen; *TempHousing*, temporary housing.

Compared with *SxScreen/PCR/ACS, Hybrid/ACS* had 20% fewer cases (985 vs 1,239) at $1,000/case prevented (Table 2). *UniversalPCR/TempHousing*, the most clinically effective strategy, had an incremental cost of $58,000/case prevented compared to *Hybrid/ACS*. All other strategies were dominated, or less effective and more costly than another strategy or combination of strategies (Table 2; Figure 2, eTable15).

#### Growing epidemic (R_e_=1.3)

With R_e_=1.3, projected cases ranged from 538 (*NoIntervention*) to 95 (*UniversalPCR/TempHousing*) (Table 2; Figure 1). All strategies had at least 60% fewer infections than *NoIntervention*. ACS strategies had fewer infections, fewer hospital days, and lower costs than *NoIntervention*, whereas hospital strategies had higher costs than *NoIntervention* (Table 2; Figure 2, eTable15). *SxScreen/PCR/ACS* had 75% fewer infections than *NoIntervention* and the lowest cost. Compared to *SxScreen/PCR/ACS, Hybrid/ACS* yielded an additional 6% decrease in infections at $27,000/case prevented. *UniversalPCR/TempHousing* had the lowest number of infections at $6,854,000/case prevented (Table 2; Figure 3).

**Figure 3.**
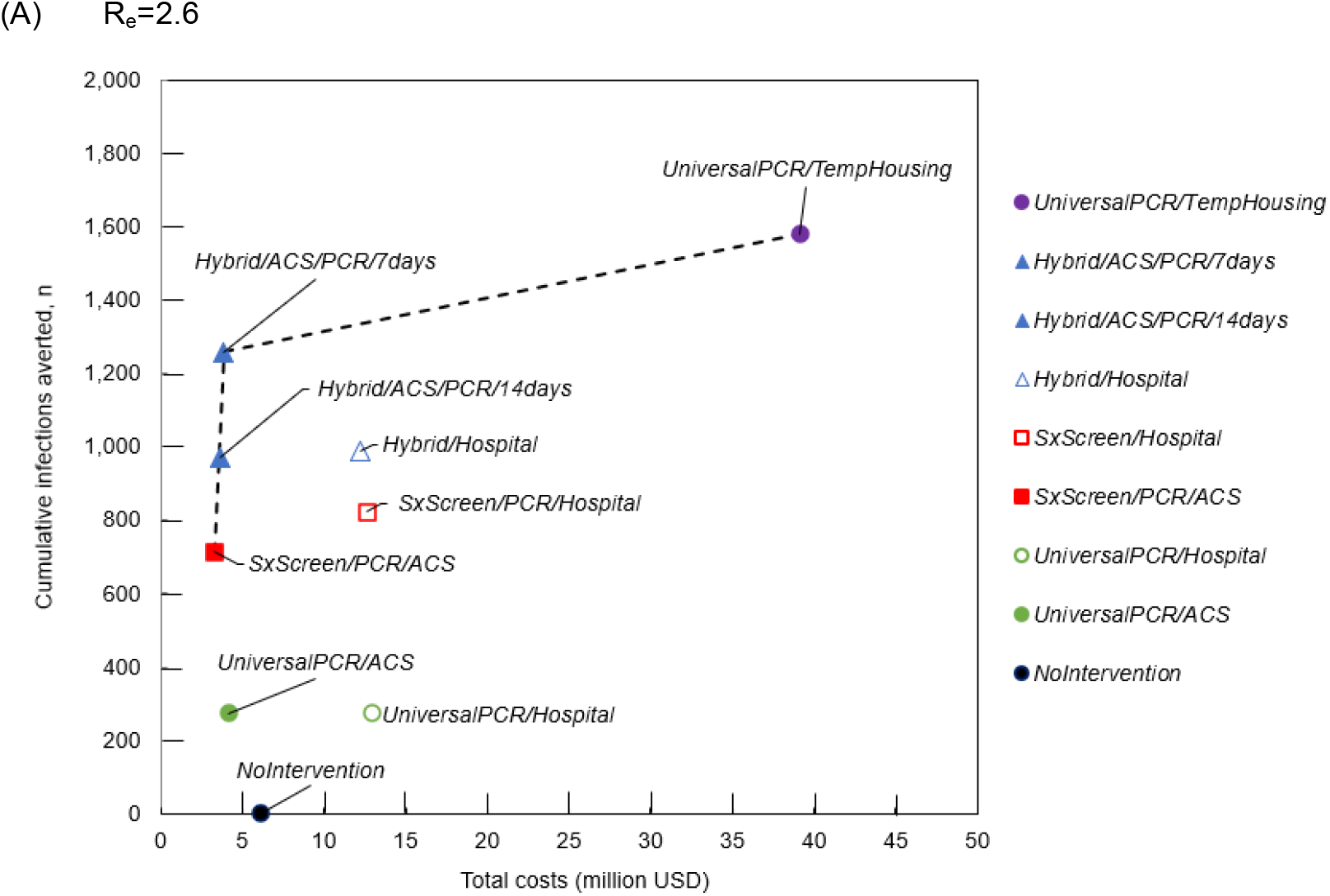
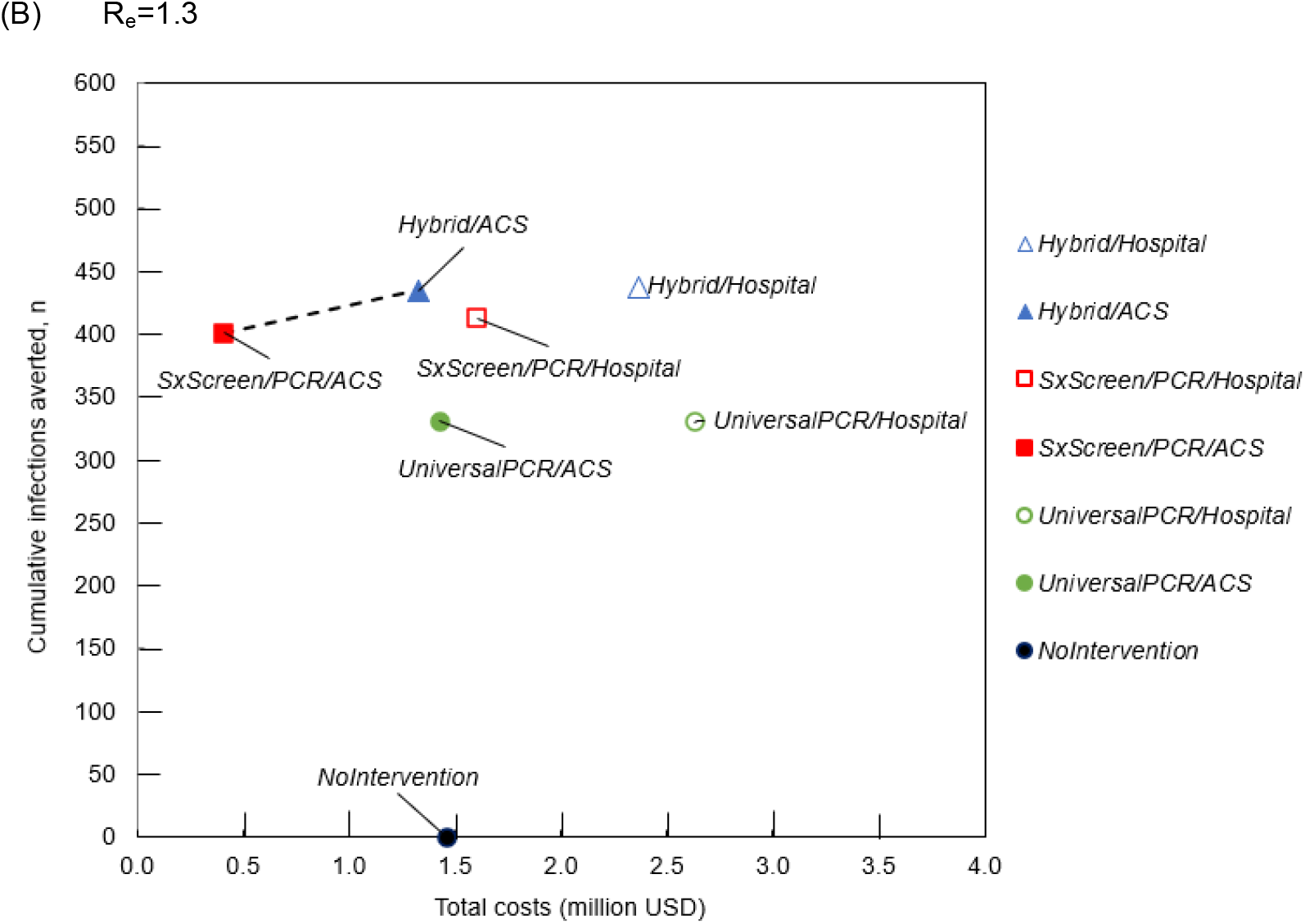
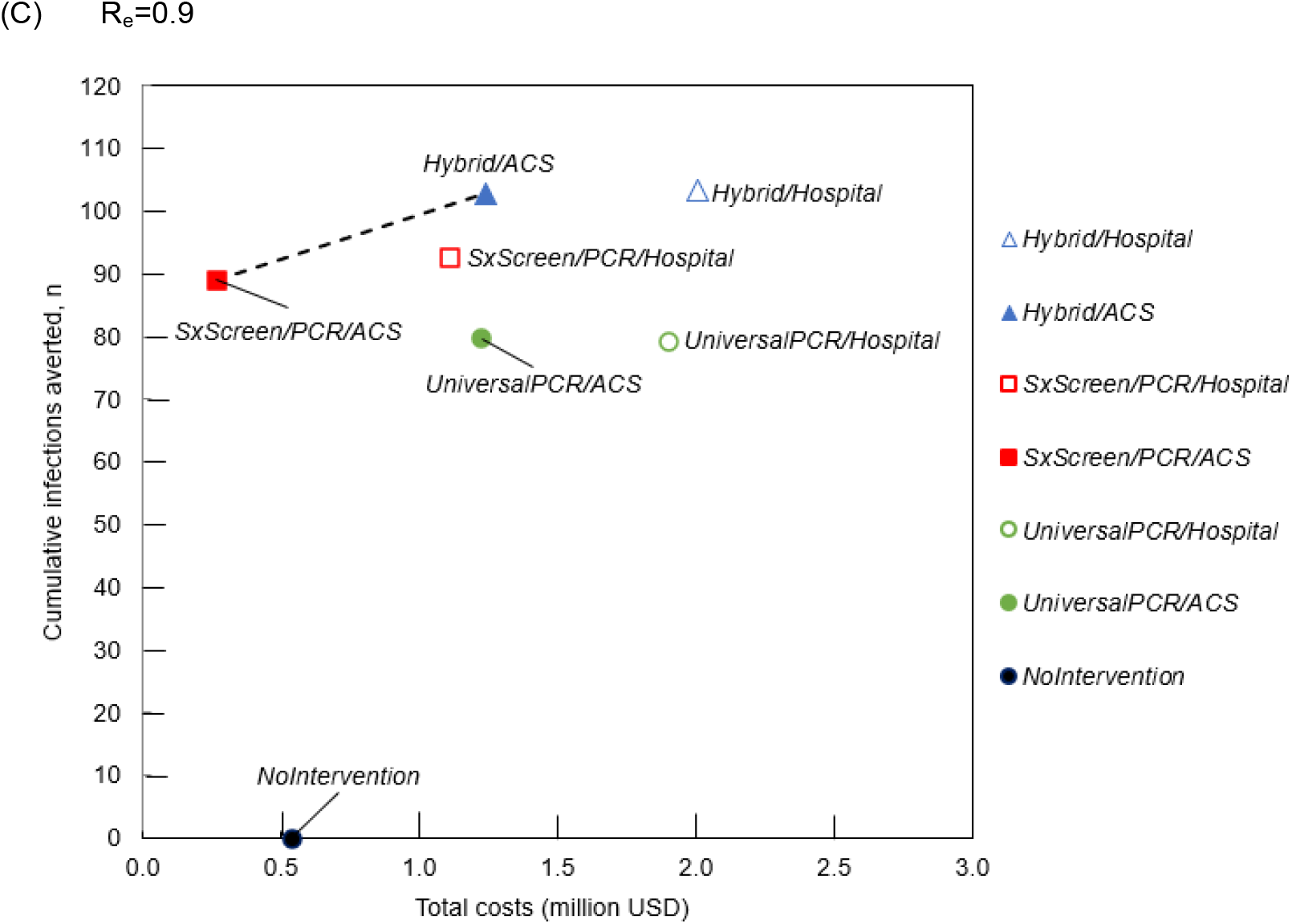
Infections averted and costs of management strategies for people experiencing sheltered homelessness in Boston during the COVID-19 pandemic over a 4-month period. Panels A, B, and C show model results for R_e_ of 2.6, 1.3, and 0.9, respectively. The circle markers represent *NoIntervention* and all strategies with universal PCR testing, which include *UniversalPCR/Hospital*, and *UniversalPCR/TempHousing*. The square markers represent strategies that are based on symptom screening, *SxScreen/PCR/Hospital* and *SxScreen/PCR/ACS*. The triangle markers represent strategies that use a combination of symptom screening and universal PCR testing, including *Hybrid/Hospital* and *Hybrid/ACS*. Additionally, the inside shading of the markers indicates the presence of ACSs for the isolation of individuals with symptoms or a positive test result. The dashed line represents the efficient frontier; strategies below this line are dominated; less clinically effective and more costly, or with a higher incremental cost per case prevented than an alternative strategy or combination of strategies. Costs are from model-generated results and are undiscounted. Results for the *UniversalPCR/TempHousing* strategy are not shown for R_e_ of 1.3 and 0.9. In addition to all base case strategies, Panel A shows the *Hybrid/ACS* strategy with PCR testing every 7 days. See Methods for strategy definitions. Abbreviations: ACS, alternate care site; COVID-19, coronavirus disease 2019; *PCR*, polymerase chain reaction; *UniversalPCR*, universal polymerase chain reaction test for everyone; USD, United States dollars; *SxScreen*, symptom screen; *TempHousing*, temporary housing.

#### Slowing epidemic (R_e_=0.9)

With R_e_=0.9, cumulative infections were fewer than in the other scenarios, ranging from 174 (*NoIntervention*) to 71 (*UniversalPCR/TempHousing*) (Table 2; Figure 1). All strategies had at least 46% fewer infections than *NoIntervention. SxScreen/PCR/ACS* had 51% fewer infections and 51% lower costs than *NoIntervention;* it was the only strategy that cost less than *NoIntervention* (Table 2; Figure 2, eTable15). Compared to *SxScreen/PCR/ACS, Hybrid/ACS* yielded an additional 8% decrease in infections at $71,000/case prevented (Table 2; Figure 3).

### Sensitivity Analyses

#### One-way sensitivity analysis

Across the 3 epidemic scenarios, changes in PCR sensitivity, PCR cost, PCR frequency, and ACS efficacy had the greatest impacts on the incremental cost per case prevented. If PCR sensitivity increased from 70% to 90% with R_e_=2.6, the number of infections with *Hybrid/ACS* decreased from 985 to 668; incremental cost per case prevented was $100 compared with *SxScreen/PCR/ACS* (eTable2). If PCR cost decreased from $51 to $25 in R_e_=2.6, the *Hybrid/ACS* strategy became cost-saving compared with *SxScreen/PCR/ACS* (eTable7). Results for higher PCR costs are also shown in eTable7. If ACS efficacy in preventing transmissions decreased, total cases increased in all ACS-based strategies, and *Hybrid/ACS* became relatively less effective compared to *SxScreen/PCR/ACS* (eTable5).

With R_e_=2.6, *Hybrid/ACS* with universal PCR testing every 7 rather than every 14 days was associated with 29% fewer infections (incremental cost of $1,000/case prevented compared with testing every 14 days, eTable16). Every 3-day testing had fewer infections, at $2,000/case prevented. In other R_e_ scenarios, the *Hybrid/ACS* strategy did not result in a cost per case prevented below $20,000 compared with *SxScreen/PCR/ACS*, regardless of universal testing frequency.

ACS-based management approaches remained less costly than hospital care unless daily ACS costs began to approach hospital costs. Although *UniversalPCR/TempHousing* had the lowest number of cases in all scenarios, with R_e_=2.6, daily costs of temporary housing needed to be ≤$20/day to have an incremental cost per case prevented of ≤$1,000 compared to *Hybrid/ACS* (eTable11). In the lower R_e_ scenarios, *UniversalPCR/TempHousing* had higher costs per case prevented.

#### Two-way sensitivity analysis

In two-way sensitivity analysis there were several combinations where *Hybrid/ACS* was cost-saving or had an incremental cost per case prevented compared to *SxScreen/PCR/ACS* of $1,000-$3,000 as the sensitivity of PCR increased and PCR cost decreased (eTable12).

## DISCUSSION

We developed a microsimulation model to examine the impact of COVID-19 testing and isolation strategies on infections and health care costs among adults experiencing sheltered homelessness. Across all epidemic scenarios, daily symptom screening with PCR testing of screen-positive individuals and ACS-based COVID-19 management was the most efficient strategy and was cost-saving relative to no intervention.

In all cases, strategies employing ACSs for isolation of symptomatic individuals with pending tests, and for those with confirmed mild or moderate COVID-19, were associated with substantially decreased costs compared to analogous strategies relying on hospital-based care while achieving similar clinical outcomes. ACSs are especially useful for managing COVID-19 in homeless populations since people with mild to moderate illness cannot be effectively isolated in shelters. With high levels of SARS-CoV-2 infection among people experiencing homelessness in Boston and other cities,^4–7,33^ ACSs could avert many hospitalizations, preserving beds for severely ill individuals and reducing costs. Boston created several such ACSs, ranging from 16-bed tents to a 500-bed field unit in a downtown convention center.^34^ In cities with smaller numbers of sheltered homeless adults (eTable14), using existing facilities (e.g. hotels/motels) as ACSs would avoid the fixed costs of new ACSs and allow for rapid implementation of care sites for people with mild to moderate COVID-19.

In a surging epidemic, adding universal PCR testing every 14 days to daily symptom screening had clinical benefits at an incremental cost of $1,000/case prevented. We selected a 2-week testing interval because this was deemed by BHCHP clinical staff to be realistic and in line with practice during the study time period; however, reducing the universal testing interval to every 7 days yielded additional benefits at $1,000/case prevented. In sensitivity analyses, this “hybrid” approach of daily symptom screening with additional periodic universal PCR testing was less expensive than daily symptom screening alone when PCR sensitivity increased and PCR cost decreased. In a growing or slowing epidemic, testing beyond daily symptom screening prevented a small number of new cases at high incremental costs. If PCR turnaround time was longer than the 1 day we modeled, all strategies would have more cases and higher costs.

Temporary housing with universal PCR testing every 2 weeks was the most effective strategy for reducing COVID-19 in all scenarios but was also the most costly, except in sensitivity analyses where temporary housing costs were reduced below plausible ranges. However, this analysis does not account for other potential benefits of temporary housing on physical or mental health.^35^ Ultimately, broader policies around supportive housing measures for people experiencing homelessness should account for more than COVID-19 mitigation, recognizing that the COVID-19 pandemic is one of many health risks of homelessness.^36^

This study complements the findings of a dynamic transition model of structural interventions for COVID-19 among people experiencing homelessness in England.^37^ In that analysis, single-room accommodations for people with COVID-19 symptoms and people without symptoms but at high risk for COVID-19 complications were projected to reduce infections, hospitalizations, and deaths by 36% to 64%. Our analysis adds to this by examining additional structural interventions (e.g. ACSs and temporary housing) in a US context, combined with various COVID-19 diagnostic approaches (e.g. symptom screening, universal PCR testing, and hybrid strategies), and by adding cost-effectiveness to inform policy and practice.

This analysis has limitations. The findings are specific to individual adults; we excluded adults experiencing homelessness as part of a family, because family shelters more likely provide private living quarters.^38^ We also excluded unsheltered homeless individuals because disease transmission dynamics and infection control considerations are distinct for this subpopulation.^39^ We assumed homogeneous mixing of sheltered homeless adults; in reality this population is spread over numerous shelters. This homogenous mixing assumption may impact the number of infections projected by our model, but we expect this impact to be small. In the base case, we did not assume increased comorbidities among homeless adults compared with the general population.^40^ The analysis is based on the possibility that ACSs and PCR tests can be made available relatively quickly to homeless adults. This may be difficult in some settings because those responsible for making ACSs and PCR tests available may not be those responsible for hospital costs, and record-keeping may be challenging. Finally, we focused this analysis on Boston, which has a 29.7% higher cost of living than the US average.^41^ Costs of temporary housing may be considerably lower in other cities. In sensitivity analysis, however, results were robust to even large changes in testing, hospital, and housing costs.

In summary, daily symptom screening and use of ACSs for those with pending test results or mild to moderate COVID-19 was associated with reduced infections and lower costs compared to no intervention. In a surging epidemic, adding universal PCR testing every 2 weeks was associated with further reduction in infections at a reasonable cost. Routine symptom screening, implementation of ACSs, and selective use of universal PCR testing should be implemented for sheltered homeless populations in the US.

## Data Availability

Data available upon request.

## ACKNOWLEDGEMENTS

We thank Elizabeth Lewis, MBA and Agnes Leung, MHA for their assistance with clinical and cost data from Boston Health Care for the Homeless Program. We also thank Guner Ege Eskibozkurt, BA and Mary Feser, BA for research assistance. All acknowledged individuals contributed as part of their institutional roles.

KAF and TPB had full access to all the data in the study and take responsibility for the integrity of the data and the accuracy of the data analysis.

## AUTHOR ROLES

All authors contributed substantively to this manuscript in the following ways: study and model design (all authors), data analysis (MHL, FMS, EL), interpretation of results (all authors), drafting the manuscript (KAF, TPB), and critical revision of the manuscript (all authors) and final approval of submitted version (all authors).

## REFERENCES

1. Henry M, Bishop K, de Sousa T, Shivji A, Watt R. The 2017 Annual Homeless Assessment Report (AHAR) to Congress PART 2: Estimates of Homelessness in the United States. The U.S. Department of Housing and Urban Development; 2018.

2. Henry M, Watt R, Mahathey A, Ouellette J, Sitler A. The 2019 Annual Homeless Assessment Report (AHAR) to Congress, Part 1: Point-in-Time Estimates of Homelessness. The U.S. Department of Housing and Urban Development; 2020.

3. CDC. Interim guidance for homeless service providers to plan and respond to coronavirus disease 2019 (COVID-19). Centers for Disease Control and Prevention. Published February 11, 2020. Accessed July 29, 2020. https://www.cdc.gov/coronavirus/2019-ncov/community/homeless-shelters/plan-prepare-respond.html

4. Baggett TP, Keyes H, Sporn N, Gaeta JM. Prevalence of SARS-CoV-2 infection in residents of a large homeless shelter in Boston. JAMA. Published online April 27, 2020. doi:10.1001/jama.2020.6887

5. Testing at Worcester homeless shelter finds 43% positive for coronavirus. WBUR. Published May 20, 2020. Accessed July 29, 2020. https://www.wbur.org/news/2020/04/17/worcester-homeless-population-covid-19-coronavirus

6. Tobolowsky FA, Gonzales E, Self JL, et al. COVID-19 outbreak among three affiliated homeless service sites - King County, Washington, 2020. MMWR Morb Mortal Wkly Rep. 2020;69(17):523–526. doi:10.15585/mmwr.mm6917e2

7. Mosites E, Parker EM, Clarke KEN. Assessment of SARS-CoV-2 infection prevalence in homeless shelters — Four U.S. cities, March 27–April 15, 2020. MMWR Morb Mortal Wkly Rep. 2020;69. doi:10.15585/mmwr.mm6917e1

8. Johns Hopkins University. COVID-19 United States cases by county. Published 2020. Accessed July 29, 2020. https://coronavirus.jhu.edu/us-map

9. Martcheva M (Maia). An Introduction to Mathematical Epidemiology. 1st ed. 2015. Springer US□: Imprint: Springer; 2015.

10. Chandrashekar A, Liu J, Martinot AJ, et al. SARS-CoV-2 infection protects against rechallenge in rhesus macaques. Science. Published online May 20, 2020:eabc4776. doi:10.1126/science.abc4776

11. CDC. Considerations for alternate care sites. Centers for Disease Control and Prevention. Published February 11, 2020. Accessed July 29, 2020. https://www.cdc.gov/coronavirus/2019-ncov/hcp/alternative-care-sites.html

12. MacKenzie OW, Trimbur MC, Vanjani R. An Isolation Hotel for People Experiencing Homelessness. N Engl J Med. 2020;383(6):e41. doi:10.1056/NEJMc2022860

13. Gold M, Siegel J, Russell L, Weinstein MC. Cost-Effectiveness in Health and Medicine. Oxford University Press; 1996.

14. CDC. Screening clients for COVID-19 at homeless shelters or encampments. Centers for Disease Control and Prevention. Published May 20, 2020. Accessed July 29, 2020. https://www.cdc.gov/coronavirus/2019-ncov/community/homeless-shelters/screening-clients-respiratory-infection-symptoms.html

15. Rui P, Okeyode T. National ambulatory medical care survey: 2016 national summary tables. Centers for Disease Control and Prevention. Published 2019. Accessed August 4, 2020. https://www.cdc.gov/nchs/data/ahcd/namcs_summary/2016_namcs_web_tables.pdf

16. Centers for Disease Control and Prevention. Percentage of visits for ILI by age group reported by a subset of ILINet providers. Published July 24, 2020. Accessed August 4, 2020. https://www.cdc.gov/coronavirus/2019-ncov/covid-data/covidview/07242020/percent-ili-visits-age.html

17. CDC. U.S. Outpatient Influenza-like Illness Surveillance Network (ILINet): Overall percentage of visits for ILI. Centers for Disease Control and Prevention. Published June 5, 2020. Accessed September 25, 2020. https://www.cdc.gov/coronavirus/2019-ncov/covid-data/covidview/06052020/percent-ili-visits.html

18. Wang D, Hu B, Hu C, et al. Clinical characteristics of 138 hospitalized patients with 2019 novel coronavirus–infected pneumonia in Wuhan, China. JAMA. 2020;323(11):1061–1069. doi:10.1001/jama.2020.1585

19. Zhou F, Yu T, Du R, Fan G, Liu Y, Liu Z. Clinical course and risk factors for mortality of adult inpatients with COVID-19 in Wuhan, China: a retrospective cohort study. The Lancet. 2020;395(10229):1054–1062.

20. Report of the WHO-China Joint Mission on Coronavirus Disease 2019 (COVID-19). WHO-China Joint Mission on Coronavirus Disease 2019 (COVID-19); 2020. Accessed July 29, 2020. https://www.who.int/docs/default-source/coronaviruse/who-china-joint-mission-on-covid-19-final-report.pdf

21. Hu Z, Song C, Xu C, et al. Clinical characteristics of 24 asymptomatic infections with COVID-19 screened among close contacts in Nanjing, China. medRxiv. Published online January 1, 2020:2020.02.20.20025619. doi:10.1101/2020.02.20.20025619

22. Liu Y, Gayle AA, Wilder-Smith A, Rocklöv J. The reproductive number of COVID-19 is higher compared to SARS coronavirus. Journal of Travel Medicine. 2020;27(taaa021). doi:10.1093/jtm/taaa021

23. Yang Y, Yang M, Shen C, et al. Evaluating the accuracy of different respiratory specimens in the laboratory diagnosis and monitoring the viral shedding of 2019-nCoV infections. medRxiv. Published online January 1, 2020:2020.02.11.20021493. doi:10.1101/2020.02.11.20021493

24. Wang W, Xu Y, Gao R, et al. Detection of SARS-CoV-2 in different types of clinical specimens. JAMA. 2020;323(18):1843–1844. doi:10.1001/jama.2020.3786

25. Li Q, Guan X, Wu P, et al. Early transmission dynamics in Wuhan, China, of novel coronavirus–infected pneumonia. N Engl J Med. 2020;382(13):1199–1207. doi:10.1056/NEJMoa2001316

26. He X, Lau EH, Wu P, et al. Temporal dynamics in viral shedding and transmissibility of COVID-19. medRxiv. Published online January 1, 2020:2020.03.15.20036707. doi:10.1101/2020.03.15.20036707

27. Linton NM, Kobayashi T, Yang Y, et al. Incubation period and other epidemiological characteristics of 2019 novel coronavirus infections with right truncation: a statistical analysis of publicly available case data. J Clin Med. 2020;9(2):538. doi:10.3390/jcm9020538

28. Yu P, Zhu J, Zhang Z, Han Y. A familial cluster of infection associated with the 2019 novel coronavirus indicating possible person-to-person transmission during the incubation period. The Journal of Infectious Diseases. 2020;221(11):1757–1761. doi:10.1093/infdis/jiaa077

29. Medicare Administrative Contractor (MAC) COVID-19 test pricing. Published May 19, 2020. Accessed July 29, 2020. https://www.cms.gov/files/document/mac-covid-19-test-pricing.pdf

30. Cox C, Rudowitz R, Neuman T, Cubanski J, Rae M. How health costs might change with COVID-19. Health System Tracker. Published April 15, 2020. Accessed July 29, 2020. https://www.healthsystemtracker.org/brief/how-health-costs-might-change-with-covid-19/

31. Rae M, Claxton G, Kurani N, McDermott D, Cox C. Potential costs of COVID-19 treatment for people with employer coverage. Peterson-Kaiser Health System Tracker. Published 2020. Accessed July 29, 2020. https://www.healthsystemtracker.org/brief/potential-costs-of-coronavirus-treatment-for-people-with-employer-coverage/

32. COVID-19: The Projected Economic Impact of the COVID-19 Pandemic on the US Healthcare System. FAIR Health, Inc.; 2020. Accessed July 29, 2020. https://s3.amazonaws.com/media2.fairhealth.org/brief/asset/COVID-19%20-%20The%20Projected%20Economic%20Impact%20of%20the%20COVID-19%20Pandemic%20on%20the%20US%20Healthcare%20System.pdf

33. Stokes S. Atlanta tests more than 2,000 people who are homeless for COVID-19. WABE. Published April 21, 2020. Accessed July 29, 2020. https://www.wabe.org/atlanta-tests-more-than-2000-people-who-are-homeless-for-covid-19/

34. Convention centers fill with beds for COVID-19, including 500 for Boston’s homeless. Accessed July 29, 2020. https://www.wbur.org/commonhealth/2020/04/09/convention-centers-transform-into-field-hospitals-in-boston-and-worcester

35. Padgett DK, Stanhope V, Henwood BF, Stefancic A. Substance use outcomes among homeless clients with serious mental illness: comparing Housing First with Treatment First programs. Community Ment Health J. 2011;47(2):227–232. doi:10.1007/s10597-009-9283-7

36. Tsai J, Wilson M. COVID-19: a potential public health problem for homeless populations. Lancet Public Health. 2020;5(4):e186–e187. doi:10.1016/S2468-2667(20)30053-0

37. Lewer D, Braithwaite I, Bullock M, Eyre MT, Aldridge RW. COVID-19 and homelessness in England: a modelling study of the COVID-19 pandemic among people experiencing homelessness, and the impact of a residential intervention to isolate vulnerable people and care for people with symptoms. medRxiv. Published online January 1, 2020:2020.05.04.20079301. doi:10.1101/2020.05.04.20079301

38. Overview of the Department of Housing and Community Development. Mass.gov. Published August 28, 2019. Accessed October 2, 2020. https://www.mass.gov/info-details/overview-of-the-department-of-housing-and-community-development#ea-program-

39. CDC. Interim guidance on unsheltered homelessness and coronavirus disease 2019 (COVID-19) for homeless service providers and local officials. Centers for Disease Control and Prevention. Published May 13, 2020. Accessed July 29, 2020. https://www.cdc.gov/coronavirus/2019-ncov/community/homeless-shelters/unsheltered-homelessness.html

40. Brown RT, Hemati K, Riley ED, et al. Geriatric conditions in a population-based sample of older homeless adults. The Gerontologist. 2016;57(4):757–766. doi:10.1093/geront/gnw011

41. Cost of living data series. Missouri Economic Research and Information Center. Published 2020. Accessed September 25, 2020. https://meric.mo.gov/data/cost-living-data-series

42. COVID-19 Dashboard. Massachusetts Department of Public Health. Published 2020. Accessed July 29, 2020. https://www.mass.gov/doc/covid-19-dashboard-april-20-2020/download

43. Mizumoto K, Kagaya K, Zarebski A, Chowell G. Estimating the asymptomatic proportion of coronavirus disease 2019 (COVID-19) cases on board the Diamond Princess cruise ship, Yokohama, Japan, 2020. Euro Surveill. 2020;25(10):2000180. doi:10.2807/1560-7917.ES.2020.25.10.2000180

44. Haridy R. CDC director warns 25 percent of COVID-19 cases may present no symptoms. New Atlas. https://newatlas.com/health-wellbeing/covid-19-cases-contagious-asymptomatic-presymptomatic-cdc-director/. Published April 1, 2020. Accessed July 29, 2020.

45. Li R, Pei S, Chen B, et al. Substantial undocumented infection facilitates the rapid dissemination of novel coronavirus (SARS-CoV2). Science. Published online March 16, 2020:eabb3221. doi:10.1126/science.abb3221

46. American community survey 1-year estimates. U.S. Census Bureau. Published 2020. Accessed July 29, 2020. https://censusreporter.org/profiles/04000US25-massachusetts/

47. Richardson S, Hirsch JS, Narasimhan M, et al. Presenting characteristics, comorbidities, and outcomes among 5700 patients hospitalized with COVID-19 in the New York City area. JAMA. Published online April 22, 2020. doi:10.1001/jama.2020.6775

48. Sanders GD, Neumann PJ, Basu A, et al. Recommendations for conduct, methodological practices, and reporting of cost-effectiveness analyses: second panel on cost-effectiveness in health and medicine. JAMA. 2016;316(10):1093–1103. doi:10.1001/jama.2016.12195

49. Sullivan PW, Ghushchyan V. Preference-Based EQ-5D index scores for chronic conditions in the United States. Med Decis Making. 2006;26(4):410–420. doi:10.1177/0272989X06290495

50. Gardner JW, Sanborn JS. Years of Potential Life Lost (YPLL) - What Does it Measure? Epidemiology. 1990;1(4).

51. Martinez R, Soliz P, Caixeta R, Ordunez P. Reflection on modern methods: years of life lost due to premature mortality-a versatile and comprehensive measure for monitoring non-communicable disease mortality. Int J Epidemiol. 2019;48(4):1367–1376. doi:10.1093/ije/dyy254

