## Supplemental Materials for "Clinical Outcomes, Costs, and Cost-effectiveness of Strategies for People Experiencing Sheltered Homelessness During the COVID-19 Pandemic"

**TABLE OF CONTENTS**

**Supplemental Methods**

**eTable1.** Additional input parameters for an analysis of management strategies for people experiencing sheltered homelessness during the COVID-19 pandemic.

**eTable2.** One-way sensitivity analysis on PCR sensitivity for people with mild/moderate illness.

**eTable3.** One-way sensitivity analysis on universal PCR testing frequency.

**eTable4.** One-way sensitivity analysis on symptom screen sensitivity for people with mild/moderate illness.

**eTable5.** One-way sensitivity analysis on efficacy of ACS for COVID-confirmed in reducing SARS-CoV-2 transmission.

**eTable6.** One-way sensitivity analysis on efficacy of temporary housing in reducing SARS-CoV-2 transmission.

**eTable7.** One-way sensitivity analysis on cost of a PCR test.

**eTable8.** One-way sensitivity analysis on cost of symptom screen.

**eTable9.** One-way sensitivity analysis on daily costs of hospital beds.

**eTable10.** One-way sensitivity analysis on daily cost of an ACS.

**eTable11.** One-way sensitivity analysis on daily cost of temporary housing.

**eTable12.** Two-way sensitivity analysis on PCR sensitivity and PCR cost for the *SxScreen/PCR/ACS* and *Hybrid/ACS* strategies.

**eTable13.** Two-way sensitivity analysis on PCR testing frequency and PCR cost for the *SxScreen/PCR/ACS* and *Hybrid/ACS* strategies.

**eTable14.** Results of an analysis of management strategies for people experiencing sheltered homelessness during the COVID-19 pandemic for a cohort of 1,000 adults experiencing sheltered homelessness.

**eTable15.** Total infections and component costs of different management strategies for adults experiencing sheltered homelessness in Boston during the COVID-19 pandemic at 4 months.

**eTable16.** One-way sensitivity analysis on PCR testing frequency for the *Hybrid/ACS* strategy.

**Legend to Figures**

**eFigure1.** Illustration of health states and illness paths in the CEACOV model.

**eFigure2.** Flow diagrams of management strategies for people experiencing sheltered homelessness in Boston during the COVID-19 pandemic.

**INTRODUCTION**

In this supplemental appendix, we provide additional model inputs, results from one-way and multi-way sensitivity analyses, and scaled results for a cohort of 1,000 adults experiencing sheltered homelessness. In addition, we provide details on the Clinical and Economic Analysis of COVID-19 interventions (CEACOV) model and management strategies for people experiencing sheltered homelessness.

**SUPPLEMENTAL METHODS**

**Health States**

CEACOV simulates individuals transitioning between the states of susceptibility to severe acute respiratory syndrome coronavirus 2 (SARS-CoV-2), infection with SARS-CoV-2 and coronavirus disease 2019 (COVID-19) illness, recovery, or death. Susceptible individuals face a daily probability of acquiring infection. After acquiring SARS-CoV-2 infection, individuals may progress through the following health states (eFigure1):

- Pre-infectious latency
- Asymptomatic infection
- Mild/moderate illness: symptomatic
- Severe illness: dyspnea and/or hypoxemia ideally managed in a hospital with standard supplemental oxygen but not requiring intensive care unit (ICU)
- Critical illness: ideally managed in an ICU with high-flow supplemental oxygen, non-invasive positive pressure ventilation, or invasive mechanical ventilation
- Recuperation: for those recuperating from critical illness and improving while remaining in the hospital or other health care facility
- Recovered

Individuals in the asymptomatic infection, mild/moderate illness, and severe illness states can transition directly to the recovered state. Individuals in the critical illness state can eventually die, or transition to the recuperation state and then to the recovered state. The recovered state is an absorbing state, and recovered individuals are assumed to have immunity to SARS-CoV-2 over the model time horizon.

**Natural History Paths**

After being infected with SARS-CoV-2, a susceptible individual first transitions to the pre-infectious latency stage. Then, the individual has an age-dependent probability of progressing along one of four “paths,” culminating in either asymptomatic infection, mild/moderate illness, severe illness, or critical illness. Before reaching a more advanced illness state, individuals first transition through intermediate states (e.g., those destined for severe illness must first pass through the asymptomatic infection state and the mild/moderate illness state, eFigure1).

**Transmission**

The basic reproduction number (R_0_) is defined as the daily rate at which an infected individual contacts susceptible individuals and infects them in a fully susceptible cohort, multiplied by the duration of infectivity:

R_0_ = K * b * D, where

- K: number of contacts per day an infected individual has with susceptible people in a fully susceptible cohort
- b: the probability of transmission per contact between an infectious individual and a susceptible person
- D: the mean duration of infectivity

In CEACOV, infected individuals in mild/moderate, severe, critical, and recuperation states can transmit SARS-CoV-2 to susceptible individuals. The effective transmission rate of infected individuals in one of these states is calculated as follows:

*Effective Transmission Rate (R_eff_) = Nominal Transmission Rate (R_nom_) * Transmission Multiplier.*

*Nominal Transmission Rate = R_0_/D.*

The Nominal Transmission Rate is defined as R_0_ in a fully susceptible cohort divided by the average duration of infectivity (D). Equivalently, it can be defined as a function of the average number of contacts per day an infected individual has with susceptible people in a fully susceptible cohort (K) multiplied by the probability of transmission per contact between an infectious individual and a susceptible person (b), therefore the contact rates and probability of transmission per contact are implicitly included in the nominal transmission rate. R_0_ estimates the expected number of secondary cases produced by an infected individual in a fully susceptible cohort. Once the epidemic is underway and a subset of the population is infected, the effective reproduction number (R_e_) measures the number of secondary cases produced by an infected individual in a progressed epidemic. This Nominal Transmission Rate captures the ratio (rather than the magnitude) of daily infectivity across different infection states (e.g., asymptomatic to mild/moderate, or mild/moderate to severe). Infected individuals do not transmit while they are in the latent state or in the recovered state. Patients in other infected states can transmit SARS-CoV-2 to susceptible individuals. The effective magnitude of the transmission rate changes over time as social interventions alter the number of contacts (K) and infectivity (b), and subsequently, the effective reproduction number (R_e_); thus, the magnitude is adjusted using the transmission multiplier.Transmission multipliers are setting-specific, time-dependent adjusting factors, roughly accounting for population density and interventions that can alter the number of contacts (e.g., social distancing) and the infectivity per contact (e.g., masking) in the setting being modeled. In this analysis, the transmission multipliers were calibrated to represent a surging, a growing, and a slowing epidemic. Given the time-dependent uptake of such policies, time-varying transmission multipliers may be used to alter the effective transmission rate (R_eff_) over time. R_eff_ determines the effective transmission rate of infected individuals on each day of simulation and is equivalent to the effective reproduction number (R_e_) divided by the duration of infectivity. Therefore, the effective transmission rate (R_eff_) in CEACOV and the effective reproduction number (R_e_) are directly related.

In this analysis, we assumed that all susceptible persons have an equal probability of contacting infected individuals and acquiring the virus (i.e., homogenous mixing). As the epidemic grows, the number of susceptible persons declines. Thus, not all of the daily contacts of infected individuals will be with susceptible persons. The daily infection rate for a susceptible person is equal to the sum of transmission rates from all infected persons across all infection states divided by the cohort size. This leads to an expected daily number of infections equal to the number of susceptible people multiplied by the infection rate on that day.

It is worth noting that the number of secondary cases each infected individual produces is defined by the effective reproduction number (R_e_). While the assumption on mixing (homogenous vs. heterogenous) affects ‘who’ an infected individual transmits to, the average number of new infections caused by a given infected individual is determined by the effective reproduction number.

Under the homogenous mixing assumption, an infected individual in Shelter A will uniformly infect susceptibles in other shelters; similarly, an infected individual in Shelter B will uniformly infect susceptibles in other shelters, including Shelter A. However, under the heterogeneous mixing assumption, an infected individual in Shelter A would most likely infect susceptibles in the same shelter, and an infected individual in Shelter B would similarly infect susceptibles in Shelter B. We assume that all shelters are roughly similar and that the assumption of homogenous mixing for this population is adequate.

**Calibration and Validation of CEACOV**

We initiated the model with a cohort of 1 million simulated persons who are meant to represent the 6.9 million population of Massachusetts (MA) in 2020. Each person’s age category was drawn at model start based on MA age distribution data.^1^ Prevalence of COVID-19 at model start was set to 0.14% to represent the approximate prevalence of COVID-19 in mid-March in MA.^2^ We tracked the number of people in each health state over a 45-day horizon.

We calibrated the transmission multiplier (see above) to the COVID-19 epidemic in MA from mid-March to mid-April (first 30 days) and used the data from the remaining 15 days to validate the model. We assumed the reported number of COVID-19-attributable deaths would be close to the actual number of deaths. Hence, the number of reported COVID-19-attributable deaths was our main calibration target. We removed 59% of deaths to account for the deaths occurring in long term care facilities (LTCF) and not in the community. To ensure good model fit, we checked the mean absolute percentage error (MAPE) and the median absolute percentage error (MEDAPE) for modeled and observed number of deaths over the validation horizon. The MAPE was 3.3%, while the MEDAPE was 3.2%.

**Resource Costs**

We did not include the cost of existing shelters, because this is not borne by the health care sector, or the cost of symptom screening, assuming this would be conducted by current shelter staff. Hospital costs were derived from IBM MarketScan Commercial Claims and Encounters database employer plans.^3,4^ All other costs were in 2020 USD and unadjusted .

To estimate daily costs for hospital beds and intensive care unit (ICU) beds, we first used the average Medicare-allowed inpatient coverage of five diagnoses related to pneumonia and ventilator use as well as the average Medicare-allowed professional cost to obtain the total hospitalization costs. Subsequently, we divided the total costs by the average number of days that a patient with pneumonia spent in the hospital or the ICU to obtain the average daily costs for hospital beds and ICU beds, respectively.

More specifically, we used the average Medicare-allowed coverage across 3 diagnosis-related groups (DRG): DRG 193 “simple pneumonia and pleurisy with major complications”, DRG 194 “pneumonia with complications or comorbidity,” and DRG 195 “pneumonia without complications” to estimate the average Medicare-allowed inpatient coverage of pneumonia patients. In addition, we used DRG 207 “respiratory system diagnosis with ventilator support required for 96 hours or more”, and DRG 208 “respiratory system diagnosis with ventilator support required for less than 96 hours” to estimate the average daily ICU cost. An average Medicare-allowed professional cost of $395 was added to the average allowed Medicare inpatient daily cost of hospitalization and ICU to obtain final total daily costs of hospitalization and ICU.^3–5^

**CDC Symptom Screen Recommendations**

The CDC recommends daily symptom screenings for COVID-19 at homeless shelters.^6^ The following list of symptoms suggests the possibility of mild to moderate illness: fever, worsening cough, shortness of breath or difficulty breathing, fatigue, muscle or body aches, headache, new loss of taste or smell, sore throat, congestion or runny nose, nausea or vomiting, and diarrhea. More severe symptoms include trouble breathing, persistent pain or pressure in the chest, new confusion, inability to wake or stay awake, and bluish lips or face.^6^

**APPENDIX REFERENCES**

1.	Census profile: Massachusetts. Census Reporter. Accessed July 29, 2020. http://censusreporter.org/profiles/04000US25-massachusetts/

2.	Archive of COVID-19 cases in Massachusetts. Massachusetts Department of Public Health. Accessed July 29, 2020. https://www.mass.gov/info-details/archive-of-covid-19-cases-in-massachusetts

3.	Rae M, Claxton G, Kurani N, McDermott D, Cox C. Potential costs of COVID-19 treatment for people with employer coverage. Peterson-Kaiser Health System Tracker. Published 2020. Accessed July 29, 2020. https://www.healthsystemtracker.org/brief/potential-costs-of-coronavirus-treatment-for-people-with-employer-coverage/

4.	Cox C, Rudowitz R, Neuman T, Cubanski J, Rae M. How health costs might change with COVID-19. Health System Tracker. Published April 15, 2020. Accessed July 29, 2020. https://www.healthsystemtracker.org/brief/how-health-costs-might-change-with-covid-19/

5.	*COVID-19: The Projected Economic Impact of the COVID-19 Pandemic on the US Healthcare System*. FAIR Health, Inc.; 2020. Accessed July 29, 2020. https://s3.amazonaws.com/media2.fairhealth.org/brief/asset/COVID-19%20-%20The%20Projected%20Economic%20Impact%20of%20the%20COVID-19%20Pandemic%20on%20the%20US%20Healthcare%20System.pdf

6.	CDC. Screening clients for COVID-19 at homeless shelters or encampments. Centers for Disease Control and Prevention. Published May 20, 2020. Accessed July 29, 2020. https://www.cdc.gov/coronavirus/2019-ncov/community/homeless-shelters/screening-clients-respiratory-infection-symptoms.html

7.	He X, Lau EH, Wu P, et al. Temporal dynamics in viral shedding and transmissibility of COVID-19. *medRxiv*. Published online January 1, 2020:2020.03.15.20036707. doi:10.1101/2020.03.15.20036707

8.	Linton NM, Kobayashi T, Yang Y, et al. Incubation period and other epidemiological characteristics of 2019 novel coronavirus infections with right truncation: a statistical analysis of publicly available case data. *J Clin Med*. 2020;9(2):538. doi:10.3390/jcm9020538

9.	*Report of the WHO-China Joint Mission on Coronavirus Disease 2019 (COVID-19)*. WHO-China Joint Mission on Coronavirus Disease 2019 (COVID-19); 2020. Accessed July 29, 2020. https://www.who.int/docs/default-source/coronaviruse/who-china-joint-mission-on-covid-19-final-report.pdf

10.	Li R, Pei S, Chen B, et al. Substantial undocumented infection facilitates the rapid dissemination of novel coronavirus (SARS-CoV2). *Science*. Published online March 16, 2020:eabb3221. doi:10.1126/science.abb3221

11.	Wang D, Hu B, Hu C, et al. Clinical characteristics of 138 hospitalized patients with 2019 novel coronavirus–infected pneumonia in Wuhan, China. *JAMA*. 2020;323(11):1061-1069. doi:10.1001/jama.2020.1585

12.	Zhou F, Yu T, Du R, Fan G, Liu Y, Liu Z. Clinical course and risk factors for mortality of adult inpatients with COVID-19 in Wuhan, China: a retrospective cohort study. *The Lancet*. 2020;395(10229):1054-1062.

13.	Hu Z, Song C, Xu C, et al. Clinical characteristics of 24 asymptomatic infections with COVID-19 screened among close contacts in Nanjing, China. *medRxiv*. Published online January 1, 2020:2020.02.20.20025619. doi:10.1101/2020.02.20.20025619

14.	Sanders GD, Neumann PJ, Basu A, et al. Recommendations for conduct, methodological practices, and reporting of cost-effectiveness analyses: second panel on cost-effectiveness in health and medicine. *JAMA*. 2016;316(10):1093-1103. doi:10.1001/jama.2016.12195

15.	Henry M, Watt R, Mahathey A, Ouellette J, Sitler A. *The 2019 Annual Homeless Assessment Report (AHAR) to Congress, Part 1: Point-in-Time Estimates of Homelessness*. The U.S. Department of Housing and Urban Development; 2020.

**eTable1. Additional input parameters for an analysis of management strategies for people experiencing sheltered homelessness during the COVID-19 pandemic.**

| **Parameter** | **Value** | | | | **Source** | |
| --- | --- | --- | --- | --- | --- | --- |
| **Natural history** |  | | | |  | |
| Initial illness distribution, % |  | | | | Model calibration* | |
| Susceptible | 97.83 | | | |  | |
| Infected |  | | | |  | |
| Pre-infectious latent | 1.07 | | | |  | |
| Asymptomatic | 0.56 | | | |  | |
| Mild/Moderate | 0.35 | | | |  | |
| Severe | 0.02 | | | |  | |
| Critical | 0.00 | | | |  | |
| Recuperating | 0.00 | | | |  | |
| Recovered | 0.17 | | | |  | |
| Daily probability of transition between disease states, stratified by COVID-19 severity, among hospitalized | Asymptomatic infection | Mild/moderate illness | Severe illness | Critical illness | |  |
| Pre-infectious latency to asymptomatic state | 0.323 | 0.323 | 0.323 | 0.323 | | Derived from ^7–10^ |
| Asymptomatic to mild/moderate state | NA | 0.394 | 0.394 | 0.394 | | ^7,9^ |
| Mild/moderate to severe state | NA | NA | 0.143 | 0.284 | | ^11^ |
| Severe to critical illness state | NA | NA | NA | 0.105 | | ^12^ |
| Critical illness to recuperation state | NA | NA | NA | 0.049 | | ^12^ |
| Daily probability of transition between disease states, stratified by COVID-19 severity, among non-hospitalized | Asymptomatic infection | Mild/moderate illness | Severe illness | Critical illness | |  |
| Pre-infectious latent to asymptomatic state | 0.323 | 0.323 | 0.323 | 0.323 | | Derived from ^7–10^ |
| Asymptomatic to mild/moderate state | NA | 0.394 | 0.394 | 0.394 | | ^7,9^ |
| Mild/moderate to severe state | NA | NA | 0.143 | 0.284 | | ^11^ |
| Severe to critical illness state | NA | NA | NA | 0.143 | | ^12^ |

**eTable1 continued. Additional input parameters for an analysis of management strategies for people experiencing sheltered homelessness during the COVID-19 pandemic.**

| **Parameter** | | **Value** | | | | **Source** |
| --- | --- | --- | --- | --- | --- | --- |
| **Natural history** | |  | | | | |
| Daily probability of transition to recovery, stratified by COVID-19 severity, among hospitalized | Asymptomatic infection | | Mild/moderate illness | Severe illness | Critical illness |  |
| Asymptomatic state to recovery | 0.099 | | NA | NA | NA | Derived from ^13^ |
| Mild/moderate state to recovery | NA | | 0.095 | NA | NA | Derived from ^9^ |
| Severe state to recovery | NA | | NA | 0.091 | 0.026 | ^12^ |
| Recuperation to recovery | NA | | NA | NA | 0.161 | ^12^ |
| Daily probability of transition to recovery, stratified by COVID-19 severity, among non-hospitalized | Asymptomatic infection | | Mild/moderate illness | Severe illness | Critical illness |  |
| Asymptomatic state to recovery | 0.099 | | NA | NA | NA | Derived from ^13^ |
| Mild/moderate state to recovery | NA | | 0.095 | NA | NA | Derived from ^9^ |
| Severe state to recovery | NA | | NA | 0.063 | 0.000 | ^12^ |
| Recuperation to recovery | NA | | NA | NA | 0.000 |  |

Abbreviations: Asymp., asymptomatic; COVID-19, coronavirus disease 2019; y, year.

*To obtain the initial illness distribution, we seeded the model with 0.01% of the starting cohort infected with the SARS-CoV-2 virus. We simulated the model for 10 days until the prevalence of SARS-CoV-2 in the cohort reached 2.2%. We used the illness distribution on Day 10 from this initialization run to inform our initial illness distribution in all model runs.

**eTable2. One-way sensitivity analysis on PCR sensitivity for people with mild/moderate illness.**

| **Effective reproduction number (R_e_)** | **PCR sensitivity for people with mild/mod. illness, %** | **Strategy** | **Cumulative infections, n** | **Total cost *,**  **2020 USD** | **Incr. cost per case prevented *, 2020 USD** |
| --- | --- | --- | --- | --- | --- |
| 2.6 | 70 |  |  |  |  |
|  | base case | *SxScreen/PCR/ACS* | 1,239 | 3,267,000 | NA |
|  |  | *Hybrid/ACS* | 985 | 3,628,000 | 1,000 |
|  |  | *UniversalPCR/ACS* | 1,681 | 4,143,000 | Dominated |
|  |  | *NoIntervention* | 1,954 | 6,098,000 | Dominated |
|  |  | *Hybrid/Hospital* | 967 | 12,202,000 | Dominated |
|  |  | *SxScreen/PCR/Hospital* | 1,133 | 12,620,000 | Dominated |
|  |  | *UniversalPCR/Hospital* | 1,679 | 12,914,000 | Dominated |
|  |  | *UniversalPCR/TempHousing* | 376 | 39,119,000 | 58,000 |
|  | 80 |  |  |  |  |
|  |  | *SxScreen/PCR/ACS* | 1,154 | 3,128,000 | NA |
|  |  | *Hybrid/ACS* | 825 | 3,324,000 | 1,000 |
|  |  | *UniversalPCR/ACS* | 1,616 | 4,214,000 | Dominated |
|  |  | *NoIntervention* | 1,948 | 6,504,000 | Dominated |
|  |  | *Hybrid/Hospital* | 809 | 10,854,000 | Dominated |
|  |  | *SxScreen/PCR/Hospital* | 1,045 | 12,156,000 | Dominated |
|  |  | *UniversalPCR/Hospital* | 1,613 | 13,451,000 | Dominated |
|  |  | *UniversalPCR/TempHousing* | 380 | 39,756,000 | 82,000 |
|  | 90 |  |  |  |  |
|  |  | *SxScreen/PCR/ACS* | 1,067 | 2,940,000 | NA |
|  |  | *Hybrid/ACS* | 668 | 2,966,000 | 100 |
|  |  | *UniversalPCR/ACS* | 1,546 | 4,222,000 | Dominated |
|  |  | *NoIntervention* | 1,935 | 6,835,000 | Dominated |
|  |  | *Hybrid/Hospital* | 658 | 9,377,000 | Dominated |
|  |  | *SxScreen/PCR/Hospital* | 956 | 11,384,000 | Dominated |
|  |  | *UniversalPCR/Hospital* | 1,548 | 13,824,000 | Dominated |
|  |  | *UniversalPCR/TempHousing* | 365 | 39,766,000 | 122,000 |

**eTable2 continued. One-way sensitivity analysis on PCR sensitivity for people with mild/moderate illness.**

| **Effective reproduction number (R_e_)** | **PCR sensitivity for people with mild/mod. illness, %** | **Strategy** | **Cumulative infections, n** | **Total cost *,**  **2020 USD** | **Incr. cost per case prevented *, 2020 USD** |
| --- | --- | --- | --- | --- | --- |
| 2.6 | 100 |  |  |  |  |
|  |  | *Hybrid/ACS* | 512 | 2,556,000 | NA |
|  |  | *SxScreen/PCR/ACS* | 994 | 2,777,000 | Dominated |
|  |  | *UniversalPCR/ACS* | 1,473 | 4,188,000 | Dominated |
|  |  | *NoIntervention* | 1,929 | 7,211,000 | Dominated |
|  |  | *Hybrid/Hospital* | 514 | 7,834,000 | Dominated |
|  |  | *SxScreen/PCR/Hospital* | 887 | 10,824,000 | Dominated |
|  |  | *UniversalPCR/Hospital* | 1,478 | 13,897,000 | Dominated |
|  |  | *UniversalPCR/TempHousing* | 375 | 39,760,000 | Dominated |
| 1.3 | 70 |  |  |  |  |
|  | base case | *SxScreen/PCR/ACS* | 137 | 409,000 | NA |
|  |  | *Hybrid/ACS* | 103 | 1,325,000 | 27,000 |
|  |  | *UniversalPCR/ACS* | 207 | 1,426,000 | Dominated |
|  |  | *NoIntervention* | 538 | 1,461,000 | Dominated |
|  |  | *SxScreen/PCR/Hospital* | 125 | 1,604,000 | Dominated |
|  |  | *Hybrid/Hospital* | 100 | 2,368,000 | 382,000 |
|  |  | *UniversalPCR/Hospital* | 207 | 2,631,000 | Dominated |
|  |  | *UniversalPCR/TempHousing* | 95 | 38,974,000 | 6,854,000 |
|  | 80 |  |  |  |  |
|  |  | *SxScreen/PCR/ACS* | 127 | 392,000 | NA |
|  |  | *Hybrid/ACS* | 92 | 1,331,000 | 27,000 |
|  |  | *UniversalPCR/ACS* | 177 | 1,414,000 | Dominated |
|  |  | *NoIntervention* | 532 | 1,555,000 | Dominated |
|  |  | *SxScreen/PCR/Hospital* | 118 | 1,588,000 | Dominated |
|  |  | *Hybrid/Hospital* | 91 | 2,327,000 | 1,033,000 |
|  |  | *UniversalPCR/Hospital* | 175 | 2,572,000 | Dominated |
|  |  | *UniversalPCR/TempHousing* | 93 | 39,614,000 | Dominated |

**eTable2 continued. One-way sensitivity analysis on PCR sensitivity for people with mild/moderate illness.**

| **Effective reproduction number (R_e_)** | **PCR sensitivity for people with mild/mod. Illness, %** | **Strategy** | **Cumulative infections, n** | **Total cost *,**  **2020 USD** | **Incr. cost per case prevented *, 2020 USD** |
| --- | --- | --- | --- | --- | --- |
| 1.3 | 90 |  |  |  |  |
|  |  | *SxScreen/PCR/ACS* | 121 | 383,000 | NA |
|  |  | *Hybrid/ACS* | 85 | 1,319,000 | 27,000 |
|  |  | *UniversalPCR/ACS* | 148 | 1,384,000 | Dominated |
|  |  | *SxScreen/PCR/Hospital* | 111 | 1,546,000 | Dominated |
|  |  | *NoIntervention* | 518 | 1,599,000 | Dominated |
|  |  | *Hybrid/Hospital* | 84 | 2,292,000 | 816,000 |
|  |  | *UniversalPCR/Hospital* | 147 | 2,486,000 | Dominated |
|  |  | *UniversalPCR/TempHousing* | 93 | 39,619,000 | Dominated |
|  | 100 |  |  |  |  |
|  |  | *SxScreen/PCR/ACS* | 114 | 377,000 | NA |
|  |  | *Hybrid/ACS* | 79 | 1,307,000 | 26,000 |
|  |  | *UniversalPCR/ACS* | 134 | 1,373,000 | Dominated |
|  |  | *SxScreen/PCR/Hospital* | 109 | 1,545,000 | Dominated |
|  |  | *NoIntervention* | 498 | 1,645,000 | Dominated |
|  |  | *Hybrid/Hospital* | 79 | 2,258,000 | Dominated |
|  |  | *UniversalPCR/Hospital* | 131 | 2,444,000 | Dominated |
|  |  | *UniversalPCR/TempHousing* | 93 | 39,613,000 | Dominated |
| 0.9 | 70 |  |  |  |  |
|  | base case | *SxScreen/PCR/ACS* | 85 | 264,000 | NA |
|  |  | *NoIntervention* | 174 | 540,000 | Dominated |
|  |  | *SxScreen/PCR/Hospital* | 82 | 1,113,000 | Dominated |
|  |  | *UniversalPCR/ACS* | 94 | 1,226,000 | Dominated |
|  |  | *Hybrid/ACS* | 71 | 1,240,000 | 71,000 |
|  |  | *UniversalPCR/Hospital* | 95 | 1,901,000 | Dominated |
|  |  | *Hybrid/Hospital* | 71 | 2,004,000 | Dominated |
|  |  | *UniversalPCR/TempHousing* | 71 | 38,954,000 | Dominated |

**eTable2 continued. One-way sensitivity analysis on PCR sensitivity for people with mild/moderate illness.**

| **Effective reproduction number (R_e_)** | **PCR sensitivity for people with mild/mod. illness, %** | **Strategy** | **Cumulative infections, n** | **Total cost *,**  **2020 USD** | **Incr. cost per case prevented *, 2020 USD** |
| --- | --- | --- | --- | --- | --- |
| 0.9 | 80 |  |  |  |  |
|  |  | *SxScreen/PCR/ACS* | 82 | 259,000 | NA |
|  |  | *NoIntervention* | 171 | 569,000 | Dominated |
|  |  | *SxScreen/PCR/Hospital* | 78 | 1,109,000 | Dominated |
|  |  | *UniversalPCR/ACS* | 87 | 1,252,000 | Dominated |
|  |  | *Hybrid/ACS* | 68 | 1,257,000 | 74,000 |
|  |  | *UniversalPCR/Hospital* | 87 | 1,944,000 | Dominated |
|  |  | *Hybrid/Hospital* | 67 | 2,001,000 | 677,000 |
|  |  | *UniversalPCR/TempHousing* | 72 | 39,601,000 | Dominated |
|  | 90 |  |  |  |  |
|  |  | *SxScreen/PCR/ACS* | 80 | 263,000 | NA |
|  |  | *NoIntervention* | 169 | 612,000 | Dominated |
|  |  | *SxScreen/PCR/Hospital* | 76 | 1,105,000 | Dominated |
|  |  | *Hybrid/ACS* | 65 | 1,252,000 | 67,000 |
|  |  | *UniversalPCR/ACS* | 83 | 1,253,000 | Dominated |
|  |  | *UniversalPCR/Hospital* | 82 | 1,973,000 | Dominated |
|  |  | *Hybrid/Hospital* | 64 | 2,024,000 | 1,036,000 |
|  |  | *UniversalPCR/TempHousing* | 72 | 39,604,000 | Dominated |
|  | 100 |  |  |  |  |
|  |  | *SxScreen/PCR/ACS* | 78 | 269,000 | NA |
|  |  | *NoIntervention* | 164 | 628,000 | Dominated |
|  |  | *SxScreen/PCR/Hospital* | 74 | 1,123,000 | Dominated |
|  |  | *UniversalPCR/ACS* | 76 | 1,249,000 | Dominated |
|  |  | *Hybrid/ACS* | 63 | 1,254,000 | 65,000 |
|  |  | *UniversalPCR/Hospital* | 77 | 1,973,000 | Dominated |
|  |  | *Hybrid/Hospital* | 62 | 2,035,000 | 3,641,000 |
|  |  | *UniversalPCR/TempHousing* | 71 | 39,596,000 | Dominated |

Abbreviations: ACS, alternate care sites; COVID-19, coronavirus disease 2019; Dominated, less clinically effective and more costly than an alternative strategy, or a combination of two alternative strategies;^14^ Incr., incremental; mild/mod., mild/moderate; *PCR,* polymerase chain reaction; *SxScreen,* symptom screen; *TempHousing,* temporary housing; *UniversalPCR*, universal polymerase chain reaction test for everyone; USD, United States dollars.

Strategies are listed in order of ascending costs, per convention of cost-effectiveness analysis.

* Costs were rounded to the nearest thousands.

**eTable3. One-way sensitivity analysis on universal PCR testing frequency.**

| **Effective reproduction number (R_e_)** | **Universal PCR testing frequency** | **Strategy** | **Cumulative infections, n** | **Total cost *,**  **2020 USD** | **Incr. cost per case prevented *, 2020 USD** |
| --- | --- | --- | --- | --- | --- |
| 2.6 | Every 30 days |  |  |  |  |
|  |  | *UniversalPCR/ACS* | 1,880 | 3,131,000 | NA |
|  |  | *SxScreen/PCR/ACS* | 1,244 | 3,295,000 | 300 |
|  |  | *Hybrid/ACS* | 1,179 | 3,594,000 | 5,000 |
|  |  | *NoIntervention* | 1,954 | 6,114,000 | Dominated |
|  |  | *UniversalPCR/Hospital* | 1,876 | 8,912,000 | Dominated |
|  |  | *SxScreen/PCR/Hospital* | 1,138 | 12,738,000 | Dominated |
|  |  | *Hybrid/Hospital* | 1,165 | 13,224,000 | Dominated |
|  |  | *UniversalPCR/TempHousing* | 374 | 39,302,000 | 44,000 |
|  | Every 14 days |  |  |  |  |
|  | base case | *SxScreen/PCR/ACS* | 1,239 | 3,267,000 | NA |
|  |  | *Hybrid/ACS* | 985 | 3,628,000 | 1,000 |
|  |  | *UniversalPCR/ACS* | 1,681 | 4,143,000 | Dominated |
|  |  | *NoIntervention* | 1,954 | 6,098,000 | Dominated |
|  |  | *Hybrid/Hospital* | 967 | 12,202,000 | Dominated |
|  |  | *SxScreen/PCR/Hospital* | 1,133 | 12,620,000 | Dominated |
|  |  | *UniversalPCR/Hospital* | 1,679 | 12,914,000 | Dominated |
|  |  | *UniversalPCR/TempHousing* | 376 | 39,119,000 | 58,000 |
|  | Every 7 days |  |  |  |  |
|  |  | *SxScreen/PCR/ACS* | 1,244 | 3,295,000 | NA |
|  |  | *Hybrid/ACS* | 700 | 3,818,000 | Dominated |
|  |  | *UniversalPCR/ACS* | 1,278 | 4,862,000 | Dominated |
|  |  | *NoIntervention* | 1,954 | 6,114,000 | Dominated |
|  |  | *Hybrid/Hospital* | 679 | 10,340,000 | Dominated |
|  |  | *SxScreen/PCR/Hospital* | 1,138 | 12,738,000 | Dominated |
|  |  | *UniversalPCR/Hospital* | 1,278 | 14,554,000 | Dominated |
|  |  | *UniversalPCR/TempHousing* | 375 | 40,555,000 | 43,000 |

**eTable3 continued. One-way sensitivity analysis on universal PCR testing frequency.**

| **Effective reproduction number (R_e_)** | **Universal PCR testing frequency** | **Strategy** | **Cumulative infections, n** | **Total cost *,**  **2020 USD** | **Incr. cost per case prevented *, 2020 USD** |
| --- | --- | --- | --- | --- | --- |
| 2.6 | Every 3 days |  |  |  |  |
|  |  | *SxScreen/PCR/ACS* | 1,244 | 3,295,000 | NA |
|  |  | *Hybrid/ACS* | 334 | 4,609,000 | 1,000 |
|  |  | *UniversalPCR/ACS* | 444 | 4,872,000 | Dominated |
|  |  | *NoIntervention* | 1,954 | 6,114,000 | Dominated |
|  |  | *Hybrid/Hospital* | 317 | 8,152,000 | Dominated |
|  |  | *UniversalPCR/Hospital* | 437 | 9,455,000 | Dominated |
|  |  | *SxScreen/PCR/Hospital* | 1,138 | 12,738,000 | Dominated |
|  |  | *UniversalPCR/TempHousing* | 376 | 42,288,000 | Dominated |
| 1.3 | Every 30 days |  |  |  |  |
|  |  | *SxScreen/PCR/ACS* | 137 | 409,000 | NA |
|  |  | *Hybrid/ACS* | 115 | 896,000 | 22,000 |
|  |  | *UniversalPCR/ACS* | 338 | 1,008,000 | Dominated |
|  |  | *NoIntervention* | 544 | 1,485,000 | Dominated |
|  |  | *SxScreen/PCR/Hospital* | 125 | 1,608,000 | Dominated |
|  |  | *Hybrid/Hospital* | 112 | 1,966,000 | 312,000 |
|  |  | *UniversalPCR/Hospital* | 345 | 2,136,000 | Dominated |
|  |  | *UniversalPCR/TempHousing* | 95 | 39,145,000 | 2,191,000 |
|  | Every 14 days |  |  |  |  |
|  | base case | *SxScreen/PCR/ACS* | 137 | 409,000 | NA |
|  |  | *Hybrid/ACS* | 103 | 1,325,000 | 27,000 |
|  |  | *UniversalPCR/ACS* | 207 | 1,426,000 | Dominated |
|  |  | *NoIntervention* | 538 | 1,461,000 | Dominated |
|  |  | *SxScreen/PCR/Hospital* | 125 | 1,604,000 | Dominated |
|  |  | *Hybrid/Hospital* | 100 | 2,368,000 | 382,000 |
|  |  | *UniversalPCR/Hospital* | 207 | 2,631,000 | Dominated |
|  |  | *UniversalPCR/TempHousing* | 95 | 38,974,000 | 6,854,000 |

**eTable3 continued. One-way sensitivity analysis on universal PCR testing frequency.**

| **Effective reproduction number (R_e_)** | **Universal PCR testing frequency** | **Strategy** | **Cumulative infections, n** | **Total cost *,**  **2020 USD** | **Incr. cost per case prevented *, 2020 USD** |
| --- | --- | --- | --- | --- | --- |
| 1.3 | Every 7 days |  |  |  |  |
|  |  | *SxScreen/PCR/ACS* | 137 | 409,000 | NA |
|  |  | *NoIntervention* | 544 | 1,485,000 | Dominated |
|  |  | *SxScreen/PCR/Hospital* | 125 | 1,608,000 | Dominated |
|  |  | *Hybrid/ACS* | 91 | 2,123,000 | 37,000 |
|  |  | *UniversalPCR/ACS* | 124 | 2,162,000 | Dominated |
|  |  | *Hybrid/Hospital* | 90 | 3,122,000 | 739,000 |
|  |  | *UniversalPCR/Hospital* | 120 | 3,217,000 | Dominated |
|  |  | *UniversalPCR/TempHousing* | 91 | 40,423,000 | Dominated |
|  | Every 3 days |  |  |  |  |
|  |  | *SxScreen/PCR/ACS* | 137 | 409,000 | NA |
|  |  | *NoIntervention* | 544 | 1,485,000 | Dominated |
|  |  | *SxScreen/PCR/Hospital* | 125 | 1,608,000 | Dominated |
|  |  | *UniversalPCR/ACS* | 83 | 3,815,000 | Dominated |
|  |  | *Hybrid/ACS* | 79 | 3,817,000 | 59,000 |
|  |  | *UniversalPCR/Hospital* | 82 | 4,757,000 | Dominated |
|  |  | *Hybrid/Hospital* | 78 | 4,780,000 | 1,119,000 |
|  |  | *UniversalPCR/TempHousing* | 95 | 42,141,000 | Dominated |
| 0.9 | Every 30 days |  |  |  |  |
|  |  | *SxScreen/PCR/ACS* | 85 | 264,000 | NA |
|  |  | *NoIntervention* | 175 | 543,000 | Dominated |
|  |  | *UniversalPCR/ACS* | 118 | 768,000 | Dominated |
|  |  | *Hybrid/ACS* | 74 | 785,000 | 46,000 |
|  |  | *SxScreen/PCR/Hospital* | 82 | 1,115,000 | Dominated |
|  |  | *UniversalPCR/Hospital* | 121 | 1,349,000 | Dominated |
|  |  | *Hybrid/Hospital* | 75 | 1,555,000 | Dominated |
|  |  | *UniversalPCR/TempHousing* | 72 | 39,131,000 | 21,790,000 |

**eTable3 continued. One-way sensitivity analysis on universal PCR testing frequency.**

| **Effective reproduction number (R_e_)** | **Universal PCR testing frequency** | **Strategy** | **Cumulative infections, n** | **Total cost *,**  **2020 USD** | **Incr. cost per case prevented *, 2020 USD** |
| --- | --- | --- | --- | --- | --- |
| 0.9 | Every 14 days |  |  |  |  |
|  | base case | *SxScreen/PCR/ACS* | 85 | 264,000 | NA |
|  |  | *NoIntervention* | 174 | 540,000 | Dominated |
|  |  | *SxScreen/PCR/Hospital* | 82 | 1,113,000 | Dominated |
|  |  | *UniversalPCR/ACS* | 94 | 1,226,000 | Dominated |
|  |  | *Hybrid/ACS* | 71 | 1,240,000 | 71,000 |
|  |  | *UniversalPCR/Hospital* | 95 | 1,901,000 | Dominated |
|  |  | *Hybrid/Hospital* | 71 | 2,004,000 | Dominated |
|  |  | *UniversalPCR/TempHousing* | 71 | 38,954,000 | Dominated |
|  | Every 7 days |  |  |  |  |
|  |  | *SxScreen/PCR/ACS* | 85 | 264,000 | NA |
|  |  | *NoIntervention* | 175 | 543,000 | Dominated |
|  |  | *SxScreen/PCR/Hospital* | 82 | 1,115,000 | Dominated |
|  |  | *UniversalPCR/ACS* | 78 | 2,049,000 | Dominated |
|  |  | *Hybrid/ACS* | 68 | 2,057,000 | 103,000 |
|  |  | *UniversalPCR/Hospital* | 76 | 2,776,000 | Dominated |
|  |  | *Hybrid/Hospital* | 67 | 2,810,000 | Dominated |
|  |  | *UniversalPCR/TempHousing* | 71 | 40,407,000 | Dominated |
|  | Every 3 days |  |  |  |  |
|  |  | *SxScreen/PCR/ACS* | 85 | 264,000 | NA |
|  |  | *NoIntervention* | 175 | 543,000 | Dominated |
|  |  | *SxScreen/PCR/Hospital* | 82 | 1,115,000 | Dominated |
|  |  | *Hybrid/ACS* | 64 | 3,763,000 | 164,000 |
|  |  | *UniversalPCR/ACS* | 65 | 3,764,000 | Dominated |
|  |  | *UniversalPCR/Hospital* | 65 | 4,527,000 | Dominated |
|  |  | *Hybrid/Hospital* | 64 | 4,553,000 | Dominated |
|  |  | *UniversalPCR/TempHousing* | 71 | 42,126,000 | Dominated |

Abbreviations: ACS, alternate care sites; COVID-19, coronavirus disease 2019; Dominated, less clinically effective and more costly than an alternative strategy, or a combination of two alternative strategies;^14^ Incr., incremental; *PCR,* polymerase chain reaction; *SxScreen,* symptom screen; *TempHousing,* temporary housing; *UniversalPCR*, universal polymerase chain reaction test for everyone; USD, United States dollars.

Strategies are listed in order of ascending costs, per convention of cost-effectiveness analysis.

* Costs were rounded to the nearest thousands.

**eTable4. One-way sensitivity analysis on symptom screen sensitivity for people with mild/moderate illness.**

| **Effective reproduction number (R_e_)** | **Sx screen sensitivity for people with mild/mod. illness, %** | **Strategy** | **Cumulative infections, n** | **Total cost *,**  **2020 USD** | **Incr. cost per case prevented *, 2020 USD** |
| --- | --- | --- | --- | --- | --- |
| 2.6 | 50 |  |  |  |  |
|  |  | *SxScreen/PCR/ACS* | 1,306 | 3,376,000 | NA |
|  |  | *Hybrid/ACS* | 1,037 | 3,784,000 | 2,000 |
|  |  | *UniversalPCR/ACS* | 1,681 | 4,143,000 | Dominated |
|  |  | *NoIntervention* | 1,954 | 6,098,000 | Dominated |
|  |  | *Hybrid/Hospital* | 1,015 | 12,598,000 | Dominated |
|  |  | *UniversalPCR/Hospital* | 1,679 | 12,914,000 | Dominated |
|  |  | *SxScreen/PCR/Hospital* | 1,214 | 13,313,000 | Dominated |
|  |  | *UniversalPCR/TempHousing* | 376 | 39,119,000 | 53,000 |
|  | 62 |  |  |  |  |
|  | base case | *SxScreen/PCR/ACS* | 1,239 | 3,267,000 | NA |
|  |  | *Hybrid/ACS* | 985 | 3,628,000 | 1,000 |
|  |  | *UniversalPCR/ACS* | 1,681 | 4,143,000 | Dominated |
|  |  | *NoIntervention* | 1,954 | 6,098,000 | Dominated |
|  |  | *Hybrid/Hospital* | 967 | 12,202,000 | Dominated |
|  |  | *SxScreen/PCR/Hospital* | 1,133 | 12,620,000 | Dominated |
|  |  | *UniversalPCR/Hospital* | 1,679 | 12,914,000 | Dominated |
|  |  | *UniversalPCR/TempHousing* | 376 | 39,119,000 | 58,000 |
|  | 70 |  |  |  |  |
|  |  | *SxScreen/PCR/ACS* | 1,210 | 3,240,000 | NA |
|  |  | *Hybrid/ACS* | 965 | 3,634,000 | 2,000 |
|  |  | *UniversalPCR/ACS* | 1,681 | 4,143,000 | Dominated |
|  |  | *NoIntervention* | 1,954 | 6,098,000 | Dominated |
|  |  | *Hybrid/Hospital* | 933 | 11,919,000 | Dominated |
|  |  | *SxScreen/PCR/Hospital* | 1,103 | 12,556,000 | Dominated |
|  |  | *UniversalPCR/Hospital* | 1,679 | 12,914,000 | Dominated |
|  |  | *UniversalPCR/TempHousing* | 376 | 39,119,000 | 60,000 |

**eTable4 continued. One-way sensitivity analysis on symptom screen sensitivity for people with mild/moderate illness.**

| **Effective reproduction number (R_e_)** | **Sx screen sensitivity for people with mild/mod. illness, %** | **Strategy** | **Cumulative infections, n** | **Total cost *,**  **2020 USD** | **Incr. cost per case prevented *, 2020 USD** |
| --- | --- | --- | --- | --- | --- |
| 2.6 | 80 |  |  |  |  |
|  |  | *SxScreen/PCR/ACS* | 1,181 | 3,167,000 | NA |
|  |  | *Hybrid/ACS* | 940 | 3,593,000 | 2,000 |
|  |  | *UniversalPCR/ACS* | 1,681 | 4,143,000 | Dominated |
|  |  | *NoIntervention* | 1,954 | 6,098,000 | Dominated |
|  |  | *Hybrid/Hospital* | 909 | 11,785,000 | Dominated |
|  |  | *SxScreen/PCR/Hospital* | 1,062 | 12,257,000 | Dominated |
|  |  | *UniversalPCR/Hospital* | 1,680 | 12,986,000 | Dominated |
|  |  | *UniversalPCR/TempHousing* | 376 | 39,119,000 | 63,000 |
| 1.3 | 50 |  |  |  |  |
|  |  | *SxScreen/PCR/ACS* | 147 | 426,000 | NA |
|  |  | *Hybrid/ACS* | 105 | 1,349,000 | 22,000 |
|  |  | *UniversalPCR/ACS* | 207 | 1,426,000 | Dominated |
|  |  | *NoIntervention* | 538 | 1,461,000 | Dominated |
|  |  | *SxScreen/PCR/Hospital* | 134 | 1,659,000 | Dominated |
|  |  | *Hybrid/Hospital* | 102 | 2,384,000 | 345,000 |
|  |  | *UniversalPCR/Hospital* | 207 | 2,631,000 | Dominated |
|  |  | *UniversalPCR/TempHousing* | 95 | 38,974,000 | 5,254,000 |
|  | 62 |  |  |  |  |
|  | base case | *SxScreen/PCR/ACS* | 137 | 409,000 | NA |
|  |  | *Hybrid/ACS* | 103 | 1,325,000 | 27,000 |
|  |  | *UniversalPCR/ACS* | 207 | 1,426,000 | Dominated |
|  |  | *NoIntervention* | 538 | 1,461,000 | Dominated |
|  |  | *SxScreen/PCR/Hospital* | 125 | 1,604,000 | Dominated |
|  |  | *Hybrid/Hospital* | 100 | 2,368,000 | 348,000 |
|  |  | *UniversalPCR/Hospital* | 207 | 2,631,000 | Dominated |
|  |  | *UniversalPCR/TempHousing* | 95 | 38,974,000 | 6,854,000 |

**eTable4 continued. One-way sensitivity analysis on symptom screen sensitivity for people with mild/moderate illness.**

| **Effective reproduction number (R_e_)** | **Sx screen sensitivity for people with mild/mod. illness, %** | **Strategy** | **Cumulative infections, n** | **Total cost *,**  **2020 USD** | **Incr. cost per case prevented *, 2020 USD** |
| --- | --- | --- | --- | --- | --- |
| 1.3 | 70 |  |  |  |  |
|  |  | *SxScreen/PCR/ACS* | 134 | 403,000 | NA |
|  |  | *Hybrid/ACS* | 103 | 1,346,000 | 30,000 |
|  |  | *UniversalPCR/ACS* | 207 | 1,426,000 | Dominated |
|  |  | *NoIntervention* | 538 | 1,461,000 | Dominated |
|  |  | *SxScreen/PCR/Hospital* | 120 | 1,589,000 | Dominated |
|  |  | *Hybrid/Hospital* | 99 | 2,365,000 | 255,000 |
|  |  | *UniversalPCR/Hospital* | 207 | 2,631,000 | Dominated |
|  |  | *UniversalPCR/TempHousing* | 95 | 38,974,000 | 9,735,000 |
|  | 80 |  |  |  |  |
|  |  | *SxScreen/PCR/ACS* | 130 | 399,000 | NA |
|  |  | *Hybrid/ACS* | 101 | 1,343,000 | 33,000 |
|  |  | *UniversalPCR/ACS* | 207 | 1,426,000 | Dominated |
|  |  | *NoIntervention* | 538 | 1,461,000 | Dominated |
|  |  | *SxScreen/PCR/Hospital* | 120 | 1,598,000 | Dominated |
|  |  | *Hybrid/Hospital* | 97 | 2,359,000 | 254,000 |
|  |  | *UniversalPCR/Hospital* | 207 | 2,631,000 | Dominated |
|  |  | *UniversalPCR/TempHousing* | 95 | 39,974,000 | 16,390,000 |
| 0.9 | 50 |  |  |  |  |
|  |  | *SxScreen/PCR/ACS* | 86 | 265,000 | NA |
|  |  | *NoIntervention* | 174 | 540,000 | Dominated |
|  |  | *SxScreen/PCR/Hospital* | 83 | 1,090,000 | Dominated |
|  |  | *UniversalPCR/ACS* | 94 | 1,226,000 | Dominated |
|  |  | *Hybrid/ACS* | 73 | 1,258,000 | 76,000 |
|  |  | *UniversalPCR/Hospital* | 95 | 1,901,000 | Dominated |
|  |  | *Hybrid/Hospital* | 72 | 2,007,000 | 749,000 |
|  |  | *UniversalPCR/TempHousing* | 71 | 38,954,000 | 58,807,000 |

**eTable4 continued. One-way sensitivity analysis on symptom screen sensitivity for people with mild/moderate illness.**

| **Effective reproduction number (R_e_)** | **Sx screen sensitivity for people with mild/mod. illness, %** | **Strategy** | **Cumulative infections, n** | **Total cost *,**  **2020 USD** | **Incr. cost per case prevented *, 2020 USD** |
| --- | --- | --- | --- | --- | --- |
| 0.9 | 62 |  |  |  |  |
|  | base case | *SxScreen/PCR/ACS* | 85 | 264,000 | NA |
|  |  | *NoIntervention* | 174 | 540,000 | Dominated |
|  |  | *SxScreen/PCR/Hospital* | 82 | 1,113,000 | Dominated |
|  |  | *UniversalPCR/ACS* | 94 | 1,226,000 | Dominated |
|  |  | *Hybrid/ACS* | 71 | 1,240,000 | 70,000 |
|  |  | *UniversalPCR/Hospital* | 95 | 1,901,000 | Dominated |
|  |  | *Hybrid/Hospital* | 71 | 2,004,000 | Dominated |
|  |  | *UniversalPCR/TempHousing* | 71 | 38,954,000 | Dominated |
|  | 70 |  |  |  |  |
|  |  | *SxScreen/PCR/ACS* | 82 | 262,000 | NA |
|  |  | *NoIntervention* | 174 | 540,000 | Dominated |
|  |  | *SxScreen/PCR/Hospital* | 80 | 1,101,000 | Dominated |
|  |  | *UniversalPCR/ACS* | 94 | 1,226,000 | Dominated |
|  |  | *Hybrid/ACS* | 71 | 1,256,000 | 90,000 |
|  |  | *UniversalPCR/Hospital* | 95 | 1,901,000 | Dominated |
|  |  | *Hybrid/Hospital* | 71 | 2,020,000 | Dominated |
|  |  | *UniversalPCR/TempHousing* | 71 | 38,954,000 | Dominated |
|  | 80 |  |  |  |  |
|  |  | *SxScreen/PCR/ACS* | 82 | 259,000 | NA |
|  |  | *NoIntervention* | 174 | 540,000 | Dominated |
|  |  | *SxScreen/PCR/Hospital* | 80 | 1,115,000 | Dominated |
|  |  | *UniversalPCR/ACS* | 94 | 1,226,000 | Dominated |
|  |  | *Hybrid/ACS* | 70 | 1,259,000 | 83,000 |
|  |  | *UniversalPCR/Hospital* | 95 | 1,901,000 | Dominated |
|  |  | *Hybrid/Hospital* | 70 | 2,014,000 | Dominated |
|  |  | *UniversalPCR/TempHousing* | 71 | 38,954,000 | Dominated |

Abbreviations: ACS, alternate care sites; COVID-19, coronavirus disease 2019; Dominated, less clinically effective and more costly than an alternative strategy, or a combination of two alternative strategies;^14^ Incr., incremental; mild/mod., mild/moderate; *PCR,* polymerase chain reaction; Sx, symptom; *SxScreen,* symptom screen; *TempHousing,* temporary housing; *UniversalPCR*, universal polymerase chain reaction test for everyone; USD, United States dollars.

Strategies are listed in order of ascending costs, per convention of cost-effectiveness analysis.

* Costs were rounded to the nearest thousands.

**eTable5. One-way sensitivity analysis on efficacy of ACS for COVID-confirmed in reducing SARS-CoV-2 transmission.**

| **Effective reproduction number (R_e_)** | **Efficacy of ACS-C in reducing SARS-CoV-2 transmission, %** | **Strategy** | **Cumulative infections, n** | **Total cost *,**  **2020 USD** | **Incr. cost per case prevented *, 2020 USD** |
| --- | --- | --- | --- | --- | --- |
| 2.6 | 70 |  |  |  |  |
|  |  | *SxScreen/PCR/ACS* | 1,573 | 4,240,000 | NA |
|  |  | *UniversalPCR/ACS* | 1,831 | 4,494,000 | Dominated |
|  |  | *Hybrid/ACS* | 1,529 | 5,252,000 | Dominated |
|  |  | *NoIntervention* | 1,954 | 6,114,000 | Dominated |
|  |  | *Hybrid/Hospital* | 971 | 12,321,000 | 13,000 |
|  |  | *SxScreen/PCR/Hospital* | 1,138 | 12,738,000 | Dominated |
|  |  | *UniversalPCR/Hospital* | 1,680 | 12,986,000 | Dominated |
|  |  | *UniversalPCR/TempHousing* | 379 | 39,772,000 | 46,000 |
|  | 80 |  |  |  |  |
|  |  | *SxScreen/PCR/ACS* | 1,477 | 3,961,000 | NA |
|  |  | *UniversalPCR/ACS* | 1,786 | 4,391,000 | Dominated |
|  |  | *Hybrid/ACS* | 1,387 | 4,813,000 | 9,000 |
|  |  | *NoIntervention* | 1,954 | 6,114,000 | Dominated |
|  |  | *Hybrid/Hospital* | 971 | 12,321,000 | 18,000 |
|  |  | *SxScreen/PCR/Hospital* | 1,138 | 12,738,000 | Dominated |
|  |  | *UniversalPCR/Hospital* | 1,680 | 12,986,000 | Dominated |
|  |  | *UniversalPCR/TempHousing* | 379 | 39,772,000 | 46,000 |
|  | 90 |  |  |  |  |
|  |  | *SxScreen/PCR/ACS* | 1,363 | 3,633,000 | NA |
|  |  | *UniversalPCR/ACS* | 1,740 | 4,282,000 | Dominated |
|  |  | *Hybrid/ACS* | 1,210 | 4,309,000 | 4,000 |
|  |  | *NoIntervention* | 1,954 | 6,114,000 | Dominated |
|  |  | *Hybrid/Hospital* | 971 | 12,321,000 | 34,000 |
|  |  | *SxScreen/PCR/Hospital* | 1,138 | 12,738,000 | Dominated |
|  |  | *UniversalPCR/Hospital* | 1,680 | 12,986,000 | Dominated |
|  |  | *UniversalPCR/TempHousing* | 379 | 39,772,000 | 46,000 |

**eTable5 continued. One-way sensitivity analysis on efficacy of ACS for COVID-confirmed in reducing SARS-CoV-2 transmission.**

| **Effective reproduction number (R_e_)** | **Efficacy of ACS-C in reducing SARS-CoV-2 transmission, %** | **Strategy** | **Cumulative infections, n** | **Total cost *,**  **2020 USD** | **Incr. cost per case prevented *, 2020 USD** |
| --- | --- | --- | --- | --- | --- |
| 2.6 | 100 |  |  |  |  |
|  | base case | *SxScreen/PCR/ACS* | 1,239 | 3,267,000 | NA |
|  |  | *Hybrid/ACS* | 985 | 3,628,000 | 1,000 |
|  |  | *UniversalPCR/ACS* | 1,681 | 4,143,000 | Dominated |
|  |  | *NoIntervention* | 1,954 | 6,098,000 | Dominated |
|  |  | *Hybrid/Hospital* | 967 | 12,202,000 | Dominated |
|  |  | *SxScreen/PCR/Hospital* | 1,133 | 12,620,000 | Dominated |
|  |  | *UniversalPCR/Hospital* | 1,679 | 12,914,000 | Dominated |
|  |  | *UniversalPCR/TempHousing* | 376 | 39,119,000 | 58,000 |
| 1.3 | 70 |  |  |  |  |
|  |  | *SxScreen/PCR/ACS* | 310 | 825,000 | NA |
|  |  | *NoIntervention* | 544 | 1,485,000 | Dominated |
|  |  | *SxScreen/PCR/Hospital* | 125 | 1,608,000 | 4,000 |
|  |  | *UniversalPCR/ACS* | 488 | 1,866,000 | Dominated |
|  |  | *Hybrid/ACS* | 333 | 1,922,000 | Dominated |
|  |  | *Hybrid/Hospital* | 100 | 2,383,000 | 31,000 |
|  |  | *UniversalPCR/Hospital* | 208 | 2,657,000 | Dominated |
|  |  | *UniversalPCR/TempHousing* | 95 | 39,626,000 | 6,964,000 |
|  | 80 |  |  |  |  |
|  |  | *SxScreen/PCR/ACS* | 234 | 641,000 | NA |
|  |  | *NoIntervention* | 544 | 1,485,000 | Dominated |
|  |  | *SxScreen/PCR/Hospital* | 125 | 1,608,000 | 9,000 |
|  |  | *Hybrid/ACS* | 223 | 1,652,000 | Dominated |
|  |  | *UniversalPCR/ACS* | 375 | 1,696,000 | Dominated |
|  |  | *Hybrid/Hospital* | 100 | 2,383,000 | 31,000 |
|  |  | *UniversalPCR/Hospital* | 208 | 2,657,000 | Dominated |
|  |  | *UniversalPCR/TempHousing* | 95 | 39,626,000 | 6,964,000 |

**eTable5 continued. One-way sensitivity analysis on efficacy of ACS for COVID-confirmed in reducing SARS-CoV-2 transmission.**

| **Effective reproduction number (R_e_)** | **Efficacy of ACS-C in reducing SARS-CoV-2 transmission, %** | **Strategy** | **Cumulative infections, n** | **Total cost *,**  **2020 USD** | **Incr. cost per case prevented *, 2020 USD** |
| --- | --- | --- | --- | --- | --- |
| 1.3 | 90 |  |  |  |  |
|  |  | *SxScreen/PCR/ACS* | 174 | 511,000 | NA |
|  |  | *Hybrid/ACS* | 145 | 1,469,000 | Dominated |
|  |  | *NoIntervention* | 544 | 1,485,000 | Dominated |
|  |  | *UniversalPCR/ACS* | 276 | 1,549,000 | Dominated |
|  |  | *SxScreen/PCR/Hospital* | 125 | 1,608,000 | 22,000 |
|  |  | *Hybrid/Hospital* | 100 | 2,383,000 | 31,000 |
|  |  | *UniversalPCR/Hospital* | 208 | 2,657,000 | Dominated |
|  |  | *UniversalPCR/TempHousing* | 95 | 39,626,000 | 6,964,000 |
|  | 100 |  |  |  |  |
|  | base case | *SxScreen/PCR/ACS* | 137 | 409,000 | NA |
|  |  | *Hybrid/ACS* | 103 | 1,325,000 | 27,000 |
|  |  | *UniversalPCR/ACS* | 207 | 1,426,000 | Dominated |
|  |  | *NoIntervention* | 538 | 1,461,000 | Dominated |
|  |  | *SxScreen/PCR/Hospital* | 125 | 1,604,000 | Dominated |
|  |  | *Hybrid/Hospital* | 100 | 2,368,000 | 382,000 |
|  |  | *UniversalPCR/Hospital* | 207 | 2,631,000 | Dominated |
|  |  | *UniversalPCR/TempHousing* | 95 | 38,974,000 | 6,854,000 |
| 0.9 | 70 |  |  |  |  |
|  |  | *SxScreen/PCR/ACS* | 154 | 452,000 | NA |
|  |  | *NoIntervention* | 175 | 543,000 | Dominated |
|  |  | *SxScreen/PCR/Hospital* | 82 | 1,115,000 | 9,000 |
|  |  | *UniversalPCR/ACS* | 200 | 1,429,000 | Dominated |
|  |  | *Hybrid/ACS* | 176 | 1,535,000 | Dominated |
|  |  | *UniversalPCR/Hospital* | 95 | 1,918,000 | Dominated |
|  |  | *Hybrid/Hospital* | 71 | 2,018,000 | 84,000 |
|  |  | *UniversalPCR/TempHousing* | 71 | 39,607,000 | Dominated |

**eTable5 continued. One-way sensitivity analysis on efficacy of ACS for COVID-confirmed in reducing SARS-CoV-2 transmission.**

| **Effective reproduction number (R_e_)** | **Efficacy of ACS-C in reducing SARS-CoV-2 transmission, %** | **Strategy** | **Cumulative infections, n** | **Total cost *,**  **2020 USD** | **Incr. cost per case prevented *, 2020 USD** |
| --- | --- | --- | --- | --- | --- |
| 0.9 | 80 |  |  |  |  |
|  |  | *SxScreen/PCR/ACS* | 124 | 370,000 | NA |
|  |  | *NoIntervention* | 175 | 543,000 | Dominated |
|  |  | *SxScreen/PCR/Hospital* | 82 | 1,115,000 | 18,000 |
|  |  | *UniversalPCR/ACS* | 153 | 1,349,000 | Dominated |
|  |  | *Hybrid/ACS* | 126 | 1,406,000 | Dominated |
|  |  | *UniversalPCR/Hospital* | 95 | 1,918,000 | Dominated |
|  |  | *Hybrid/Hospital* | 71 | 2,018,000 | 84,000 |
|  |  | *UniversalPCR/TempHousing* | 71 | 39,607,000 | Dominated |
|  | 90 |  |  |  |  |
|  |  | *SxScreen/PCR/ACS* | 101 | 307,000 | NA |
|  |  | *NoIntervention* | 175 | 543,000 | Dominated |
|  |  | *SxScreen/PCR/Hospital* | 82 | 1,115,000 | 42,000 |
|  |  | *UniversalPCR/ACS* | 119 | 1,289,000 | Dominated |
|  |  | *Hybrid/ACS* | 94 | 1,321,000 | Dominated |
|  |  | *UniversalPCR/Hospital* | 95 | 1,918,000 | Dominated |
|  |  | *Hybrid/Hospital* | 71 | 2,018,000 | 84,000 |
|  |  | *UniversalPCR/TempHousing* | 71 | 39,607,000 | Dominated |
|  | 100 |  |  |  |  |
|  | base case | *SxScreen/PCR/ACS* | 85 | 264,000 | NA |
|  |  | *NoIntervention* | 174 | 540,000 | Dominated |
|  |  | *SxScreen/PCR/Hospital* | 82 | 1,113,000 | Dominated |
|  |  | *UniversalPCR/ACS* | 94 | 1,226,000 | Dominated |
|  |  | *Hybrid/ACS* | 71 | 1,240,000 | 71,000 |
|  |  | *UniversalPCR/Hospital* | 95 | 1,901,000 | Dominated |
|  |  | *Hybrid/Hospital* | 71 | 2,004,000 | Dominated |
|  |  | *UniversalPCR/TempHousing* | 71 | 38,954,000 | Dominated |

Abbreviations: ACS, alternate care sites; COVID-19, coronavirus disease 2019; Dominated, less clinically effective and more costly than an alternative strategy, or a combination of two alternative strategies;^14^ Incr., incremental; *PCR,* polymerase chain reaction; *SxScreen,* symptom screen; *TempHousing,* temporary housing; *UniversalPCR*, universal polymerase chain reaction test for everyone; USD, United States dollars.

Strategies are listed in order of ascending costs, per convention of cost-effectiveness analysis.

* Costs were rounded to the nearest thousands.

**eTable6. One-way sensitivity analysis on efficacy of temporary housing in reducing SARS-CoV-2 transmission.**

| **Effective reproduction number (R_e_)** | **Efficacy of TempHousing in reducing SARS-CoV-2 transmission, %** | **Strategy** | **Cumulative infections, n** | **Total cost *,**  **2020 USD** | **Incr. cost per case prevented *, 2020 USD** |
| --- | --- | --- | --- | --- | --- |
| 2.6 | 50 |  |  |  |  |
|  |  | *SxScreen/PCR/ACS* | 1,244 | 3,295,000 | NA |
|  |  | *Hybrid/ACS* | 989 | 3,669,000 | 1,000 |
|  |  | *UniversalPCR/ACS* | 1,682 | 4,176,000 | Dominated |
|  |  | *NoIntervention* | 1,954 | 6,114,000 | Dominated |
|  |  | *Hybrid/Hospital* | 971 | 12,321,000 | Dominated |
|  |  | *SxScreen/PCR/Hospital* | 1,138 | 12,738,000 | Dominated |
|  |  | *UniversalPCR/Hospital* | 1,680 | 12,986,000 | Dominated |
|  |  | *UniversalPCR/TempHousing* | 798 | 39,975,000 | 190,000 |
|  | 60 |  |  |  |  |
|  | base case | *SxScreen/PCR/ACS* | 1,239 | 3,267,000 | NA |
|  |  | *Hybrid/ACS* | 985 | 3,628,000 | 1,000 |
|  |  | *UniversalPCR/ACS* | 1,681 | 4,143,000 | Dominated |
|  |  | *NoIntervention* | 1,954 | 6,098,000 | Dominated |
|  |  | *Hybrid/Hospital* | 967 | 12,202,000 | Dominated |
|  |  | *SxScreen/PCR/Hospital* | 1,133 | 12,620,000 | Dominated |
|  |  | *UniversalPCR/Hospital* | 1,679 | 12,914,000 | Dominated |
|  |  | *UniversalPCR/TempHousing* | 376 | 39,119,000 | 58,000 |
|  | 70 |  |  |  |  |
|  |  | *SxScreen/PCR/ACS* | 1,244 | 3,295,000 | NA |
|  |  | *Hybrid/ACS* | 989 | 3,669,000 | 1,000 |
|  |  | *UniversalPCR/ACS* | 1,682 | 4,176,000 | Dominated |
|  |  | *NoIntervention* | 1,954 | 6,114,000 | Dominated |
|  |  | *Hybrid/Hospital* | 971 | 12,321,000 | Dominated |
|  |  | *SxScreen/PCR/Hospital* | 1,138 | 12,738,000 | Dominated |
|  |  | *UniversalPCR/Hospital* | 1,680 | 12,986,000 | Dominated |
|  |  | *UniversalPCR/TempHousing* | 180 | 39,672,000 | 45,000 |

**eTable6 continued. One-way sensitivity analysis on efficacy of temporary housing in reducing SARS-CoV-2 transmission.**

| **Effective reproduction number (R_e_)** | **Efficacy of TempHousing in reducing SARS-CoV-2 transmission, %** | **Strategy** | **Cumulative infections, n** | **Total cost *,**  **2020 USD** | **Incr. cost per case prevented *, 2020 USD** |
| --- | --- | --- | --- | --- | --- |
| 2.6 | 80 |  |  |  |  |
|  |  | *SxScreen/PCR/ACS* | 1,244 | 3,295,000 | NA |
|  |  | *Hybrid/ACS* | 989 | 3,669,000 | 1,000 |
|  |  | *UniversalPCR/ACS* | 1,682 | 4,176,000 | Dominated |
|  |  | *NoIntervention* | 1,954 | 6,114,000 | Dominated |
|  |  | *Hybrid/Hospital* | 971 | 12,321,000 | Dominated |
|  |  | *SxScreen/PCR/Hospital* | 1,138 | 12,738,000 | Dominated |
|  |  | *UniversalPCR/Hospital* | 1,680 | 12,986,000 | Dominated |
|  |  | *UniversalPCR/TempHousing* | 104 | 39,616,000 | 41,000 |
| 1.3 | 50 |  |  |  |  |
|  |  | *SxScreen/PCR/ACS* | 137 | 409,000 | NA |
|  |  | *Hybrid/ACS* | 103 | 1,341,000 | 27,000 |
|  |  | *UniversalPCR/ACS* | 208 | 1,444,000 | Dominated |
|  |  | *NoIntervention* | 544 | 1,485,000 | Dominated |
|  |  | *SxScreen/PCR/Hospital* | 125 | 1,608,000 | Dominated |
|  |  | *Hybrid/Hospital* | 100 | 2,383,000 | 379,000 |
|  |  | *UniversalPCR/Hospital* | 208 | 2,657,000 | Dominated |
|  |  | *UniversalPCR/TempHousing* | 121 | 39,630,000 | Dominated |
|  | 60 |  |  |  |  |
|  | base case | *SxScreen/PCR/ACS* | 137 | 409,000 | NA |
|  |  | *Hybrid/ACS* | 103 | 1,325,000 | 27,000 |
|  |  | *UniversalPCR/ACS* | 207 | 1,426,000 | Dominated |
|  |  | *NoIntervention* | 538 | 1,461,000 | Dominated |
|  |  | *SxScreen/PCR/Hospital* | 125 | 1,604,000 | Dominated |
|  |  | *Hybrid/Hospital* | 100 | 2,368,000 | 382,000 |
|  |  | *UniversalPCR/Hospital* | 207 | 2,631,000 | Dominated |
|  |  | *UniversalPCR/TempHousing* | 95 | 38,974,000 | 6,854,000 |

**eTable6 continued. One-way sensitivity analysis on efficacy of temporary housing in reducing SARS-CoV-2 transmission.**

| **Effective reproduction number (R_e_)** | **Efficacy of TempHousing in reducing SARS-CoV-2 transmission, %** | **Strategy** | **Cumulative infections, n** | **Total cost *,**  **2020 USD** | **Incr. cost per case prevented *, 2020 USD** |
| --- | --- | --- | --- | --- | --- |
| 1.3 | 70 |  |  |  |  |
|  |  | *SxScreen/PCR/ACS* | 137 | 409,000 | NA |
|  |  | *Hybrid/ACS* | 103 | 1,341,000 | 27,000 |
|  |  | *UniversalPCR/ACS* | 208 | 1,444,000 | Dominated |
|  |  | *NoIntervention* | 544 | 1,485,000 | Dominated |
|  |  | *SxScreen/PCR/Hospital* | 125 | 1,608,000 | Dominated |
|  |  | *Hybrid/Hospital* | 100 | 2,383,000 | 379,000 |
|  |  | *UniversalPCR/Hospital* | 208 | 2,657,000 | Dominated |
|  |  | *UniversalPCR/TempHousing* | 76 | 39,601,000 | 1,529,000 |
|  | 80 |  |  |  |  |
|  |  | *SxScreen/PCR/ACS* | 137 | 409,000 | NA |
|  |  | *Hybrid/ACS* | 103 | 1,341,000 | 27,000 |
|  |  | *UniversalPCR/ACS* | 208 | 1,444,000 | Dominated |
|  |  | *NoIntervention* | 544 | 1,485,000 | Dominated |
|  |  | *SxScreen/PCR/Hospital* | 125 | 1,608,000 | Dominated |
|  |  | *Hybrid/Hospital* | 100 | 2,383,000 | 379,000 |
|  |  | *UniversalPCR/Hospital* | 208 | 2,657,000 | Dominated |
|  |  | *UniversalPCR/TempHousing* | 65 | 39,605,000 | 1,053,000 |
| 0.9 | 50 |  |  |  |  |
|  |  | *SxScreen/PCR/ACS* | 85 | 264,000 | NA |
|  |  | *NoIntervention* | 175 | 543,000 | Dominated |
|  |  | *SxScreen/PCR/Hospital* | 82 | 1,115,000 | Dominated |
|  |  | *UniversalPCR/ACS* | 95 | 1,242,000 | Dominated |
|  |  | *Hybrid/ACS* | 71 | 1,256,000 | 72,000 |
|  |  | *UniversalPCR/Hospital* | 95 | 1,918,000 | Dominated |
|  |  | *Hybrid/Hospital* | 71 | 2,018,000 | Dominated |
|  |  | *UniversalPCR/TempHousing* | 80 | 39,605,000 | Dominated |

**eTable6 continued. One-way sensitivity analysis on efficacy of temporary housing in reducing SARS-CoV-2 transmission.**

| **Effective reproduction number (R_e_)** | **Efficacy of TempHousing in reducing SARS-CoV-2 transmission, %** | **Strategy** | **Cumulative infections, n** | **Total cost *,**  **2020 USD** | **Incr. cost per case prevented *, 2020 USD** |
| --- | --- | --- | --- | --- | --- |
| 0.9 | 60 |  |  |  |  |
|  | base case | *SxScreen/PCR/ACS* | 85 | 264,000 | NA |
|  |  | *NoIntervention* | 174 | 540,000 | Dominated |
|  |  | *SxScreen/PCR/Hospital* | 82 | 1,113,000 | Dominated |
|  |  | *UniversalPCR/ACS* | 94 | 1,226,000 | Dominated |
|  |  | *Hybrid/ACS* | 71 | 1,240,000 | 71,000 |
|  |  | *UniversalPCR/Hospital* | 95 | 1,901,000 | Dominated |
|  |  | *Hybrid/Hospital* | 71 | 2,004,000 | Dominated |
|  |  | *UniversalPCR/TempHousing* | 71 | 38,954,000 | Dominated |
|  | 70 |  |  |  |  |
|  |  | *SxScreen/PCR/ACS* | 85 | 264,000 | NA |
|  |  | *NoIntervention* | 175 | 543,000 | Dominated |
|  |  | *SxScreen/PCR/Hospital* | 82 | 1,115,000 | Dominated |
|  |  | *UniversalPCR/ACS* | 95 | 1,242,000 | Dominated |
|  |  | *Hybrid/ACS* | 71 | 1,256,000 | 72,000 |
|  |  | *UniversalPCR/Hospital* | 95 | 1,918,000 | Dominated |
|  |  | *Hybrid/Hospital* | 71 | 2,018,000 | Dominated |
|  |  | *UniversalPCR/TempHousing* | 63 | 39,600,000 | 4,668,000 |
|  | 80 |  |  |  |  |
|  |  | *SxScreen/PCR/ACS* | 85 | 264,000 | NA |
|  |  | *NoIntervention* | 175 | 543,000 | Dominated |
|  |  | *SxScreen/PCR/Hospital* | 82 | 1,115,000 | Dominated |
|  |  | *UniversalPCR/ACS* | 95 | 1,242,000 | Dominated |
|  |  | *Hybrid/ACS* | 71 | 1,256,000 | 72,000 |
|  |  | *UniversalPCR/Hospital* | 95 | 1,918,000 | Dominated |
|  |  | *Hybrid/Hospital* | 71 | 2,018,000 | Dominated |
|  |  | *UniversalPCR/TempHousing* | 57 | 39,596,000 | 2,697,000 |

Abbreviations: ACS, alternate care sites; COVID-19, coronavirus disease 2019; Dominated, less clinically effective and more costly than an alternative strategy, or a combination of two alternative strategies;^14^ Incr., incremental; *PCR,* polymerase chain reaction; *SxScreen,* symptom screen; *TempHousing,* temporary housing; *UniversalPCR*, universal polymerase chain reaction test for everyone; USD, United States dollars.

Strategies are listed in order of ascending costs, per convention of cost-effectiveness analysis.

* Costs were rounded to the nearest thousands.

**eTable7. One-way sensitivity analysis on cost of a PCR test (n=2,258).**

| **Effective reproduction number (R_e_)** | **PCR test cost, 2020 USD** | **Strategy** | **Cumulative infections, n** | **Total cost *,**  **2020 USD** | **Incr. cost per case prevented *, 2020 USD** |
| --- | --- | --- | --- | --- | --- |
| 2.6 | 5 |  |  |  |  |
|  |  | *Hybrid/ACS* | 989 | 2,711,000 | NA |
|  |  | *UniversalPCR/ACS* | 1,681 | 3,245,000 | Dominated |
|  |  | *SxScreen/PCR/ACS* | 1,239 | 3,246,000 | Dominated |
|  |  | *NoIntervention* | 1,954 | 6,097,000 | Dominated |
|  |  | *Hybrid/Hospital* | 967 | 11,364,000 | Dominated |
|  |  | *UniversalPCR/Hospital* | 1,679 | 12,056,000 | Dominated |
|  |  | *SxScreen/PCR/Hospital* | 1,133 | 12,693,000 | Dominated |
|  |  | *UniversalPCR/TempHousing* | 379 | 38,843,000 | 59,000 |
|  | 10 |  |  |  |  |
|  |  | *Hybrid/ACS* | 985 | 2,815,000 | NA |
|  |  | *SxScreen/PCR/ACS* | 1,239 | 3,251,000 | Dominated |
|  |  | *UniversalPCR/ACS* | 1,681 | 3,346,000 | Dominated |
|  |  | *NoIntervention* | 1,954 | 6,099,000 | Dominated |
|  |  | *Hybrid/Hospital* | 967 | 11,468,000 | Dominated |
|  |  | *UniversalPCR/Hospital* | 1,679 | 12,157,000 | Dominated |
|  |  | *SxScreen/PCR/Hospital* | 1,133 | 12,698,000 | Dominated |
|  |  | *UniversalPCR/TempHousing* | 379 | 38,944,000 | 59,000 |
|  | 25 |  |  |  |  |
|  |  | *Hybrid/ACS* | 985 | 3,127,000 | NA |
|  |  | *SxScreen/PCR/ACS* | 1,239 | 3,267,000 | Dominated |
|  |  | *UniversalPCR/ACS* | 1,681 | 3,650,000 | Dominated |
|  |  | *NoIntervention* | 1,954 | 6,105,000 | Dominated |
|  |  | *Hybrid/Hospital* | 967 | 11,780,000 | Dominated |
|  |  | *UniversalPCR/Hospital* | 1,679 | 12,460,000 | Dominated |
|  |  | *SxScreen/PCR/Hospital* | 1,133 | 12,713,000 | Dominated |
|  |  | *UniversalPCR/TempHousing* | 379 | 39,247,000 | 59,000 |

**eTable7 continued. One-way sensitivity analysis on cost of a PCR test.**

| **Effective reproduction number (R_e_)** | **PCR test cost, 2020 USD** | **Strategy** | **Cumulative infections, n** | **Total cost *,**  **2020 USD** | **Incr. cost per case prevented *, 2020 USD** |
| --- | --- | --- | --- | --- | --- |
| 2.6 | 51 |  |  |  |  |
|  | base case | *SxScreen/PCR/ACS* | 1,239 | 3,267,000 | NA |
|  |  | *Hybrid/ACS* | 985 | 3,628,000 | 1,000 |
|  |  | *UniversalPCR/ACS* | 1,681 | 4,143,000 | Dominated |
|  |  | *NoIntervention* | 1,954 | 6,098,000 | Dominated |
|  |  | *Hybrid/Hospital* | 967 | 12,202,000 | Dominated |
|  |  | *SxScreen/PCR/Hospital* | 1,133 | 12,620,000 | Dominated |
|  |  | *UniversalPCR/Hospital* | 1,679 | 12,914,000 | Dominated |
|  |  | *UniversalPCR/TempHousing* | 376 | 39,119,000 | 58,000 |
|  | 75 |  |  |  |  |
|  |  | *SxScreen/PCR/ACS* | 1,239 | 3,320,000 | NA |
|  |  | *Hybrid/ACS* | 985 | 4,168,000 | 3,000 |
|  |  | *UniversalPCR/ACS* | 1,681 | 4,662,000 | Dominated |
|  |  | *NoIntervention* | 1,954 | 6,123,000 | Dominated |
|  |  | *SxScreen/PCR/Hospital* | 1,133 | 12,761,000 | Dominated |
|  |  | *Hybrid/Hospital* | 967 | 12,821,000 | Dominated |
|  |  | *UniversalPCR/Hospital* | 1,679 | 13,472,000 | Dominated |
|  |  | *UniversalPCR/TempHousing* | 379 | 40,257,000 | 43,000 |
|  | 100 |  |  |  |  |
|  |  | *SxScreen/PCR/ACS* | 1,239 | 3,373,000 | NA |
|  |  | *Hybrid/ACS* | 985 | 4,708,000 | 5,000 |
|  |  | *UniversalPCR/ACS* | 1,681 | 5,181,000 | Dominated |
|  |  | *NoIntervention* | 1,954 | 6,148,000 | Dominated |
|  |  | *Hybrid/Hospital* | 967 | 13,440,000 | Dominated |
|  |  | *SxScreen/PCR/Hospital* | 1,133 | 12,902,000 | Dominated |
|  |  | *UniversalPCR/Hospital* | 1,679 | 14,030,000 | Dominated |
|  |  | *UniversalPCR/TempHousing* | 379 | 40,762,000 | 43,000 |

**eTable7 continued. One-way sensitivity analysis on cost of a PCR test.**

| **Effective reproduction number (R_e_)** | **PCR test cost, 2020 USD** | **Strategy** | **Cumulative infections, n** | **Total cost *,**  **2020 USD** | **Incr. cost per case prevented *, 2020 USD** |
| --- | --- | --- | --- | --- | --- |
| 1.3 | 5 |  |  |  |  |
|  |  | *SxScreen/PCR/ACS* | 137 | 402,000 | NA |
|  |  | *Hybrid/ACS* | 103 | 409,000 | 200 |
|  |  | *UniversalPCR/ACS* | 207 | 515,000 | Dominated |
|  |  | *Hybrid/Hospital* | 100 | 1,451,000 | 379,000 |
|  |  | *NoIntervention* | 538 | 1,480,000 | Dominated |
|  |  | *SxScreen/PCR/Hospital* | 125 | 1,601,000 | Dominated |
|  |  | *UniversalPCR/Hospital* | 207 | 1,728,000 | Dominated |
|  |  | *UniversalPCR/TempHousing* | 95 | 38,696,000 | 6,965,000 |
|  | 10 |  |  |  |  |
|  |  | *SxScreen/PCR/ACS* | 137 | 403,000 | NA |
|  |  | *Hybrid/ACS* | 103 | 510,000 | 3,000 |
|  |  | *UniversalPCR/ACS* | 207 | 616,000 | Dominated |
|  |  | *NoIntervention* | 538 | 1,481,000 | Dominated |
|  |  | *Hybrid/Hospital* | 100 | 1,553,000 | 379,000 |
|  |  | *SxScreen/PCR/Hospital* | 125 | 1,602,000 | Dominated |
|  |  | *UniversalPCR/Hospital* | 207 | 1,829,000 | Dominated |
|  |  | *UniversalPCR/TempHousing* | 95 | 38,797,000 | 6,964,000 |
|  | 25 |  |  |  |  |
|  |  | *SxScreen/PCR/ACS* | 137 | 405,000 | NA |
|  |  | *Hybrid/ACS* | 103 | 814,000 | 12,000 |
|  |  | *UniversalPCR/ACS* | 207 | 919,000 | Dominated |
|  |  | *NoIntervention* | 538 | 1,482,000 | Dominated |
|  |  | *SxScreen/PCR/Hospital* | 125 | 1,604,000 | Dominated |
|  |  | *Hybrid/Hospital* | 100 | 1,856,000 | 379,000 |
|  |  | *UniversalPCR/Hospital* | 207 | 2,132,000 | Dominated |
|  |  | *UniversalPCR/TempHousing* | 95 | 39,101,000 | 6,965,000 |

**eTable7 continued. One-way sensitivity analysis on cost of a PCR test.**

| **Effective reproduction number (R_e_)** | **PCR test cost, 2020 USD** | **Strategy** | **Cumulative infections, n** | **Total cost *,**  **2020 USD** | **Incr. cost per case prevented *, 2020 USD** |
| --- | --- | --- | --- | --- | --- |
| 1.3 | 51 |  |  |  |  |
|  | base case | *SxScreen/PCR/ACS* | 137 | 409,000 | NA |
|  |  | *Hybrid/ACS* | 103 | 1,325,000 | 27,000 |
|  |  | *UniversalPCR/ACS* | 207 | 1,426,000 | Dominated |
|  |  | *NoIntervention* | 538 | 1,461,000 | Dominated |
|  |  | *SxScreen/PCR/Hospital* | 125 | 1,604,000 | Dominated |
|  |  | *Hybrid/Hospital* | 100 | 2,368,000 | 382,000 |
|  |  | *UniversalPCR/Hospital* | 207 | 2,631,000 | Dominated |
|  |  | *UniversalPCR/TempHousing* | 95 | 38,974,000 | 6,854,000 |
|  | 75 |  |  |  |  |
|  |  | *SxScreen/PCR/ACS* | 137 | 413,000 | NA |
|  |  | *NoIntervention* | 538 | 1,487,000 | Dominated |
|  |  | *SxScreen/PCR/Hospital* | 125 | 1,611,000 | Dominated |
|  |  | *Hybrid/ACS* | 103 | 1,827,000 | 41,000 |
|  |  | *UniversalPCR/ACS* | 207 | 1,928,000 | Dominated |
|  |  | *Hybrid/Hospital* | 100 | 2,869,000 | 379,000 |
|  |  | *UniversalPCR/Hospital* | 207 | 3,142,000 | Dominated |
|  |  | *UniversalPCR/TempHousing* | 95 | 40,111,000 | 6,964,000 |
|  | 100 |  |  |  |  |
|  |  | *SxScreen/PCR/ACS* | 137 | 417,000 | NA |
|  |  | *NoIntervention* | 538 | 1,513,000 | Dominated |
|  |  | *SxScreen/PCR/Hospital* | 125 | 1,618,000 | Dominated |
|  |  | *Hybrid/ACS* | 103 | 2,329,000 | 56,000 |
|  |  | *UniversalPCR/ACS* | 207 | 2,430,000 | Dominated |
|  |  | *Hybrid/Hospital* | 100 | 3,370,000 | 347,000 |
|  |  | *UniversalPCR/Hospital* | 207 | 3,653,000 | Dominated |
|  |  | *UniversalPCR/TempHousing* | 95 | 40,616,000 | 7,449,000 |

**eTable7 continued. One-way sensitivity analysis on cost of a PCR test.**

| **Effective reproduction number (R_e_)** | **PCR test cost, 2020 USD** | **Strategy** | **Cumulative infections, n** | **Total cost *,**  **2020 USD** | **Incr. cost per case prevented *, 2020 USD** |
| --- | --- | --- | --- | --- | --- |
| 0.9 | 5 |  |  |  |  |
|  |  | *SxScreen/PCR/ACS* | 85 | 259,000 | NA |
|  |  | *UniversalPCR/ACS* | 95 | 314,000 | Dominated |
|  |  | *Hybrid/ACS* | 71 | 325,000 | 5,000 |
|  |  | *NoIntervention* | 175 | 542,000 | Dominated |
|  |  | *UniversalPCR/Hospital* | 95 | 989,000 | Dominated |
|  |  | *Hybrid/Hospital* | 71 | 1,087,000 | Dominated |
|  |  | *SxScreen/PCR/Hospital* | 82 | 1,110,000 | Dominated |
|  |  | *UniversalPCR/TempHousing* | 71 | 38,677,000 | Dominated |
|  | 10 |  |  |  |  |
|  |  | *SxScreen/PCR/ACS* | 85 | 260,000 | NA |
|  |  | *UniversalPCR/ACS* | 95 | 415,000 | Dominated |
|  |  | *Hybrid/ACS* | 71 | 426,000 | 12,000 |
|  |  | *NoIntervention* | 175 | 542,000 | Dominated |
|  |  | *UniversalPCR/Hospital* | 95 | 1,090,000 | Dominated |
|  |  | *SxScreen/PCR/Hospital* | 82 | 1,111,000 | Dominated |
|  |  | *Hybrid/Hospital* | 71 | 1,189,000 | Dominated |
|  |  | *UniversalPCR/TempHousing* | 71 | 38,778,000 | Dominated |
|  | 25 |  |  |  |  |
|  |  | *SxScreen/PCR/ACS* | 85 | 262,000 | NA |
|  |  | *NoIntervention* | 175 | 542,000 | Dominated |
|  |  | *UniversalPCR/ACS* | 95 | 717,000 | Dominated |
|  |  | *Hybrid/ACS* | 71 | 730,000 | 34,000 |
|  |  | *SxScreen/PCR/Hospital* | 82 | 1,113,000 | Dominated |
|  |  | *UniversalPCR/Hospital* | 95 | 1,393,000 | Dominated |
|  |  | *Hybrid/Hospital* | 71 | 1,492,000 | Dominated |
|  |  | *UniversalPCR/TempHousing* | 71 | 39,081,000 | Dominated |

**eTable7 continued. One-way sensitivity analysis on cost of a PCR test.**

| **Effective reproduction number (R_e_)** | **PCR test cost, 2020 USD** | **Strategy** | **Cumulative infections, n** | **Total cost *,**  **2020 USD** | **Incr. cost per case prevented *, 2020 USD** |
| --- | --- | --- | --- | --- | --- |
| 0.9 | 51 |  |  |  |  |
|  | base case | *SxScreen/PCR/ACS* | 85 | 264,000 | NA |
|  |  | *NoIntervention* | 174 | 540,000 | Dominated |
|  |  | *SxScreen/PCR/Hospital* | 82 | 1,113,000 | Dominated |
|  |  | *UniversalPCR/ACS* | 94 | 1,226,000 | Dominated |
|  |  | *Hybrid/ACS* | 71 | 1,240,000 | 71,000 |
|  |  | *UniversalPCR/Hospital* | 95 | 1,901,000 | Dominated |
|  |  | *Hybrid/Hospital* | 71 | 2,004,000 | Dominated |
|  |  | *UniversalPCR/TempHousing* | 71 | 38,954,000 | Dominated |
|  | 75 |  |  |  |  |
|  |  | *SxScreen/PCR/ACS* | 85 | 267,000 | NA |
|  |  | *NoIntervention* | 175 | 544,000 | Dominated |
|  |  | *SxScreen/PCR/Hospital* | 82 | 1,118,000 | Dominated |
|  |  | *UniversalPCR/ACS* | 95 | 1,727,000 | Dominated |
|  |  | *Hybrid/ACS* | 71 | 1,742,000 | 107,000 |
|  |  | *UniversalPCR/Hospital* | 95 | 2,402,000 | Dominated |
|  |  | *Hybrid/Hospital* | 71 | 2,504,000 | Dominated |
|  |  | *UniversalPCR/TempHousing* | 71 | 40,092,000 | Dominated |
|  | 100 |  |  |  |  |
|  |  | *SxScreen/PCR/ACS* | 85 | 270,000 | NA |
|  |  | *NoIntervention* | 174 | 548,000 | Dominated |
|  |  | *SxScreen/PCR/Hospital* | 82 | 1,123,000 | Dominated |
|  |  | *UniversalPCR/ACS* | 94 | 2,228,000 | Dominated |
|  |  | *Hybrid/ACS* | 71 | 2,244,000 | 141,000 |
|  |  | *UniversalPCR/Hospital* | 95 | 2,903,000 | Dominated |
|  |  | *Hybrid/Hospital* | 71 | 3,004,000 | Dominated |
|  |  | *UniversalPCR/TempHousing* | 71 | 40,597,000 | Dominated |

Abbreviations: ACS, alternate care sites; COVID-19, coronavirus disease 2019; Dominated, less clinically effective and more costly than an alternative strategy, or a combination of two alternative strategies;^14^ Incr., incremental; *PCR,* polymerase chain reaction; *SxScreen,* symptom screen; *TempHousing,* temporary housing; *UniversalPCR*, universal polymerase chain reaction test for everyone; USD, United States dollars.

Strategies are listed in order of ascending costs, per convention of cost-effectiveness analysis.

* Costs were rounded to the nearest thousands.

**eTable8. One-way sensitivity analysis on cost of symptom screen.**

| **Effective reproduction number (R_e_)** | **Cost of symptom screen, 2020 USD** | **Strategy** | **Cumulative infections, n** | **Total cost *,**  **2020 USD** | **Incr. cost per case prevented *, 2020 USD** |
| --- | --- | --- | --- | --- | --- |
| 2.6 | 0 |  |  |  |  |
|  | base case | *SxScreen/PCR/ACS* | 1,239 | 3,267,000 | NA |
|  |  | *Hybrid/ACS* | 985 | 3,628,000 | 1,000 |
|  |  | *UniversalPCR/ACS* | 1,681 | 4,143,000 | Dominated |
|  |  | *NoIntervention* | 1,954 | 6,098,000 | Dominated |
|  |  | *Hybrid/Hospital* | 967 | 12,202,000 | Dominated |
|  |  | *SxScreen/PCR/Hospital* | 1,133 | 12,620,000 | Dominated |
|  |  | *UniversalPCR/Hospital* | 1,679 | 12,914,000 | Dominated |
|  |  | *UniversalPCR/TempHousing* | 376 | 39,119,000 | 58,000 |
|  | 1 |  |  |  |  |
|  |  | *SxScreen/PCR/ACS* | 1,244 | 3,562,000 | NA |
|  |  | *Hybrid/ACS* | 989 | 3,937,000 | 1,000 |
|  |  | *UniversalPCR/ACS* | 1,682 | 4,176,000 | Dominated |
|  |  | *NoIntervention* | 1,954 | 6,114,000 | Dominated |
|  |  | *Hybrid/Hospital* | 971 | 12,590,000 | Dominated |
|  |  | *UniversalPCR/Hospital* | 1,680 | 12,986,000 | Dominated |
|  |  | *SxScreen/PCR/Hospital* | 1,138 | 13,006,000 | Dominated |
|  |  | *UniversalPCR/TempHousing* | 379 | 39,772,000 | 59,000 |
|  | 5 |  |  |  |  |
|  |  | *UniversalPCR/ACS* | 1,682 | 4,176,000 | NA |
|  |  | *SxScreen/PCR/ACS* | 1,244 | 4,629,000 | 1,000 |
|  |  | *Hybrid/ACS* | 989 | 5,011,000 | 2,000 |
|  |  | *NoIntervention* | 1,954 | 6,114,000 | Dominated |
|  |  | *UniversalPCR/Hospital* | 1,680 | 12,986,000 | Dominated |
|  |  | *Hybrid/Hospital* | 971 | 13,664,000 | Dominated |
|  |  | *SxScreen/PCR/Hospital* | 1,138 | 14,077,000 | Dominated |
|  |  | *UniversalPCR/TempHousing* | 379 | 39,772,000 | 57,000 |

**eTable8 continued. One-way sensitivity analysis on cost of symptom screen.**

| **Effective reproduction number (R_e_)** | **Cost of symptom screen, 2020 USD** | **Strategy** | **Cumulative infections, n** | **Total cost *,**  **2020 USD** | **Incr. cost per case prevented *, 2020 USD** |
| --- | --- | --- | --- | --- | --- |
| 1.3 | 0 |  |  |  |  |
|  | base case | *SxScreen/PCR/ACS* | 137 | 409,000 | NA |
|  |  | *Hybrid/ACS* | 103 | 1,325,000 | 27,000 |
|  |  | *UniversalPCR/ACS* | 207 | 1,426,000 | Dominated |
|  |  | *NoIntervention* | 538 | 1,461,000 | Dominated |
|  |  | *SxScreen/PCR/Hospital* | 125 | 1,604,000 | Dominated |
|  |  | *Hybrid/Hospital* | 100 | 2,368,000 | 382,000 |
|  |  | *UniversalPCR/Hospital* | 207 | 2,631,000 | Dominated |
|  |  | *UniversalPCR/TempHousing* | 95 | 38,974,000 | 6,854,000 |
|  | 1 |  |  |  |  |
|  |  | *SxScreen/PCR/ACS* | 137 | 684,000 | NA |
|  |  | *UniversalPCR/ACS* | 208 | 1,444,000 | Dominated |
|  |  | *NoIntervention* | 544 | 1,485,000 | Dominated |
|  |  | *Hybrid/ACS* | 103 | 1,616,000 | 27,000 |
|  |  | *SxScreen/PCR/Hospital* | 125 | 1,882,000 | Dominated |
|  |  | *UniversalPCR/Hospital* | 208 | 2,657,000 | Dominated |
|  |  | *Hybrid/Hospital* | 100 | 2,658,000 | 379,000 |
|  |  | *UniversalPCR/TempHousing* | 95 | 39,626,000 | 6,913,000 |
|  | 5 |  |  |  |  |
|  |  | *UniversalPCR/ACS* | 208 | 1,444,000 | NA |
|  |  | *NoIntervention* | 544 | 1,485,000 | Dominated |
|  |  | *SxScreen/PCR/ACS* | 137 | 1,781,000 | 5,000 |
|  |  | *UniversalPCR/Hospital* | 208 | 2,657,000 | Dominated |
|  |  | *Hybrid/ACS* | 103 | 2,714,000 | 27,000 |
|  |  | *SxScreen/PCR/Hospital* | 125 | 2,980,000 | Dominated |
|  |  | *Hybrid/Hospital* | 100 | 3,756,000 | 379,000 |
|  |  | *UniversalPCR/TempHousing* | 95 | 39,626,000 | 6,707,000 |

**eTable8 continued. One-way sensitivity analysis on cost of symptom screen.**

| **Effective reproduction number (R_e_)** | **Cost of symptom screen, 2020 USD** | **Strategy** | **Cumulative infections, n** | **Total cost *,**  **2020 USD** | **Incr. cost per case prevented *, 2020 USD** |
| --- | --- | --- | --- | --- | --- |
| 0.9 | 0 |  |  |  |  |
|  | base case | *SxScreen/PCR/ACS* | 85 | 264,000 | NA |
|  |  | *NoIntervention* | 174 | 540,000 | Dominated |
|  |  | *SxScreen/PCR/Hospital* | 82 | 1,113,000 | Dominated |
|  |  | *UniversalPCR/ACS* | 94 | 1,226,000 | Dominated |
|  |  | *Hybrid/ACS* | 71 | 1,240,000 | 71,000 |
|  |  | *UniversalPCR/Hospital* | 95 | 1,901,000 | Dominated |
|  |  | *Hybrid/Hospital* | 71 | 2,004,000 | Dominated |
|  |  | *UniversalPCR/TempHousing* | 71 | 38,954,000 | Dominated |
|  | 1 |  |  |  |  |
|  |  | *SxScreen/PCR/ACS* | 85 | 539,000 | NA |
|  |  | *NoIntervention* | 175 | 543,000 | Dominated |
|  |  | *UniversalPCR/ACS* | 95 | 1,242,000 | Dominated |
|  |  | *SxScreen/PCR/Hospital* | 82 | 1,390,000 | Dominated |
|  |  | *Hybrid/ACS* | 71 | 1,531,000 | 72,000 |
|  |  | *UniversalPCR/Hospital* | 95 | 1,918,000 | Dominated |
|  |  | *Hybrid/Hospital* | 71 | 2,293,000 | Dominated |
|  |  | *UniversalPCR/TempHousing* | 71 | 39,607,000 | Dominated |
|  | 5 |  |  |  |  |
|  |  | *NoIntervention* | 175 | 543,000 | NA |
|  |  | *UniversalPCR/ACS* | 95 | 1,242,000 | 9,000 |
|  |  | *SxScreen/PCR/ACS* | 85 | 1,638,000 | 42,000 |
|  |  | *UniversalPCR/Hospital* | 95 | 1,918,000 | Dominated |
|  |  | *SxScreen/PCR/Hospital* | 82 | 2,489,000 | Dominated |
|  |  | *Hybrid/ACS* | 71 | 2,630,000 | 72,000 |
|  |  | *Hybrid/Hospital* | 71 | 3,393,000 | Dominated |
|  |  | *UniversalPCR/TempHousing* | 71 | 39,607,000 | Dominated |

Abbreviations: ACS, alternate care sites; COVID-19, coronavirus disease 2019; Dominated, less clinically effective and more costly than an alternative strategy, or a combination of two alternative strategies;^14^ Incr., incremental; *PCR,* polymerase chain reaction; *SxScreen,* symptom screen; *TempHousing,* temporary housing; *UniversalPCR*, universal polymerase chain reaction test for everyone; USD, United States dollars.

Strategies are listed in order of ascending costs, per convention of cost-effectiveness analysis.

* Costs were rounded to the nearest thousands.

**eTable9. One-way sensitivity analysis on daily costs of hospital beds.**

| **Effective reproduction number (R_e_)** | **Daily costs of hospital beds** | **Strategy** | **Cumulative infections, n** | **Total cost *,**  **2020 USD** | **Incr. cost per case prevented *, 2020 USD** |
| --- | --- | --- | --- | --- | --- |
| 2.6 | 0.5X base case |  |  |  |  |
|  |  | *SxScreen/PCR/ACS* | 1,239 | 2,903,000 | NA |
|  |  | *NoIntervention* | 1,954 | 3,065,000 | Dominated |
|  |  | *Hybrid/ACS* | 985 | 3,365,000 | 2,000 |
|  |  | *UniversalPCR/ACS* | 1,681 | 3,608,000 | Dominated |
|  |  | *SxScreen/PCR/Hospital* | 1,133 | 6,390,000 | Dominated |
|  |  | *Hybrid/Hospital* | 967 | 6,688,000 | Dominated |
|  |  | *UniversalPCR/Hospital* | 1,680 | 7,006,000 | Dominated |
|  |  | *UniversalPCR/TempHousing* | 376 | 39,650,000 | 59,000 |
|  | Base case |  |  |  |  |
|  |  | *SxScreen/PCR/ACS* | 1,239 | 3,267,000 | NA |
|  |  | *Hybrid/ACS* | 985 | 3,628,000 | 1,000 |
|  |  | *UniversalPCR/ACS* | 1,681 | 4,143,000 | Dominated |
|  |  | *NoIntervention* | 1,954 | 6,098,000 | Dominated |
|  |  | *Hybrid/Hospital* | 967 | 12,202,000 | Dominated |
|  |  | *SxScreen/PCR/Hospital* | 1,133 | 12,620,000 | Dominated |
|  |  | *UniversalPCR/Hospital* | 1,679 | 12,914,000 | Dominated |
|  |  | *UniversalPCR/TempHousing* | 376 | 39,119,000 | 58,000 |
|  | 1.5X base case |  |  |  |  |
|  |  | *SxScreen/PCR/ACS* | 1,239 | 3,687,000 | NA |
|  |  | *Hybrid/ACS* | 985 | 3,972,000 | 1,000 |
|  |  | *UniversalPCR/ACS* | 1,681 | 4,744,000 | Dominated |
|  |  | *NoIntervention* | 1,954 | 9,164,000 | Dominated |
|  |  | *Hybrid/Hospital* | 967 | 17,955,000 | Dominated |
|  |  | *UniversalPCR/Hospital* | 1,680 | 18,967,000 | Dominated |
|  |  | *SxScreen/PCR/Hospital* | 1,138 | 19,086,000 | Dominated |
|  |  | *UniversalPCR/TempHousing* | 376 | 39,894,000 | 59,000 |

**eTable9 continued. One-way sensitivity analysis on daily costs of hospital beds.**

| **Effective reproduction number (R_e_)** | **Daily costs of hospital beds** | **Strategy** | **Cumulative infections, n** | **Total cost *,**  **2020 USD** | **Incr. cost per case prevented *, 2020 USD** |
| --- | --- | --- | --- | --- | --- |
| 1.3 | 0.5X base case |  |  |  |  |
|  |  | *SxScreen/PCR/ACS* | 137 | 361,000 | NA |
|  |  | *NoIntervention* | 544 | 744,000 | Dominated |
|  |  | *SxScreen/PCR/Hospital* | 125 | 807,000 | Dominated |
|  |  | *Hybrid/ACS* | 103 | 1,307,000 | 28,000 |
|  |  | *UniversalPCR/ACS* | 208 | 1,375,000 | Dominated |
|  |  | *Hybrid/Hospital* | 100 | 1,708,000 | 146,000 |
|  |  | *UniversalPCR/Hospital* | 208 | 1,843,000 | Dominated |
|  |  | *UniversalPCR/TempHousing* | 95 | 39,586,000 | 7,083,000 |
|  | Base case |  |  |  |  |
|  |  | *SxScreen/PCR/ACS* | 137 | 409,000 | NA |
|  |  | *Hybrid/ACS* | 103 | 1,325,000 | 27,000 |
|  |  | *UniversalPCR/ACS* | 207 | 1,426,000 | Dominated |
|  |  | *No Intervention* | 538 | 1,461,000 | Dominated |
|  |  | *SxScreen/Hospital* | 125 | 1,604,000 | Dominated |
|  |  | *Hybrid/Hospital* | 100 | 2,368,000 | 382,000 |
|  |  | *UniversalPCR/Hospital* | 207 | 2,631,000 | Dominated |
|  |  | *UniversalPCR/TempHousing* | 95 | 38,974,000 | 6,854,000 |
|  | 1.5X base case |  |  |  |  |
|  |  | *SxScreen/PCR/ACS* | 137 | 458,000 | NA |
|  |  | *Hybrid/ACS* | 103 | 1,375,000 | 27,000 |
|  |  | *UniversalPCR/ACS* | 208 | 1,512,000 | Dominated |
|  |  | *NoIntervention* | 544 | 2,225,000 | Dominated |
|  |  | *SxScreen/PCR/Hospital* | 125 | 2,408,000 | Dominated |
|  |  | *Hybrid/Hospital* | 100 | 3,058,000 | 611,000 |
|  |  | *UniversalPCR/Hospital* | 208 | 3,471,000 | Dominated |
|  |  | *UniversalPCR/TempHousing* | 95 | 39,666,000 | 6,845,000 |

**eTable9 continued. One-way sensitivity analysis on daily costs of hospital beds.**

| **Effective reproduction number (R_e_)** | **Daily costs of hospital beds** | **Strategy** | **Cumulative infections, n** | **Total cost *,**  **2020 USD** | **Incr. cost per case prevented *, 2020 USD** |
| --- | --- | --- | --- | --- | --- |
| 0.9 | 0.5X base case |  |  |  |  |
|  |  | *SxScreen/PCR/ACS* | 85 | 234,000 | NA |
|  |  | *NoIntervention* | 174 | 272,000 | Dominated |
|  |  | *SxScreen/PCR/Hospital* | 82 | 560,000 | Dominated |
|  |  | *UniversalPCR/ACS* | 94 | 1,211,000 | Dominated |
|  |  | *Hybrid/ACS* | 71 | 1,230,000 | 72,000 |
|  |  | *UniversalPCR/Hospital* | 95 | 1,473,000 | Dominated |
|  |  | *Hybrid/Hospital* | 71 | 1,525,000 | Dominated |
|  |  | *UniversalPCR/TempHousing* | 71 | 39,578,000 | 178,311,000 |
|  | Base case |  |  |  |  |
|  |  | *SxScreen/PCR/ACS* | 85 | 264,000 | NA |
|  |  | *NoIntervention* | 174 | 540,000 | Dominated |
|  |  | *SxScreen/PCR/Hospital* | 82 | 1,113,000 | Dominated |
|  |  | *UniversalPCR/ACS* | 94 | 1,226,000 | Dominated |
|  |  | *Hybrid/ACS* | 71 | 1,240,000 | 71,000 |
|  |  | *UniversalPCR/Hospital* | 95 | 1,901,000 | Dominated |
|  |  | *Hybrid/Hospital* | 71 | 2,004,000 | Dominated |
|  |  | *UniversalPCR/TempHousing* | 71 | 38,954,000 | Dominated |
|  | 1.5X base case |  |  |  |  |
|  |  | *SxScreen/PCR/ACS* | 85 | 295,000 | NA |
|  |  | *NoIntervention* | 174 | 814,000 | Dominated |
|  |  | *UniversalPCR/ACS* | 82 | 1,273,000 | Dominated |
|  |  | *Hybrid/ACS* | 94 | 1,282,000 | 71,000 |
|  |  | *SxScreen/PCR/Hospital* | 71 | 1,671,000 | Dominated |
|  |  | *UniversalPCR/Hospital* | 95 | 2,362,000 | Dominated |
|  |  | *Hybrid/Hospital* | 71 | 2,512,000 | Dominated |
|  |  | *UniversalPCR/TempHousing* | 71 | 39,636,000 | 178,339,000 |

Abbreviations: ACS, alternate care sites; COVID-19, coronavirus disease 2019; Dominated, less clinically effective and more costly than an alternative strategy, or a combination of two alternative strategies;^14^ Incr., incremental; *PCR,* polymerase chain reaction; *SxScreen,* symptom screen; *TempHousing,* temporary housing; *UniversalPCR*, universal polymerase chain reaction test for everyone; USD, United States dollars.

Strategies are listed in order of ascending costs, per convention of cost-effectiveness analysis.

* Costs were rounded to the nearest thousands.

**eTable10. One-way sensitivity analysis on daily cost of an ACS.**

| **Effective reproduction number (R_e_)** | **Daily cost of an ACS, 2020 USD** | **Strategy** | **Cumulative infections, n** | **Total cost *,**  **2020 USD** | **Incr. cost per case prevented *, 2020 USD** |
| --- | --- | --- | --- | --- | --- |
| 2.6 | 150 |  |  |  |  |
|  |  | *SxScreen/PCR/ACS* | 1,244 | 2,050,000 | NA |
|  |  | *Hybrid/ACS* | 989 | 2,656,000 | 2,000 |
|  |  | *UniversalPCR/ACS* | 1,682 | 3,159,000 | Dominated |
|  |  | *NoIntervention* | 1,954 | 6,114,000 | Dominated |
|  |  | *Hybrid/Hospital* | 971 | 12,321,000 | Dominated |
|  |  | *SxScreen/PCR/Hospital* | 1,138 | 12,738,000 | Dominated |
|  |  | *UniversalPCR/Hospital* | 1,680 | 12,986,000 | Dominated |
|  |  | *UniversalPCR/TempHousing* | 379 | 39,772,000 | 61,000 |
|  | 300 |  |  |  |  |
|  | base case | *SxScreen/PCR/ACS* | 1,244 | 3,262,000 | NA |
|  |  | *Hybrid/ACS* | 989 | 3,642,000 | 1,000 |
|  |  | *UniversalPCR/ACS* | 1,682 | 4,150,000 | Dominated |
|  |  | *NoIntervention* | 1,954 | 6,114,000 | Dominated |
|  |  | *Hybrid/Hospital* | 971 | 12,321,000 | Dominated |
|  |  | *SxScreen/PCR/Hospital* | 1,138 | 12,738,000 | Dominated |
|  |  | *UniversalPCR/Hospital* | 1,680 | 12,986,000 | Dominated |
|  |  | *UniversalPCR/TempHousing* | 376 | 39,119,000 | 58,000 |
|  | 450 |  |  |  |  |
|  |  | *SxScreen/PCR/ACS* | 1,244 | 4,475,000 | NA |
|  |  | *Hybrid/ACS* | 989 | 4,629,000 | 1,000 |
|  |  | *UniversalPCR/ACS* | 1,682 | 5,141,000 | Dominated |
|  |  | *NoIntervention* | 1,954 | 6,114,000 | Dominated |
|  |  | *Hybrid/Hospital* | 971 | 12,321,000 | Dominated |
|  |  | *SxScreen/PCR/Hospital* | 1,138 | 12,738,000 | Dominated |
|  |  | *UniversalPCR/Hospital* | 1,680 | 12,986,000 | Dominated |
|  |  | *UniversalPCR/TempHousing* | 379 | 39,772,000 | 58,000 |

**eTable10 continued. One-way sensitivity analysis on daily cost of an ACS.**

| **Effective reproduction number (R_e_)** | **Daily cost of an ACS, 2020 USD** | **Strategy** | **Cumulative infections, n** | **Total cost *,**  **2020 USD** | **Incr. cost per case prevented *, 2020 USD** |
| --- | --- | --- | --- | --- | --- |
| 1.3 | 150 |  |  |  |  |
|  |  | *SxScreen/PCR/ACS* | 137 | 255,000 | NA |
|  |  | *Hybrid/ACS* | 103 | 1,219,000 | 28,000 |
|  |  | *UniversalPCR/ACS* | 208 | 1,304,000 | Dominated |
|  |  | *NoIntervention* | 544 | 1,485,000 | Dominated |
|  |  | *SxScreen/PCR/Hospital* | 125 | 1,608,000 | Dominated |
|  |  | *Hybrid/Hospital* | 100 | 2,383,000 | 423,000 |
|  |  | *UniversalPCR/Hospital* | 208 | 2,657,000 | Dominated |
|  |  | *UniversalPCR/TempHousing* | 95 | 39,626,000 | 6,964,000 |
|  | 300 |  |  |  |  |
|  | base case | *SxScreen/PCR/ACS* | 137 | 405,000 | NA |
|  |  | *Hybrid/ACS* | 103 | 1,338,000 | 27,000 |
|  |  | *UniversalPCR/ACS* | 208 | 1,440,000 | Dominated |
|  |  | *NoIntervention* | 544 | 1,485,000 | Dominated |
|  |  | *SxScreen/PCR/Hospital* | 125 | 1,608,000 | Dominated |
|  |  | *Hybrid/Hospital* | 100 | 2,383,000 | 380,000 |
|  |  | *UniversalPCR/Hospital* | 208 | 2,657,000 | Dominated |
|  |  | *UniversalPCR/TempHousing* | 95 | 38,974,000 | 6,854,000 |
|  | 450 |  |  |  |  |
|  |  | *SxScreen/PCR/ACS* | 137 | 555,000 | NA |
|  |  | *Hybrid/ACS* | 103 | 1,456,000 | 26,000 |
|  |  | *NoIntervention* | 544 | 1,485,000 | Dominated |
|  |  | *UniversalPCR/ACS* | 208 | 1,577,000 | Dominated |
|  |  | *SxScreen/PCR/Hospital* | 125 | 1,608,000 | Dominated |
|  |  | *Hybrid/Hospital* | 100 | 2,383,000 | 337,000 |
|  |  | *UniversalPCR/Hospital* | 208 | 2,657,000 | Dominated |
|  |  | *UniversalPCR/TempHousing* | 95 | 39,626,000 | 6,964,000 |

**eTable10 continued. One-way sensitivity analysis on daily cost of an ACS.**

| **Effective reproduction number (R_e_)** | **Daily cost of an ACS, 2020 USD** | **Strategy** | **Cumulative infections, n** | **Total cost *,**  **2020 USD** | **Incr. cost per case prevented *, 2020 USD** |
| --- | --- | --- | --- | --- | --- |
| 0.9 | 150 |  |  |  |  |
|  |  | *SxScreen/PCR/ACS* | 85 | 164,000 | NA |
|  |  | *NoIntervention* | 175 | 543,000 | Dominated |
|  |  | *SxScreen/PCR/Hospital* | 82 | 1,115,000 | Dominated |
|  |  | *UniversalPCR/ACS* | 95 | 1,166,000 | Dominated |
|  |  | *Hybrid/ACS* | 71 | 1,168,000 | 73,000 |
|  |  | *UniversalPCR/Hospital* | 95 | 1,918,000 | Dominated |
|  |  | *Hybrid/Hospital* | 71 | 2,018,000 | Dominated |
|  |  | *UniversalPCR/TempHousing* | 71 | 39,607,000 | Dominated |
|  | 300 |  |  |  |  |
|  | base case | *SxScreen/PCR/ACS* | 85 | 262,000 | NA |
|  |  | *NoIntervention* | 175 | 543,000 | Dominated |
|  |  | *SxScreen/PCR/Hospital* | 82 | 1,115,000 | Dominated |
|  |  | *UniversalPCR/ACS* | 95 | 1,240,000 | Dominated |
|  |  | *Hybrid/ACS* | 71 | 1,254,000 | 72,000 |
|  |  | *UniversalPCR/Hospital* | 95 | 1,918,000 | Dominated |
|  |  | *Hybrid/Hospital* | 71 | 2,018,000 | Dominated |
|  |  | *UniversalPCR/TempHousing* | 71 | 38,954,000 | Dominated |
|  | 450 |  |  |  |  |
|  |  | *SxScreen/PCR/ACS* | 85 | 360,000 | NA |
|  |  | *NoIntervention* | 175 | 543,000 | Dominated |
|  |  | *SxScreen/PCR/Hospital* | 82 | 1,115,000 | Dominated |
|  |  | *UniversalPCR/ACS* | 95 | 1,314,000 | Dominated |
|  |  | *Hybrid/ACS* | 71 | 1,339,000 | 71,000 |
|  |  | *UniversalPCR/Hospital* | 95 | 1,918,000 | Dominated |
|  |  | *Hybrid/Hospital* | 71 | 2,018,000 | Dominated |
|  |  | *UniversalPCR/TempHousing* | 71 | 39,607,000 | Dominated |

Abbreviations: ACS, alternate care sites; COVID-19, coronavirus disease 2019; Dominated, less clinically effective and more costly than an alternative strategy, or a combination of two alternative strategies;^14^ Incr., incremental; *PCR,* polymerase chain reaction; *SxScreen,* symptom screen; *TempHousing,* temporary housing; *UniversalPCR*, universal polymerase chain reaction test for everyone; USD, United States dollars.

Strategies are listed in order of ascending costs, per convention of cost-effectiveness analysis.

* Costs were rounded to the nearest thousands.

**eTable11. One-way sensitivity analysis on daily cost of temporary housing.**

| **Effective reproduction number (R_e_)** | **Daily cost of temporary housing, 2020 USD** | **Strategy** | **Cumulative infections, n** | **Total cost *,**  **2020 USD** | **Incr. cost per case prevented *, 2020 USD** |
| --- | --- | --- | --- | --- | --- |
| 2.6 | 0 |  |  |  |  |
|  |  | *UniversalPCR/TempHousing* | 379 | 1,274,000 | NA |
|  |  | *SxScreen/PCR/ACS* | 1,239 | 3,267,000 | Dominated |
|  |  | *Hybrid/ACS* | 985 | 3,628,000 | Dominated |
|  |  | *UniversalPCR/ACS* | 1,681 | 4,143,000 | Dominated |
|  |  | *NoIntervention* | 1,954 | 6,098,000 | Dominated |
|  |  | *Hybrid/Hospital* | 967 | 12,202,000 | Dominated |
|  |  | *SxScreen/PCR/Hospital* | 1,133 | 12,620,000 | Dominated |
|  |  | *UniversalPCR/Hospital* | 1,679 | 12,914,000 | Dominated |
|  | 20 |  |  |  |  |
|  |  | *SxScreen/PCR/ACS* | 1,239 | 3,267,000 | NA |
|  |  | *Hybrid/ACS* | 985 | 3,628,000 | 1,000 |
|  |  | *UniversalPCR/ACS* | 1,681 | 4,143,000 | Dominated |
|  |  | *NoIntervention* | 1,954 | 6,098,000 | Dominated |
|  |  | *UniversalPCR/TempHousing* | 379 | 6,735,000 | 5,000 |
|  |  | *Hybrid/Hospital* | 967 | 12,202,000 | Dominated |
|  |  | *SxScreen/PCR/Hospital* | 1,133 | 12,620,000 | Dominated |
|  |  | *UniversalPCR/Hospital* | 1,679 | 12,914,000 | Dominated |
|  | 40 |  |  |  |  |
|  |  | *SxScreen/PCR/ACS* | 1,239 | 3,267,000 | NA |
|  |  | *Hybrid/ACS* | 985 | 3,628,000 | 1,000 |
|  |  | *UniversalPCR/ACS* | 1,681 | 4,143,000 | Dominated |
|  |  | *NoIntervention* | 1,954 | 6,098,000 | Dominated |
|  |  | *UniversalPCR/TempHousing* | 379 | 12,196,000 | 14,000 |
|  |  | *Hybrid/Hospital* | 967 | 12,202,000 | Dominated |
|  |  | *SxScreen/PCR/Hospital* | 1,133 | 12,620,000 | Dominated |
|  |  | *UniversalPCR/Hospital* | 1,679 | 12,914,000 | Dominated |

**eTable11 continued. One-way sensitivity analysis on daily cost of temporary housing.**

| **Effective reproduction number (R_e_)** | **Daily cost of temporary housing, 2020 USD** | **Strategy** | **Cumulative infections, n** | **Total cost *,**  **2020 USD** | **Incr. cost per case prevented *, 2020 USD** |
| --- | --- | --- | --- | --- | --- |
| 2.6 | 60 |  |  |  |  |
|  |  | *SxScreen/PCR/ACS* | 1,239 | 3,267,000 | NA |
|  |  | *Hybrid/ACS* | 985 | 3,628,000 | 1,000 |
|  |  | *UniversalPCR/ACS* | 1,681 | 4,143,000 | Dominated |
|  |  | *NoIntervention* | 1,954 | 6,098,000 | Dominated |
|  |  | *Hybrid/Hospital* | 967 | 12,202,000 | Dominated |
|  |  | *SxScreen/PCR/Hospital* | 1,133 | 12,620,000 | Dominated |
|  |  | *UniversalPCR/Hospital* | 1,679 | 12,914,000 | Dominated |
|  |  | *UniversalPCR/TempHousing* | 379 | 17,657,000 | 23,000 |
|  | 80 |  |  |  |  |
|  |  | *SxScreen/PCR/ACS* | 1,239 | 3,267,000 | NA |
|  |  | *Hybrid/ACS* | 985 | 3,628,000 | 1,000 |
|  |  | *UniversalPCR/ACS* | 1,681 | 4,143,000 | Dominated |
|  |  | *NoIntervention* | 1,954 | 6,098,000 | Dominated |
|  |  | *Hybrid/Hospital* | 967 | 12,202,000 | Dominated |
|  |  | *SxScreen/PCR/Hospital* | 1,133 | 12,620,000 | Dominated |
|  |  | *UniversalPCR/Hospital* | 1,679 | 12,914,000 | Dominated |
|  |  | *UniversalPCR/TempHousing* | 379 | 23,117,000 | 32,000 |
|  | 100 |  |  |  |  |
|  |  | *SxScreen/PCR/ACS* | 1,239 | 3,267,000 | NA |
|  |  | *Hybrid/ACS* | 985 | 3,628,000 | 1,000 |
|  |  | *UniversalPCR/ACS* | 1,681 | 4,143,000 | Dominated |
|  |  | *NoIntervention* | 1,954 | 6,098,000 | Dominated |
|  |  | *Hybrid/Hospital* | 967 | 12,202,000 | Dominated |
|  |  | *SxScreen/PCR/Hospital* | 1,133 | 12,620,000 | Dominated |
|  |  | *UniversalPCR/Hospital* | 1,679 | 12,914,000 | Dominated |
|  |  | *UniversalPCR/TempHousing* | 379 | 28,578,000 | 41,000 |

**eTable11 continued. One-way sensitivity analysis on daily cost of temporary housing.**

| **Effective reproduction number (R_e_)** | **Daily cost of temporary housing, 2020 USD** | **Strategy** | **Cumulative infections, n** | **Total cost *,**  **2020 USD** | **Incr. cost per case prevented *, 2020 USD** |
| --- | --- | --- | --- | --- | --- |
| 2.6 | 120 |  |  |  |  |
|  |  | *SxScreen/PCR/ACS* | 1,239 | 3,267,000 | NA |
|  |  | *Hybrid/ACS* | 985 | 3,628,000 | 1,000 |
|  |  | *UniversalPCR/ACS* | 1,681 | 4,143,000 | Dominated |
|  |  | *NoIntervention* | 1,954 | 6,098,000 | Dominated |
|  |  | *Hybrid/Hospital* | 967 | 12,202,000 | Dominated |
|  |  | *SxScreen/PCR/Hospital* | 1,133 | 12,620,000 | Dominated |
|  |  | *UniversalPCR/Hospital* | 1,679 | 12,914,000 | Dominated |
|  |  | *UniversalPCR/TempHousing* | 379 | 34,039,000 | 50,000 |
|  | 141 |  |  |  |  |
|  | base case | *SxScreen/PCR/ACS* | 1,239 | 3,267,000 | NA |
|  |  | *Hybrid/ACS* | 985 | 3,628,000 | 1,000 |
|  |  | *UniversalPCR/ACS* | 1,681 | 4,143,000 | Dominated |
|  |  | *NoIntervention* | 1,954 | 6,098,000 | Dominated |
|  |  | *Hybrid/Hospital* | 967 | 12,202,000 | Dominated |
|  |  | *SxScreen/PCR/Hospital* | 1,133 | 12,620,000 | Dominated |
|  |  | *UniversalPCR/Hospital* | 1,679 | 12,914,000 | Dominated |
|  |  | *UniversalPCR/TempHousing* | 376 | 39,119,000 | 58,000 |
|  | 160 |  |  |  |  |
|  |  | *SxScreen/PCR/ACS* | 1,239 | 3,267,000 | NA |
|  |  | *Hybrid/ACS* | 985 | 3,628,000 | 1,000 |
|  |  | *UniversalPCR/ACS* | 1,681 | 4,143,000 | Dominated |
|  |  | *NoIntervention* | 1,954 | 6,098,000 | Dominated |
|  |  | *Hybrid/Hospital* | 967 | 12,202,000 | Dominated |
|  |  | *SxScreen/PCR/Hospital* | 1,133 | 12,620,000 | Dominated |
|  |  | *UniversalPCR/Hospital* | 1,679 | 12,914,000 | Dominated |
|  |  | *UniversalPCR/TempHousing* | 379 | 44,960,000 | 68,000 |

**eTable11 continued. One-way sensitivity analysis on daily cost of temporary housing.**

| **Effective reproduction number (R_e_)** | **Daily cost of temporary housing, 2020 USD** | **Strategy** | **Cumulative infections, n** | **Total cost *,**  **2020 USD** | **Incr. cost per case prevented *, 2020 USD** |
| --- | --- | --- | --- | --- | --- |
| 1.3 | 0 |  |  |  |  |
|  |  | *SxScreen/PCR/ACS* | 137 | 409,000 | NA |
|  |  | *UniversalPCR/TempHousing* | 95 | 1,110,000 | 17,000 |
|  |  | *Hybrid/ACS* | 103 | 1,325,000 | Dominated |
|  |  | *UniversalPCR/ACS* | 207 | 1,426,000 | Dominated |
|  |  | *NoIntervention* | 538 | 1,461,000 | Dominated |
|  |  | *SxScreen/PCR/Hospital* | 125 | 1,604,000 | Dominated |
|  |  | *Hybrid/Hospital* | 100 | 2,368,000 | Dominated |
|  |  | *UniversalPCR/Hospital* | 207 | 2,631,000 | Dominated |
|  | 20 |  |  |  |  |
|  |  | *SxScreen/PCR/ACS* | 137 | 409,000 | NA |
|  |  | *Hybrid/ACS* | 103 | 1,325,000 | 27,000 |
|  |  | *UniversalPCR/ACS* | 207 | 1,426,000 | Dominated |
|  |  | *NoIntervention* | 538 | 1,461,000 | Dominated |
|  |  | *SxScreen/PCR/Hospital* | 125 | 1,604,000 | Dominated |
|  |  | *Hybrid/Hospital* | 100 | 2,368,000 | 348,000 |
|  |  | *UniversalPCR/Hospital* | 207 | 2,631,000 | Dominated |
|  |  | *UniversalPCR/TempHousing* | 95 | 6,574,000 | 896,000 |
|  | 40 |  |  |  |  |
|  |  | *SxScreen/PCR/ACS* | 137 | 409,000 | NA |
|  |  | *Hybrid/ACS* | 103 | 1,325,000 | 27,000 |
|  |  | *UniversalPCR/ACS* | 207 | 1,426,000 | Dominated |
|  |  | *NoIntervention* | 538 | 1,461,000 | Dominated |
|  |  | *SxScreen/PCR/Hospital* | 125 | 1,604,000 | Dominated |
|  |  | *Hybrid/Hospital* | 100 | 2,368,000 | 348,000 |
|  |  | *UniversalPCR/Hospital* | 207 | 2,631,000 | Dominated |
|  |  | *UniversalPCR/TempHousing* | 95 | 12,037,000 | 2,060,000 |

**eTable11 continued. One-way sensitivity analysis on daily cost of temporary housing.**

| **Effective reproduction number (R_e_)** | **Daily cost of temporary housing, 2020 USD** | **Strategy** | **Cumulative infections, n** | **Total cost *,**  **2020 USD** | **Incr. cost per case prevented *, 2020 USD** |
| --- | --- | --- | --- | --- | --- |
| 1.3 | 60 |  |  |  |  |
|  |  | *SxScreen/PCR/ACS* | 137 | 409,000 | NA |
|  |  | *Hybrid/ACS* | 103 | 1,325,000 | 27,000 |
|  |  | *UniversalPCR/ACS* | 207 | 1,426,000 | Dominated |
|  |  | *NoIntervention* | 538 | 1,461,000 | Dominated |
|  |  | *SxScreen/PCR/Hospital* | 125 | 1,604,000 | Dominated |
|  |  | *Hybrid/Hospital* | 100 | 2,368,000 | 348,000 |
|  |  | *UniversalPCR/Hospital* | 207 | 2,631,000 | Dominated |
|  |  | *UniversalPCR/TempHousing* | 95 | 17,500,000 | 3,223,000 |
|  | 80 |  |  |  |  |
|  |  | *SxScreen/PCR/ACS* | 137 | 409,000 | NA |
|  |  | *Hybrid/ACS* | 103 | 1,325,000 | 27,000 |
|  |  | *UniversalPCR/ACS* | 207 | 1,426,000 | Dominated |
|  |  | *NoIntervention* | 538 | 1,461,000 | Dominated |
|  |  | *SxScreen/PCR/Hospital* | 125 | 1,604,000 | Dominated |
|  |  | *Hybrid/Hospital* | 100 | 2,368,000 | 348,000 |
|  |  | *UniversalPCR/Hospital* | 207 | 2,631,000 | Dominated |
|  |  | *UniversalPCR/TempHousing* | 95 | 22,963,000 | 4,387,000 |
|  | 100 |  |  |  |  |
|  |  | *SxScreen/PCR/ACS* | 137 | 409,000 | NA |
|  |  | *Hybrid/ACS* | 103 | 1,325,000 | 27,000 |
|  |  | *UniversalPCR/ACS* | 207 | 1,426,000 | Dominated |
|  |  | *NoIntervention* | 538 | 1,461,000 | Dominated |
|  |  | *SxScreen/PCR/Hospital* | 125 | 1,604,000 | Dominated |
|  |  | *Hybrid/Hospital* | 100 | 2,368,000 | 348,000 |
|  |  | *UniversalPCR/Hospital* | 207 | 2,631,000 | Dominated |
|  |  | *UniversalPCR/TempHousing* | 95 | 28,427,000 | 5,551,000 |

**eTable11 continued. One-way sensitivity analysis on daily cost of temporary housing.**

| **Effective reproduction number (R_e_)** | **Daily cost of temporary housing, 2020 USD** | **Strategy** | **Cumulative infections, n** | **Total cost *,**  **2020 USD** | **Incr. cost per case prevented *, 2020 USD** |
| --- | --- | --- | --- | --- | --- |
| 1.3 | 120 |  |  |  |  |
|  |  | *SxScreen/PCR/ACS* | 137 | 409,000 | NA |
|  |  | *Hybrid/ACS* | 103 | 1,325,000 | 27,000 |
|  |  | *UniversalPCR/ACS* | 207 | 1,426,000 | Dominated |
|  |  | *NoIntervention* | 538 | 1,461,000 | Dominated |
|  |  | *SxScreen/PCR/Hospital* | 125 | 1,604,000 | Dominated |
|  |  | *Hybrid/Hospital* | 100 | 2,368,000 | 348,000 |
|  |  | *UniversalPCR/Hospital* | 207 | 2,631,000 | Dominated |
|  |  | *UniversalPCR/TempHousing* | 95 | 33,890,000 | 6,715,000 |
|  | 141 |  |  |  |  |
|  | base case | *SxScreen/PCR/ACS* | 137 | 409,000 | NA |
|  |  | *Hybrid/ACS* | 103 | 1,325,000 | 27,000 |
|  |  | *UniversalPCR/ACS* | 207 | 1,426,000 | Dominated |
|  |  | *NoIntervention* | 538 | 1,461,000 | Dominated |
|  |  | *SxScreen/PCR/Hospital* | 125 | 1,604,000 | Dominated |
|  |  | *Hybrid/Hospital* | 100 | 2,368,000 | 382,000 |
|  |  | *UniversalPCR/Hospital* | 207 | 2,631,000 | Dominated |
|  |  | *UniversalPCR/TempHousing* | 95 | 38,974,000 | 6,854,000 |
|  | 160 |  |  |  |  |
|  |  | *SxScreen/PCR/ACS* | 137 | 409,000 | NA |
|  |  | *Hybrid/ACS* | 103 | 1,325,000 | 27,000 |
|  |  | *UniversalPCR/ACS* | 207 | 1,426,000 | Dominated |
|  |  | *NoIntervention* | 538 | 1,461,000 | Dominated |
|  |  | *SxScreen/PCR/Hospital* | 125 | 1,604,000 | Dominated |
|  |  | *Hybrid/Hospital* | 100 | 2,368,000 | 348,000 |
|  |  | *UniversalPCR/Hospital* | 207 | 2,631,000 | Dominated |
|  |  | *UniversalPCR/TempHousing* | 95 | 44,816,000 | 9,042,000 |

**eTable11 continued. One-way sensitivity analysis on daily cost of temporary housing.**

| **Effective reproduction number (R_e_)** | **Daily cost of temporary housing, 2020 USD** | **Strategy** | **Cumulative infections, n** | **Total cost *,**  **2020 USD** | **Incr. cost per case prevented *, 2020 USD** |
| --- | --- | --- | --- | --- | --- |
| 0.9 | 0 |  |  |  |  |
|  |  | *SxScreen/PCR/ACS* | 85 | 264,000 | NA |
|  |  | *NoIntervention* | 174 | 540,000 | Dominated |
|  |  | *UniversalPCR/TempHousing* | 71 | 1,089,000 | 60,000 |
|  |  | *SxScreen/PCR/Hospital* | 82 | 1,113,000 | Dominated |
|  |  | *UniversalPCR/ACS* | 94 | 1,226,000 | Dominated |
|  |  | *Hybrid/ACS* | 71 | 1,240,000 | 667,000 |
|  |  | *UniversalPCR/Hospital* | 95 | 1,901,000 | Dominated |
|  |  | *Hybrid/Hospital* | 71 | 2,004,000 | Dominated |
|  | 20 |  |  |  |  |
|  |  | *SxScreen/PCR/ACS* | 85 | 264,000 | NA |
|  |  | *NoIntervention* | 174 | 540,000 | Dominated |
|  |  | *SxScreen/PCR/Hospital* | 82 | 1,113,000 | Dominated |
|  |  | *UniversalPCR/ACS* | 94 | 1,226,000 | Dominated |
|  |  | *Hybrid/ACS* | 71 | 1,240,000 | 70,000 |
|  |  | *UniversalPCR/Hospital* | 95 | 1,901,000 | Dominated |
|  |  | *Hybrid/Hospital* | 71 | 2,004,000 | Dominated |
|  |  | *UniversalPCR/TempHousing* | 71 | 6,552,000 | Dominated |
|  | 40 |  |  |  |  |
|  |  | *SxScreen/PCR/ACS* | 85 | 264,000 | NA |
|  |  | *NoIntervention* | 174 | 540,000 | Dominated |
|  |  | *SxScreen/PCR/Hospital* | 82 | 1,113,000 | Dominated |
|  |  | *UniversalPCR/ACS* | 94 | 1,226,000 | Dominated |
|  |  | *Hybrid/ACS* | 71 | 1,240,000 | 70,000 |
|  |  | *UniversalPCR/Hospital* | 95 | 1,901,000 | Dominated |
|  |  | *Hybrid/Hospital* | 71 | 2,004,000 | Dominated |
|  |  | *UniversalPCR/TempHousing* | 71 | 12,016,000 | Dominated |

**eTable11 continued. One-way sensitivity analysis on daily cost of temporary housing.**

| **Effective reproduction number (R_e_)** | **Daily cost of temporary housing, 2020 USD** | **Strategy** | **Cumulative infections, n** | **Total cost *,**  **2020 USD** | **Incr. cost per case prevented *, 2020 USD** |
| --- | --- | --- | --- | --- | --- |
| 0.9 | 60 |  |  |  |  |
|  |  | *SxScreen/PCR/ACS* | 85 | 264,000 | NA |
|  |  | *NoIntervention* | 174 | 540,000 | Dominated |
|  |  | *SxScreen/PCR/Hospital* | 82 | 1,113,000 | Dominated |
|  |  | *UniversalPCR/ACS* | 94 | 1,226,000 | Dominated |
|  |  | *Hybrid/ACS* | 71 | 1,240,000 | 70,000 |
|  |  | *UniversalPCR/Hospital* | 95 | 1,901,000 | Dominated |
|  |  | *Hybrid/Hospital* | 71 | 2,004,000 | Dominated |
|  |  | *UniversalPCR/TempHousing* | 71 | 17,479,000 | Dominated |
|  | 80 |  |  |  |  |
|  |  | *SxScreen/PCR/ACS* | 85 | 264,000 | NA |
|  |  | *NoIntervention* | 174 | 540,000 | Dominated |
|  |  | *SxScreen/PCR/Hospital* | 82 | 1,113,000 | Dominated |
|  |  | *UniversalPCR/ACS* | 94 | 1,226,000 | Dominated |
|  |  | *Hybrid/ACS* | 71 | 1,240,000 | 70,000 |
|  |  | *UniversalPCR/Hospital* | 95 | 1,901,000 | Dominated |
|  |  | *Hybrid/Hospital* | 71 | 2,004,000 | Dominated |
|  |  | *UniversalPCR/TempHousing* | 71 | 22,943,000 | Dominated |
|  | 100 |  |  |  |  |
|  |  | *SxScreen/PCR/ACS* | 85 | 264,000 | NA |
|  |  | *NoIntervention* | 174 | 540,000 | Dominated |
|  |  | *SxScreen/PCR/Hospital* | 82 | 1,113,000 | Dominated |
|  |  | *UniversalPCR/ACS* | 94 | 1,226,000 | Dominated |
|  |  | *Hybrid/ACS* | 71 | 1,240,000 | 70,000 |
|  |  | *UniversalPCR/Hospital* | 95 | 1,901,000 | Dominated |
|  |  | *Hybrid/Hospital* | 71 | 2,004,000 | Dominated |
|  |  | *UniversalPCR/TempHousing* | 71 | 28,406,000 | Dominated |

**eTable11 continued. One-way sensitivity analysis on daily cost of temporary housing.**

| **Effective reproduction number (R_e_)** | **Daily cost of temporary housing, 2020 USD** | **Strategy** | **Cumulative infections, n** | **Total cost *,**  **2020 USD** | **Incr. cost per case prevented *, 2020 USD** |
| --- | --- | --- | --- | --- | --- |
| 0.9 | 120 |  |  |  |  |
|  |  | *SxScreen/PCR/ACS* | 85 | 264,000 | NA |
|  |  | *NoIntervention* | 174 | 540,000 | Dominated |
|  |  | *SxScreen/PCR/Hospital* | 82 | 1,113,000 | Dominated |
|  |  | *UniversalPCR/ACS* | 94 | 1,226,000 | Dominated |
|  |  | *Hybrid/ACS* | 71 | 1,240,000 | 70,000 |
|  |  | *UniversalPCR/Hospital* | 95 | 1,901,000 | Dominated |
|  |  | *Hybrid/Hospital* | 71 | 2,004,000 | Dominated |
|  |  | *UniversalPCR/TempHousing* | 71 | 33,870,000 | Dominated |
|  | 141 |  |  |  |  |
|  | base case | *SxScreen/PCR/ACS* | 85 | 264,000 | NA |
|  |  | *NoIntervention* | 174 | 540,000 | Dominated |
|  |  | *SxScreen/PCR/Hospital* | 82 | 1,113,000 | Dominated |
|  |  | *UniversalPCR/ACS* | 94 | 1,226,000 | Dominated |
|  |  | *Hybrid/ACS* | 71 | 1,240,000 | 71,000 |
|  |  | *UniversalPCR/Hospital* | 95 | 1,901,000 | Dominated |
|  |  | *Hybrid/Hospital* | 71 | 2,004,000 | Dominated |
|  |  | *UniversalPCR/TempHousing* | 71 | 38,954,000 | Dominated |
|  | 160 |  |  |  |  |
|  |  | *SxScreen/PCR/ACS* | 85 | 264,000 | NA |
|  |  | *NoIntervention* | 174 | 540,000 | Dominated |
|  |  | *SxScreen/PCR/Hospital* | 82 | 1,113,000 | Dominated |
|  |  | *UniversalPCR/ACS* | 94 | 1,226,000 | Dominated |
|  |  | *Hybrid/ACS* | 71 | 1,240,000 | 70,000 |
|  |  | *UniversalPCR/Hospital* | 95 | 1,901,000 | Dominated |
|  |  | *Hybrid/Hospital* | 71 | 2,004,000 | Dominated |
|  |  | *UniversalPCR/TempHousing* | 71 | 44,797,000 | Dominated |

Abbreviations: ACS, alternate care sites; COVID-19, coronavirus disease 2019; Dominated, less clinically effective and more costly than an alternative strategy, or a combination of two alternative strategies;^14^ Incr., incremental; *PCR,* polymerase chain reaction; *SxScreen,* symptom screen; *TempHousing,* temporary housing; *UniversalPCR*, universal polymerase chain reaction test for everyone; USD, United States dollars.

Strategies are listed in order of ascending costs, per convention of cost-effectiveness analysis.

* Costs were rounded to the nearest thousands.

**eTable12. Two-way sensitivity analysis on PCR sensitivity and PCR cost for the *SxScreen/PCR/ACS* and *Hybrid/ACS* strategies.**

| **PCR sensitivity for people with mild/mod. illness, %** | **PCR test cost, 2020 USD** | **Strategy** | **Cumulative infections, n** | **Total cost *,**  **2020 USD** | **Incr. cost per case prevented *, 2020 USD** |
| --- | --- | --- | --- | --- | --- |
| **Effective reproduction number (R_e_) = 2.6** | | | | | |
| 60 | 5 | *Hybrid/ACS* | 1,159 | 3,075,000 | NA |
|  |  | *SxScreen/PCR/ACS* | 1,346 | 3,406,000 | Dominated |
|  | 10 | *Hybrid/ACS* | 1,159 | 3,180,000 | NA |
|  |  | *SxScreen/PCR/ACS* | 1,346 | 3,412,000 | Dominated |
|  | 25 | *SxScreen/PCR/ACS* | 1,346 | 3,430,000 | NA |
|  |  | *Hybrid/ACS* | 1,159 | 3,494,000 | 300 |
|  | 51 | *SxScreen/PCR/ACS* | 1,346 | 3,462,000 | NA |
|  | (base case) | *Hybrid/ACS* | 1,159 | 4,039,000 | 3,000 |
|  | 75 | *SxScreen/PCR/ACS* | 1,346 | 3,491,000 | NA |
|  |  | *Hybrid/ACS* | 1,159 | 4,542,000 | 6,000 |
| 70 | 5 | *Hybrid/ACS* | 985 | 2,711,000 | NA |
|  |  | *SxScreen/PCR/ACS* | 1,239 | 3,246,000 | Dominated |
|  | 10 | *Hybrid/ACS* | 985 | 2,815,000 | NA |
|  |  | *SxScreen/PCR/ACS* | 1,239 | 3,251,000 | Dominated |
|  | 25 | *Hybrid/ACS* | 985 | 3,127,000 | NA |
|  |  | *SxScreen/PCR/ACS* | 1,239 | 3,267,000 | Dominated |
|  | 51 | *SxScreen/PCR/ACS* | 1,239 | 3,267,000 | NA |
|  | (base case) | *Hybrid/ACS* | 985 | 3,628,000 | 1,000 |
|  | 75 | *SxScreen/PCR/ACS* | 1,239 | 3,320,000 | NA |
|  |  | *Hybrid/ACS* | 985 | 4,168,000 | 3,000 |

**eTable12 continued. Two-way sensitivity analysis on PCR sensitivity and PCR cost for the *SxScreen/PCR/ACS* and *Hybrid/ACS* strategies.**

| **PCR sensitivity for people with mild/mod. illness, %** | **PCR test cost, 2020 USD** | **Strategy** | **Cumulative infections, n** | **Total cost *,**  **2020 USD** | **Incr. cost per case prevented *, 2020 USD** |
| --- | --- | --- | --- | --- | --- |
| **Effective reproduction number (R_e_) = 2.6** | | | | | |
| 80 | 5 | *Hybrid/ACS* | 825 | 2,372,000 | NA |
|  |  | *SxScreen/PCR/ACS* | 1,154 | 3,085,000 | Dominated |
|  | 10 | *Hybrid/ACS* | 825 | 2,475,000 | NA |
|  |  | *SxScreen/PCR/ACS* | 1,154 | 3,090,000 | Dominated |
|  | 25 | *Hybrid/ACS* | 825 | 2,786,000 | NA |
|  |  | *SxScreen/PCR/ACS* | 1,154 | 3,104,000 | Dominated |
|  | 51 | *SxScreen/PCR/ACS* | 1,154 | 3,128,000 | NA |
|  | (base case) | *Hybrid/ACS* | 825 | 3,324,000 | 1,000 |
|  | 75 | *SxScreen/PCR/ACS* | 1,154 | 3,150,000 | NA |
|  |  | *Hybrid/ACS* | 825 | 3,820,000 | 2,000 |
| 90 | 5 | *Hybrid/ACS* | 668 | 2,019,000 | NA |
|  |  | *SxScreen/PCR/ACS* | 1,067 | 2,903,000 | Dominated |
|  | 10 | *Hybrid/ACS* | 668 | 2,122,000 | NA |
|  |  | *SxScreen/PCR/ACS* | 1,067 | 2,907,000 | Dominated |
|  | 25 | *Hybrid/ACS* | 668 | 2,431,000 | NA |
|  |  | *SxScreen/PCR/ACS* | 1,067 | 2,919,000 | Dominated |
|  | 51 | *SxScreen/PCR/ACS* | 1,067 | 2,940,000 | NA |
|  | (base case) | *Hybrid/ACS* | 668 | 2,966,000 | 100 |
|  | 75 | *SxScreen/PCR/ACS* | 1,067 | 2,960,000 | NA |
|  |  | *Hybrid/ACS* | 668 | 3,459,000 | 1,000 |

**eTable12 continued. Two-way sensitivity analysis on PCR sensitivity and PCR cost for the *SxScreen/PCR/ACS* and *Hybrid/ACS* strategies.**

| **PCR sensitivity for people with mild/mod. illness, %** | **PCR test cost, 2020 USD** | **Strategy** | **Cumulative infections, n** | **Total cost *,**  **2020 USD** | **Incr. cost per case prevented *, 2020 USD** |
| --- | --- | --- | --- | --- | --- |
| **Effective reproduction number (R_e_) = 1.3** | | | | | |
| 60 | 5 | *SxScreen/PCR/ACS* | 148 | 413,000 | NA |
|  |  | *Hybrid/ACS* | 116 | 433,000 | 1,000 |
|  | 10 | *SxScreen/PCR/ACS* | 148 | 414,000 | NA |
|  |  | *Hybrid/ACS* | 116 | 534,000 | 4,000 |
|  | 25 | *SxScreen/PCR/ACS* | 148 | 417,000 | NA |
|  |  | *Hybrid/ACS* | 116 | 839,000 | 13,000 |
|  | 51 | *SxScreen/PCR/ACS* | 148 | 420,000 | NA |
|  | (base case) | *Hybrid/ACS* | 116 | 1,366,000 | 30,000 |
|  | 75 | *SxScreen/PCR/ACS* | 148 | 425,000 | NA |
|  |  | *Hybrid/ACS* | 116 | 1,852,000 | 45,000 |
| 70 | 5 | *SxScreen/PCR/ACS* | 137 | 402,000 | NA |
|  |  | *Hybrid/ACS* | 103 | 409,000 | 200 |
|  | 10 | *SxScreen/PCR/ACS* | 137 | 403,000 | NA |
|  |  | *Hybrid/ACS* | 103 | 510,000 | 3,000 |
|  | 25 | *SxScreen/PCR/ACS* | 137 | 405,000 | NA |
|  |  | *Hybrid/ACS* | 103 | 814,000 | 12,000 |
|  | 51 | *SxScreen/PCR/ACS* | 137 | 409,000 | NA |
|  | (base case) | *Hybrid/ACS* | 103 | 1,341,000 | 27,000 |
|  | 75 | *SxScreen/PCR/ACS* | 137 | 413,000 | NA |
|  |  | *Hybrid/ACS* | 103 | 1,827,000 | 41,000 |

**eTable12 continued. Two-way sensitivity analysis on PCR sensitivity and PCR cost for the *SxScreen/PCR/ACS* and *Hybrid/ACS* strategies.**

| **PCR sensitivity for people with mild/mod. illness, %** | **PCR test cost, 2020 USD** | **Strategy** | **Cumulative infections, n** | **Total cost *,**  **2020 USD** | **Incr. cost per case prevented *, 2020 USD** |
| --- | --- | --- | --- | --- | --- |
| **Effective reproduction number (R_e_) = 1.3** | | | | | |
| 80 | 5 | *SxScreen/PCR/ACS* | 127 | 386,000 | NA |
|  |  | *Hybrid/ACS* | 92 | 400,000 | 400 |
|  | 10 | *SxScreen/PCR/ACS* | 127 | 386,000 | NA |
|  |  | *Hybrid/ACS* | 92 | 501,000 | 3,000 |
|  | 25 | *SxScreen/PCR/ACS* | 127 | 388,000 | NA |
|  |  | *Hybrid/ACS* | 92 | 805,000 | 12,000 |
|  | 51 | *SxScreen/PCR/ACS* | 127 | 392,000 | NA |
|  | (base case) | *Hybrid/ACS* | 92 | 1,331,000 | 27,000 |
|  | 75 | *SxScreen/PCR/ACS* | 127 | 395,000 | NA |
|  |  | *Hybrid/ACS* | 92 | 1,817,000 | 41,000 |
| 90 | 5 | *SxScreen/PCR/ACS* | 121 | 378,000 | NA |
|  |  | *Hybrid/ACS* | 85 | 388,000 | 300 |
|  | 10 | *SxScreen/PCR/ACS* | 121 | 378,000 | NA |
|  |  | *Hybrid/ACS* | 85 | 489,000 | 3,000 |
|  | 25 | *SxScreen/PCR/ACS* | 121 | 380,000 | NA |
|  |  | *Hybrid/ACS* | 85 | 793,000 | 12,000 |
|  | 51 | *SxScreen/PCR/ACS* | 121 | 383,000 | NA |
|  | (base case) | *Hybrid/ACS* | 85 | 1,319,000 | 27,000 |
|  | 75 | *SxScreen/PCR/ACS* | 121 | 386,000 | NA |
|  |  | *Hybrid/ACS* | 85 | 1,805,000 | 40,000 |

**eTable12 continued. Two-way sensitivity analysis on PCR sensitivity and PCR cost for the *SxScreen/PCR/ACS* and *Hybrid/ACS* strategies.**

| **PCR sensitivity for people with mild/mod. illness, %** | **PCR test cost, 2020 USD** | **Strategy** | **Cumulative infections, n** | **Total cost *,**  **2020 USD** | **Incr. cost per case prevented *, 2020 USD** |
| --- | --- | --- | --- | --- | --- |
| **Effective reproduction number (R_e_) = 0.9** | | | | | |
| 60 | 5 | *SxScreen/PCR/ACS* | 87 | 263,000 | NA |
|  |  | *Hybrid/ACS* | 76 | 330,000 | 6,000 |
|  | 10 | *SxScreen/PCR/ACS* | 87 | 264,000 | NA |
|  |  | *Hybrid/ACS* | 76 | 432,000 | 15,000 |
|  | 25 | *SxScreen/PCR/ACS* | 87 | 266,000 | NA |
|  |  | *Hybrid/ACS* | 76 | 735,000 | 41,000 |
|  | 51 | *SxScreen/PCR/ACS* | 87 | 272,200 | NA |
|  | (base case) | *Hybrid/ACS* | 76 | 1,261,000 | 88,000 |
|  | 75 | *SxScreen/PCR/ACS* | 87 | 271,000 | NA |
|  |  | *Hybrid/ACS* | 76 | 1,747,000 | 128,000 |
| 70 | 5 | *SxScreen/PCR/ACS* | 85 | 259,000 | NA |
|  |  | *Hybrid/ACS* | 71 | 325,000 | 5,000 |
|  | 10 | *SxScreen/PCR/ACS* | 85 | 260,000 | NA |
|  |  | *Hybrid/ACS* | 71 | 426,000 | 12,000 |
|  | 25 | *SxScreen/PCR/ACS* | 85 | 262,000 | NA |
|  |  | *Hybrid/ACS* | 71 | 730,000 | 34,000 |
|  | 51 | *SxScreen/PCR/ACS* | 85 | 264,000 | NA |
|  | (base case) | *Hybrid/ACS* | 71 | 1,256,000 | 72,000 |
|  | 75 | *SxScreen/PCR/ACS* | 85 | 267,000 | NA |
|  |  | *Hybrid/ACS* | 71 | 1,742,000 | 107,000 |

**eTable12 continued. Two-way sensitivity analysis on PCR sensitivity and PCR cost for the *SxScreen/PCR/ACS* and *Hybrid/ACS* strategies.**

| **PCR sensitivity for people with mild/mod. illness, %** | **PCR test cost, 2020 USD** | **Strategy** | **Cumulative infections, n** | **Total cost *,**  **2020 USD** | **Incr. cost per case prevented *, 2020 USD** |
| --- | --- | --- | --- | --- | --- |
| **Effective reproduction number (R_e_) = 0.9** | | | | | |
| 80 | 5 | *SxScreen/PCR/ACS* | 82 | 255,000 | NA |
|  |  | *Hybrid/ACS* | 68 | 326,000 | 5,000 |
|  | 10 | *SxScreen/PCR/ACS* | 82 | 255,000 | NA |
|  |  | *Hybrid/ACS* | 68 | 427,000 | 13,000 |
|  | 25 | *SxScreen/PCR/ACS* | 82 | 257,000 | NA |
|  |  | *Hybrid/ACS* | 68 | 731,000 | 35,000 |
|  | 51 | *SxScreen/PCR/ACS* | 82 | 259,000 | NA |
|  | (base case) | *Hybrid/ACS* | 68 | 1,257,000 | 74,000 |
|  | 75 | *SxScreen/PCR/ACS* | 82 | 262,000 | NA |
|  |  | *Hybrid/ACS* | 68 | 1,742,000 | 109,000 |
| 90 | 5 | *SxScreen/PCR/ACS* | 80 | 259,000 | NA |
|  |  | *Hybrid/ACS* | 65 | 322,000 | 4,000 |
|  | 10 | *SxScreen/PCR/ACS* | 80 | 259,000 | NA |
|  |  | *Hybrid/ACS* | 65 | 423,000 | 11,000 |
|  | 25 | *SxScreen/PCR/ACS* | 80 | 261,000 | NA |
|  |  | *Hybrid/ACS* | 65 | 727,000 | 32,000 |
|  | 51 | *SxScreen/PCR/ACS* | 80 | 263,000 | NA |
|  | (base case) | *Hybrid/ACS* | 65 | 1,252,000 | 67,000 |
|  | 75 | *SxScreen/PCR/ACS* | 80 | 265,000 | NA |
|  |  | *Hybrid/ACS* | 65 | 1,738,000 | 100,000 |

Abbreviations: ACS, alternate care sites; COVID-19, coronavirus disease 2019; Dominated, less clinically effective and more costly than an alternative strategy, or a combination of two alternative strategies;^14^ Incr., incremental; mild/mod., mild/moderate; *PCR,* polymerase chain reaction; *TempHousing,* temporary housing; *UniversalPCR*, universal polymerase chain reaction test for everyone; USD, United States dollars; *SxScreen,* symptom screen.

Strategies are listed in order of ascending costs, per convention of cost-effectiveness analysis.

* Costs were rounded to the nearest thousands.

**eTable13. Two-way sensitivity analysis on PCR testing frequency and PCR cost for the *SxScreen/PCR/ACS* and *Hybrid/ACS* strategies.**

| **PCR testing frequency** | **PCR test cost, 2020 USD** | **Strategy** | **Cumulative infections, n** | **Total cost *,**  **2020 USD** | **Incr. cost per case prevented *, 2020 USD** |
| --- | --- | --- | --- | --- | --- |
| **Effective reproduction number (R_e_) = 2.6** | | | | | |
| Every 30 days | 5 | *Hybrid/ACS* | 1,179 | 3,034,000 | NA |
|  |  | *SxScreen/PCR/ACS* | 1,244 | 3,246,000 | Dominated |
|  | 10 | *Hybrid/ACS* | 1,179 | 3,095,000 | NA |
|  |  | *SxScreen/PCR/ACS* | 1,244 | 3,251,000 | Dominated |
|  | 25 | *SxScreen/PCR/ACS* | 1,244 | 3,267,000 | NA |
|  |  | *Hybrid/ACS* | 1,179 | 3,277,000 | 200 |
|  | 51 | *SxScreen/PCR/ACS* | 1,244 | 3,295,000 | NA |
|  | (base case) | *Hybrid/ACS* | 1,179 | 3,594,000 | 5,000 |
|  | 75 | *SxScreen/PCR/ACS* | 1,244 | 3,320,000 | NA |
|  |  | *Hybrid/ACS* | 1,179 | 3,886,000 | 9,000 |
| Every 14 days | 5 | *Hybrid/ACS* | 989 | 2,711,000 | NA |
| (base case) |  | *SxScreen/PCR/ACS* | 1,244 | 3,246,000 | Dominated |
|  | 10 | *Hybrid/ACS* | 989 | 2,815,000 | NA |
|  |  | *SxScreen/PCR/ACS* | 1,244 | 3,251,000 | Dominated |
|  | 25 | *Hybrid/ACS* | 989 | 3,127,000 | NA |
|  |  | *SxScreen/PCR/ACS* | 1,244 | 3,267,000 | Dominated |
|  | 51 | *SxScreen/PCR/ACS* | 1,244 | 3,295,000 | NA |
|  | (base case) | *Hybrid/ACS* | 989 | 3,669,000 | 1,000 |
|  | 75 | *SxScreen/PCR/ACS* | 1,244 | 3,320,000 | NA |
|  |  | *Hybrid/ACS* | 989 | 4,168,000 | 3,000 |

**eTable13 continued. Two-way sensitivity analysis on PCR testing frequency and PCR cost for the *SxScreen/PCR/ACS* and *Hybrid/ACS* strategies.**

| **PCR testing frequency** | **PCR test cost, 2020 USD** | **Strategy** | **Cumulative infections, n** | **Total cost *,**  **2020 USD** | **Incr. cost per case prevented *, 2020 USD** |
| --- | --- | --- | --- | --- | --- |
| **Effective reproduction number (R_e_) = 2.6** | | | | | |
| Every 7 days | 5 | *Hybrid/ACS* | 700 | 2,171,000 | NA |
|  |  | *SxScreen/PCR/ACS* | 1,244 | 3,246,000 | Dominated |
|  | 10 | *Hybrid/ACS* | 700 | 2,350,000 | NA |
|  |  | *SxScreen/PCR/ACS* | 1,244 | 3,251,000 | Dominated |
|  | 25 | *Hybrid/ACS* | 700 | 2,887,000 | NA |
|  |  | *SxScreen/PCR/ACS* | 1,244 | 3,267,000 | Dominated |
|  | 51 | *SxScreen/PCR/ACS* | 1,244 | 3,295,000 | NA |
|  | (base case) | *Hybrid/ACS* | 700 | 3,818,000 | 1,000 |
|  | 75 | *SxScreen/PCR/ACS* | 1,244 | 3,320,000 | NA |
|  |  | *Hybrid/ACS* | 700 | 4,677,000 | 2,000 |
| Every 3 days | 5 | *Hybrid/ACS* | 334 | 1,438,000 | NA |
|  |  | *SxScreen/PCR/ACS* | 1,244 | 3,246,000 | Dominated |
|  | 10 | *Hybrid/ACS* | 334 | 1,783,000 | NA |
|  |  | *SxScreen/PCR/ACS* | 1,244 | 3,251,000 | Dominated |
|  | 25 | *Hybrid/ACS* | 334 | 2,817,000 | NA |
|  |  | *SxScreen/PCR/ACS* | 1,244 | 3,267,000 | Dominated |
|  | 51 | *SxScreen/PCR/ACS* | 1,244 | 3,295,000 | NA |
|  | (base case) | *Hybrid/ACS* | 334 | 4,609,000 | 1,000 |
|  | 75 | *SxScreen/PCR/ACS* | 1,244 | 3,320,000 | NA |
|  |  | *Hybrid/ACS* | 334 | 6,263,000 | 3,000 |

**eTable13 continued. Two-way sensitivity analysis on PCR testing frequency and PCR cost for the *SxScreen/PCR/ACS* and *Hybrid/ACS* strategies.**

| **PCR testing frequency** | **PCR test cost, 2020 USD** | **Strategy** | **Cumulative infections, n** | **Total cost *,**  **2020 USD** | **Incr. cost per case prevented *, 2020 USD** |
| --- | --- | --- | --- | --- | --- |
| **Effective reproduction number (R_e_) = 1.3** | | | | | |
| Every 30 days | 5 | *Hybrid/ACS* | 115 | 387,000 | NA |
|  |  | *SxScreen/PCR/ACS* | 137 | 402,000 | Dominated |
|  | 10 | *SxScreen/PCR/ACS* | 137 | 403,000 | NA |
|  |  | *Hybrid/ACS* | 115 | 442,000 | 2,000 |
|  | 25 | *SxScreen/PCR/ACS* | 137 | 405,000 | NA |
|  |  | *Hybrid/ACS* | 115 | 609,000 | 9,000 |
|  | 51 | *SxScreen/PCR/ACS* | 137 | 409,000 | NA |
|  | (base case) | *Hybrid/ACS* | 115 | 896,000 | 22,000 |
|  | 75 | *SxScreen/PCR/ACS* | 137 | 413,000 | NA |
|  |  | *Hybrid/ACS* | 115 | 1,162,000 | 34,000 |
| Every 14 days | 5 | *SxScreen/PCR/ACS* | 137 | 402,000 | NA |
| (base case) |  | *Hybrid/ACS* | 103 | 409,000 | 200 |
|  | 10 | *SxScreen/PCR/ACS* | 137 | 403,000 | NA |
|  |  | *Hybrid/ACS* | 103 | 510,000 | 3,000 |
|  | 25 | *SxScreen/PCR/ACS* | 137 | 405,000 | NA |
|  |  | *Hybrid/ACS* | 103 | 814,000 | 12,000 |
|  | 51 | *SxScreen/PCR/ACS* | 137 | 409,000 | NA |
|  | (base case) | *Hybrid/ACS* | 103 | 1,341,000 | 27,000 |
|  | 75 | *SxScreen/PCR/ACS* | 137 | 413,000 | NA |
|  |  | *Hybrid/ACS* | 103 | 1,827,000 | 41,000 |

**eTable13 continued. Two-way sensitivity analysis on PCR testing frequency and PCR cost for the *SxScreen/PCR/ACS* and *Hybrid/ACS* strategies.**

| **PCR testing frequency** | **PCR test cost, 2020 USD** | **Strategy** | **Cumulative infections, n** | **Total cost *,**  **2020 USD** | **Incr. cost per case prevented *, 2020 USD** |
| --- | --- | --- | --- | --- | --- |
| **Effective reproduction number (R_e_) = 1.3** | | | | | |
| Every 7 days | 5 | *SxScreen/PCR/ACS* | 137 | 402,000 | NA |
|  |  | *Hybrid/ACS* | 91 | 473,000 | 2,000 |
|  | 10 | *SxScreen/PCR/ACS* | 137 | 403,000 | NA |
|  |  | *Hybrid/ACS* | 91 | 653,000 | 5,000 |
|  | 25 | *SxScreen/PCR/ACS* | 137 | 405,000 | NA |
|  |  | *Hybrid/ACS* | 91 | 1,191,000 | 17,000 |
|  | 51 | *SxScreen/PCR/ACS* | 137 | 409,000 | NA |
|  | (base case) | *Hybrid/ACS* | 91 | 2,123,000 | 37,000 |
|  | 75 | *SxScreen/PCR/ACS* | 137 | 413,000 | NA |
|  |  | *Hybrid/ACS* | 91 | 2,984,000 | 56,000 |
| Every 3 days | 5 | *SxScreen/PCR/ACS* | 137 | 402,000 | NA |
|  |  | *Hybrid/ACS* | 79 | 625,000 | 4,000 |
|  | 10 | *SxScreen/PCR/ACS* | 137 | 403,000 | NA |
|  |  | *Hybrid/ACS* | 79 | 972,000 | 10,000 |
|  | 25 | *SxScreen/PCR/ACS* | 137 | 405,000 | NA |
|  |  | *Hybrid/ACS* | 79 | 2,013,000 | 28,000 |
|  | 51 | *SxScreen/PCR/ACS* | 137 | 409,000 | NA |
|  | (base case) | *Hybrid/ACS* | 79 | 3,817,000 | 59,000 |
|  | 75 | *SxScreen/PCR/ACS* | 137 | 413,000 | NA |
|  |  | *Hybrid/ACS* | 79 | 5,483,000 | 88,000 |

**eTable13 continued. Two-way sensitivity analysis on PCR testing frequency and PCR cost for the *SxScreen/PCR/ACS* and *Hybrid/ACS* strategies.**

| **PCR testing frequency** | **PCR test cost, 2020 USD** | **Strategy** | **Cumulative infections, n** | **Total cost *,**  **2020 USD** | **Incr. cost per case prevented *, 2020 USD** |
| --- | --- | --- | --- | --- | --- |
| **Effective reproduction number (R_e_) = 0.9** | | | | | |
| Every 30 days | 5 | *SxScreen/PCR/ACS* | 85 | 259,000 | NA |
|  |  | *Hybrid/ACS* | 74 | 279,000 | 2,000 |
|  | 10 | *SxScreen/PCR/ACS* | 85 | 260,000 | NA |
|  |  | *Hybrid/ACS* | 74 | 334,000 | 7,000 |
|  | 25 | *SxScreen/PCR/ACS* | 85 | 262,000 | NA |
|  |  | *Hybrid/ACS* | 74 | 499,000 | 21,000 |
|  | 51 | *SxScreen/PCR/ACS* | 85 | 264,000 | NA |
|  | (base case) | *Hybrid/ACS* | 74 | 785,000 | 46,000 |
|  | 75 | *SxScreen/PCR/ACS* | 85 | 267,000 | NA |
|  |  | *Hybrid/ACS* | 74 | 1,050,000 | 69,000 |
| Every 14 days | 5 | *SxScreen/PCR/ACS* | 85 | 259,000 | NA |
| (base case) |  | *Hybrid/ACS* | 71 | 325,000 | 5,000 |
|  | 10 | *SxScreen/PCR/ACS* | 85 | 260,000 | NA |
|  |  | *Hybrid/ACS* | 71 | 426,000 | 12,000 |
|  | 25 | *SxScreen/PCR/ACS* | 85 | 262,000 | NA |
|  |  | *Hybrid/ACS* | 71 | 730,000 | 34,000 |
|  | 51 | *SxScreen/PCR/ACS* | 85 | 264,000 | NA |
|  | (base case) | *Hybrid/ACS* | 71 | 1,256,000 | 72,000 |
|  | 75 | *SxScreen/PCR/ACS* | 85 | 267,000 | NA |
|  |  | *Hybrid/ACS* | 71 | 1,742,000 | 107,000 |

**eTable13 continued. Two-way sensitivity analysis on PCR testing frequency and PCR cost for the *SxScreen/PCR/ACS* and *Hybrid/ACS* strategies.**

| **PCR testing frequency** | **PCR test cost, 2020 USD** | **Strategy** | **Cumulative infections, n** | **Total cost *,**  **2020 USD** | **Incr. cost per case prevented *, 2020 USD** |
| --- | --- | --- | --- | --- | --- |
| **Effective reproduction number (R_e_) = 0.9** | | | | | |
| Every 7 days | 5 | *SxScreen/PCR/ACS* | 85 | 259,000 | NA |
|  |  | *Hybrid/ACS* | 68 | 406,000 | 8,000 |
|  | 10 | *SxScreen/PCR/ACS* | 85 | 260,000 | NA |
|  |  | *Hybrid/ACS* | 68 | 585,000 | 19,000 |
|  | 25 | *SxScreen/PCR/ACS* | 85 | 262,000 | NA |
|  |  | *Hybrid/ACS* | 68 | 1,124,000 | 49,000 |
|  | 51 | *SxScreen/PCR/ACS* | 85 | 264,000 | NA |
|  | (base case) | *Hybrid/ACS* | 68 | 2,057,000 | 103,000 |
|  | 75 | *SxScreen/PCR/ACS* | 85 | 267,000 | NA |
|  |  | *Hybrid/ACS* | 68 | 2,918,000 | 152,000 |
| Every 3 days | 5 | *SxScreen/PCR/ACS* | 85 | 259,000 | NA |
|  |  | *Hybrid/ACS* | 64 | 569,000 | 15,000 |
|  | 10 | *SxScreen/PCR/ACS* | 85 | 260,000 | NA |
|  |  | *Hybrid/ACS* | 64 | 916,000 | 31,000 |
|  | 25 | *SxScreen/PCR/ACS* | 85 | 262,000 | NA |
|  |  | *Hybrid/ACS* | 64 | 1,958,000 | 80,000 |
|  | 51 | *SxScreen/PCR/ACS* | 85 | 264,000 | NA |
|  | (base case) | *Hybrid/ACS* | 64 | 3,763,000 | 164,000 |
|  | 75 | *SxScreen/PCR/ACS* | 85 | 267,000 | NA |
|  |  | *Hybrid/ACS* | 64 | 5,429,000 | 242,000 |

Abbreviations: ACS, alternate care sites; COVID-19, coronavirus disease 2019; Dominated, less clinically effective and more costly than an alternative strategy, or a combination of two alternative strategies;^14^ Incr., incremental; *PCR,* polymerase chain reaction; *SxScreen,* symptom screen; *TempHousing,* temporary housing; *UniversalPCR*, universal polymerase chain reaction test for everyone; USD, United States dollars.

Strategies are listed in order of ascending costs, per convention of cost-effectiveness analysis.

* Costs were rounded to the nearest thousands.

**eTable14. Results of an analysis of management strategies for people experiencing sheltered homelessness during the COVID-19 pandemic for a cohort of 1,000 adults experiencing sheltered homelessness.**

| **Strategy** | **Cumulative infections, n** | **Peak daily hospital bed use, n** | **Hospital costs *, 2020 USD** | **ACS costs *, 2020 USD** | **Total costs *,**  **2020 USD** | **Costs compared with *NoIntervention **, 2020 USD** | **Incr. cost per case prevented *, 2020 USD** |
| --- | --- | --- | --- | --- | --- | --- | --- |
| **Effective reproduction number (R_e_) = 2.6** | | | | | | | |
| *SxScreen/PCR/ACS* | 549 | 2 | 343,000 | 1,080,000 | 1,447,000 | - 1,254,000 | NA |
| *Hybrid/ACS* | 436 | 2 | 266,000 | 878,000 | 1,607,000 | - 1,094,000 | 1,000 |
| *UniversalPCR/ACS* | 744 | 4 | 499,000 | 885,000 | 1,835,000 | - 866,000 | Dominated |
| *NoIntervention* | 865 | 28 | 2,692,000 | NA | 2,701,000 | NA | Dominated |
| *Hybrid/Hospital* | 428 | 35 | 4,941,000 | NA | 5,404,000 | + 2,703,000 | Dominated |
| *SxScreen/PCR/Hospital* | 502 | 41 | 5,567,000 | NA | 5,589,000 | + 2,888,000 | Dominated |
| *UniversalPCR/Hospital* | 743 | 50 | 5,269,000 | NA | 5,719,000 | + 3,018,000 | Dominated |
| *UniversalPCR/TempHousing* | 167 | 1 | 107,000 | NA | 17,325,000 | + 14,624,000 | 58,000 |
| **Effective reproduction number (R_e_) = 1.3** | | | | | | | |
| *SxScreen/PCR/ACS* | 61 | 0 | 43,000 | 135,000 | 181,000 | - 466,000 | NA |
| *Hybrid/ACS* | 46 | 0 | 30,000 | 106,000 | 587,000 | - 60,000 | 27,000 |
| *UniversalPCR/ACS* | 92 | 0 | 61,000 | 122,000 | 632,000 | - 15,000 | Dominated |
| *NoIntervention* | 238 | 4 | 645,000 | NA | 647,000 | NA | Dominated |
| *SxScreen/PCR/Hospital* | 56 | 10 | 707,000 | NA | 710,000 | + 63,000 | Dominated |
| *Hybrid/Hospital* | 44 | 10 | 30,000 | NA | 1,049,000 | + 402,000 | 382,000 |
| *UniversalPCR/Hospital* | 92 | 9 | 716,000 | NA | 1,165,000 | + 518,000 | Dominated |
| *UniversalPCR/TempHousing* | 42 | 4 | 35,000 | NA | 17,260,000 | + 16,613,000 | 6,853,000 |

**eTable14 continued. Results of an analysis of management strategies for people experiencing sheltered homelessness during the COVID-19 pandemic for a cohort of 1,000 adults experiencing sheltered homelessness.**

| **Strategy** | **Cumulative infections, n** | **Peak daily hospital bed use, n** | **Hospital costs *, 2020 USD** | **ACS costs *, 2020 USD** | **Total costs *,**  **2020 USD** | **Costs compared with *NoIntervention **, 2020 USD** | **Incr. cost per case prevented *, 2020 USD** |
| --- | --- | --- | --- | --- | --- | --- | --- |
| **Effective reproduction number (R_e_) = 0.9** | | | | | | | |
| *SxScreen/PCR/ACS* | 38 | 0 | 27,000 | 88,000 | 117,000 | - 426,000 | NA |
| *NoIntervention* | 77 | 2 | 238,000 | NA | 239,000 | - 304,000 | Dominated |
| *SxScreen/PCR/Hospital* | 36 | 9 | 491,000 | NA | 493,000 | - 50,000 | Dominated |
| *UniversalPCR/ACS* | 42 | 0 | 27,000 | 66,000 | 543,000 | NA | Dominated |
| *Hybrid/ACS* | 32 | 0 | 23,000 | 76,000 | 549,000 | + 6,000 | 71,000 |
| *UniversalPCR/Hospital* | 42 | 9 | 393,000 | NA | 842,000 | + 299,000 | Dominated |
| *Hybrid/Hospital* | 31 | 10 | 437,000 | NA | 888,000 | + 345,000 | Dominated |
| *UniversalPCR/TempHousing* | 31 | 0 | 26,000 | NA | 17,252,000 | + 16,709,000 | Dominated |

Abbreviations: ACS, alternate care sites; Dominated, less clinically effective and more costly than an alternative strategy, or with a higher incremental cost-effectiveness ratio than another strategy;^14^ Incr., incremental; *SxScreen*, symptom screen; *TempHousing*, temporary housing; *UniversalPCR*, polymerase chain reaction test for everyone; USD, United States dollars.

Strategies are listed in order of ascending costs, per convention of cost-effectiveness analysis. City-specific population sizes of sheltered homeless adults are as follows: 2,258 in Boston, 1,343 in Houston, 2,053 in Chicago, 2,770 in Philadelphia, 3,556 in Seattle, 6,945 in Los Angeles, and 31,805 in New York. ^15^

* Costs were rounded to the nearest thousands.

**eTable15. Total infections and component costs of different management strategies for adults experiencing sheltered homelessness in Boston during the COVID-19 pandemic at 4 months (n=2,258).**

| **Strategy** | **Cumulative infections, n** | **ICU bed costs, ***  **2020 USD** | **Hospital (non-ICU) bed costs, ***  **2020 USD** | **Test costs, * 2020 USD** | **ACS costs, * 2020 USD** | **Temporary housing costs, ***  **2020 USD** | **Total costs, * 2020 USD** |
| --- | --- | --- | --- | --- | --- | --- | --- |
| **Effective reproduction number (R_e_) = 2.6** | | | | | | | |
| *SxScreen/PCR/ACS* | 1,239 | 349,000 | 425,000 | 54,000 | 2,439,000 | NA | 3,267,000 |
| *Hybrid/ACS* | 985 | 270,000 | 330,000 | 1,046,000 | 1,982,000 | NA | 3,628,000 |
| *UniversalPCR/ACS* | 1,681 | 517,000 | 610,000 | 1,017,000 | 1,999,000 | NA | 4,143,000 |
| *NoIntervention* | 1,954 | 621,000 | 5,458,000 | 19,000 | NA | NA | 6,098,000 |
| *Hybrid/Hospital* | 967 | 275,000 | 10,882,000 | 1,045,000 | NA | NA | 12,202,000 |
| *SxScreen/PCR/Hospital* | 1,133 | 318,000 | 12,253,000 | 49,000 | NA | NA | 12,620,000 |
| *UniversalPCR/Hospital* | 1,679 | 501,000 | 11,396,000 | 1,017,000 | NA | NA | 12,914,000 |
| *UniversalPCR/TempHousing* | 376 | 115,000 | 127,000 | 1,015,000 | NA | 37,862,000 | 39,119,000 |
| **Effective reproduction number (R_e_) = 1.3** | | | | | | | |
| *SxScreen/PCR/ACS* | 137 | 46,000 | 51,000 | 8,000 | 304,000 | NA | 409,000 |
| *Hybrid/ACS* | 103 | 30,000 | 37,000 | 1,018,000 | 240,000 | NA | 1,325,000 |
| *UniversalPCR/ACS* | 207 | 63,000 | 74,000 | 1,014,000 | 275,000 | NA | 1,426,000 |
| *NoIntervention* | 538 | 145,000 | 1,311,000 | 5,000 | NA | NA | 1,461,000 |
| *SxScreen/PCR/Hospital* | 125 | 39,000 | 1,558,000 | 7,000 | NA | NA | 1,604,000 |
| *Hybrid/Hospital* | 100 | 31,000 | 1,319,000 | 1,018,000 | NA | NA | 2,368,000 |
| *UniversalPCR/Hospital* | 207 | 62,000 | 1,555,000 | 1,014,000 | NA | NA | 2,631,000 |
| *UniversalPCR/TempHousing* | 95 | 41,000 | 38,000 | 1,016,000 | NA | 37,879,000 | 38,974,000 |

**eTable15 continued. Total infections and component costs of different management strategies for adults experiencing sheltered homelessness in Boston during the COVID-19 pandemic at 4 months.**

| **Strategy** | **Cumulative infections, n** | **ICU bed costs, ***  **2020 USD** | **Hospital (non-ICU) bed costs, ***  **2020 USD** | **Test costs, * 2020 USD** | **ACS costs, * 2020 USD** | **Temporary housing costs, ***  **2020 USD** | **Total costs, * 2020 USD** |
| --- | --- | --- | --- | --- | --- | --- | --- |
| **Effective reproduction number (R_e_) = 0.9** | | | | | | | |
| *SxScreen/PCR/ACS* | 85 | 28,000 | 33,000 | 5,000 | 198,000 | NA | 264,000 |
| *NoIntervention* | 174 | 50,000 | 488,000 | 2,000 | NA | NA | 540,000 |
| *SxScreen/PCR/Hospital* | 82 | 31,000 | 1,077,000 | 5,000 | NA | NA | 1,113,000 |
| *UniversalPCR/ACS* | 94 | 28,000 | 34,000 | 1,014,000 | 150,000 | NA | 1,226,000 |
| *Hybrid/ACS* | 71 | 25,000 | 26,000 | 1,017,000 | 172,000 | NA | 1,240,000 |
| *UniversalPCR/Hospital* | 95 | 31,000 | 856,000 | 1,014,000 | NA | NA | 1,901,000 |
| *Hybrid/Hospital* | 71 | 24,000 | 963,000 | 1,017,000 | NA | NA | 2,004,000 |
| *UniversalPCR/TempHousing* | 71 | 28,000 | 30,000 | 1,015,000 | NA | 37,881,000 | 38,954,000 |

Abbreviations: ACS, alternate care sites; COVID-19, coronavirus disease 2019; ICU, intensive care units; *PCR,* polymerase chain reaction; *UniversalPCR*, universal polymerase chain reaction test for everyone; *SxScreen,* symptom screen; *TempHousing,* temporary housing; USD, United States dollars.

Strategies are listed in order of ascending costs, per convention of cost-effectiveness analysis.

* All costs were rounded to the nearest thousands.

**eTable16. One-way sensitivity analysis on PCR testing frequency for the *Hybrid/ACS* strategy.**

| **Strategy** | **Cumulative infections, n** | **Incr. infections prevented, n** | **Total cost *,**  **2020 USD** | **Cost compared with *NoIntervention* *, 2020 USD** | **Incr. cost per case prevented *, 2020 USD** |
| --- | --- | --- | --- | --- | --- |
| **Effective reproduction number (R_e_) = 2.6** | | | | | |
| *SxScreen/PCR/ACS* | 1,239 | NA | 3,267,000 | - 2,831,000 | NA |
| *Hybrid/ACS - PCR once every 30 days* | 1,189 | 50 | 3,587,000 | - 2,511,000 | Dominated |
| *Hybrid/ACS - PCR once every 14 days* | 985 | 205 | 3,628,000 | - 2,470,000 | Dominated |
| *Hybrid/ACS - PCR once every 7 days* | 696 | 289 | 3,849,000 | - 2,249,000 | 1,000 |
| *Hybrid/ACS - PCR once every 3 days* | 340 | 356 | 4,633,000 | - 1,465,000 | 2,000 |
| *NoIntervention* | 1,954 | - 1,614 | 6,098,000 | NA | Dominated |
| **Effective reproduction number (R_e_) = 1.3** | | | | | |
| *SxScreen/PCR/ACS* | 137 | NA | 409,000 | - 1,052,000 | NA |
| *Hybrid/ACS - PCR once every 30 days* | 114 | 24 | 891,000 | - 570,000 | 20,000 |
| *Hybrid/ACS - PCR once every 14 days* | 103 | 10 | 1,325,000 | - 136,000 | 41,000 |
| *NoIntervention* | 538 | - 435 | 1,461,000 | NA | Dominated |
| *Hybrid/ACS - PCR once every 7 days* | 92 | 446 | 2,123,000 | + 662,000 | 72,000 |
| *Hybrid/ACS - PCR once every 3 days* | 81 | 11 | 3,808,000 | + 2,347,000 | 153,000 |
| **Effective reproduction number (R_e_) = 0.9** | | | | | |
| *SxScreen/PCR/ACS* | 85 | NA | 264,000 | - 276,000 | NA |
| *NoIntervention* | 174 | - 89 | 540,000 | NA | Dominated |
| *Hybrid/ACS - PCR once every 30 days* | 76 | 98 | 785,000 | + 245,000 | 60,000 |
| *Hybrid/ACS - PCR once every 14 days* | 71 | 5 | 1,240,000 | + 700,000 | 88,000 |
| *Hybrid/ACS - PCR once every 7 days* | 70 | 1 | 2,061,000 | + 1,521,000 | Dominated |
| *Hybrid/ACS - PCR once every 3 days* | 66 | 4 | 3,772,000 | + 3,232,000 | 484,000 |

Abbreviations: ACS, alternate care sites; Dominated, less clinically effective and more costly than an alternative strategy, or a combination of two alternative strategies; ^14^ Incr., incremental; *PCR,* polymerase chain reaction; *SxScreen,* symptom screen; *TempHousing,* temporary housing; USD, United States dollars.

Strategies are listed in order of ascending costs, per convention of cost-effectiveness analysis.

* All costs were rounded to the nearest thousands.

**LEGENDS TO FIGURES**

**eFigure1.** Illustration of health states and illness paths in the CEACOV model.

CEACOV simulates individuals transitioning between the states of susceptibility to severe acute respiratory syndrome coronavirus 2 (SARS-CoV-2), infection with SARS-CoV-2 and coronavirus illness 2019 (COVID-19), recovery from COVID-19, and death. Susceptible individuals face a daily probability of infection. After acquiring SARS-CoV-2 infection, individuals may progress through the following health states: pre-infectious latency, asymptomatic infection, mild/moderate illness, severe illness, critical illness, recuperation, and recovered.

After being infected with SARS-CoV-2, a susceptible individual first transitions to the pre-infectious latency stage. Then, the individual has an age-dependent probability of progressing along one of four “paths,” culminating in either asymptomatic infection, mild/moderate illness, severe illness, or critical illness. Before a reaching a more advanced illness state, individuals must first transition through intermediate states (e.g., those destined for severe illness must first pass through the asymptomatic infection state and the mild/moderate illness state).

Abbreviations: Asympt., asymptomatic; CEACOV, Clinical and Economic Analysis of COVID-19 interventions model; Mild/Mod., mild/moderate.

**eFigure2.** Flow diagrams of management strategies for people experiencing sheltered homelessness in Boston during the COVID-19 pandemic.

We assess eight strategies for testing and management:

1) ***NoIntervention:*** Only basic infection control practices are implemented in shelters.

2) ***SxScreen/PCR/Hospital:*** CDC-recommended symptom screening daily in shelters.^6^ Screen-negative individuals remain in shelters. Screen-positive individuals are sent to the hospital for PCR testing. PCR-positive individuals remain in hospital; PCR-negative individuals return to shelter.

3) ***SxScreen/PCR/ACS:*** CDC-recommended symptom screening daily in shelters. Screen-negative individuals remain in shelters. Screen-positive individuals are sent to an ACS for people under investigation, where they undergo PCR testing and await results. PCR-positive individuals with mild/moderate illness are transferred to ACSs for confirmed COVID-19 cases. PCR-negative individuals return to shelter.

4) ***UniversalPCR/Hospital:*** Universal PCR testing every 2 weeks in shelters. Those with symptoms at the time of testing await results at the hospital; individuals without symptoms await results in shelters. PCR-negative individuals return to or stay in shelters. PCR-positive individuals, regardless of illness severity, remain in or are sent to the hospital.

5) ***UniversalPCR/ACS:*** Universal PCR testing every 2 weeks in shelters. Those with symptoms at the time of testing are sent to an ACS for people under investigation while awaiting results; individuals without symptoms await results in shelters. PCR-negative individuals return to or stay in shelters. PCR-positive individuals with mild/moderate illness are transferred to ACSs for confirmed COVID-19 cases.

6) ***UniversalPCR/TempHousing:*** All shelter residents are pre-emptively moved to temporary housing for the duration of the 4-month period. Universal PCR testing occurs every 2 weeks. PCR-positive individuals with mild/moderate illness remain in temporary housing and are transferred to the hospital if they progress to severe or critical disease.

7) ***Hybrid/Hospital:*** This includes the *SxScreen/PCR/Hospital* strategy and adds shelter-based universal PCR testing every 2 weeks for those without symptoms.

8) ***Hybrid/ACS:*** This includes the *SxScreen/PCR/ACS* strategy and adds shelter-based universal PCR testing every 2 weeks for those without symptoms.

In all 8 strategies, people with severe or critical illness are sent to the hospital. Individuals are eligible for repeat PCR testing after 5 days since their most recent negative test. See eFigure2 for details.

Abbreviations: ACS, alternate care sites; COVID, coronavirus illness 2019; *PCR,* polymerase chain reaction; *Sx Screen,* symptom screen; *TempHousing,* temporary housing; *UniversalPCR*, universal polymerase chain reaction test for everyone.

**eFigure1.**

**eFigure2.**

1. ***NoIntervention***

2. ***SxScreen/PCR/Hospital***

3. ***SxScreen/PCR/ACS***

4. ***UniversalPCR/Hospital***

5. ***UniversalPCR/ACS***

6. ***UniversalPCR/TempHousing***

7. ***Hybrid/Hospital***

8. ***Hybrid/ACS***
